# Exploring metabolic changes in gout – Insights from a genetic study

**DOI:** 10.1101/2024.06.12.24308809

**Authors:** Ville Salo, Johannes Kettunen, FinnGen, Estonian Biobank Research Team, Eeva Sliz

## Abstract

Gout is the most common inflammatory joint disease caused by the crystallization of urate inside the joints. Patients with gout typically have abnormal blood lipid and sugar levels, which are associated with cardiovascular diseases. However, the relationship between gout and metabolic changes is unclear. Our goal was two-fold: to identify new gout risk factors using genome-wide association analysis and subsequently to investigate the effects of the identified risk alleles on metabolic measures in the bloodstream. We performed a genome-wide meta-analysis for gout in the FinnGen project, the Estonian Biobank, and the UK Biobank, encompassing a total of 992,583 individuals, including 17,972 gout cases. Given that gout is commonly recognized as a disease affecting the elderly, and males specifically, we further explored age- and sex-stratified genetic associations in FinnGen (10,885 cases and 366,392 controls). Finally, we determined the metabolomic consequences of the gout risk-increasing alleles using data from a large metabolomics GWAS. In the meta-analysis, we observed 32 genome-wide significant (P<5×10^−8^) loci, one of which was novel. In the age- and sex-stratified analyses, we additionally identified one novel gout-associated locus in the male subgroup. The metabolomic findings suggested that the majority of the gout risk alleles primarily affected urate concentration in the bloodstream but not the concentrations of lipids and other metabolites. Therefore, it appears that the associations between gout and metabolic factors at the population level are likely explained by shared lifestyle risk factors. In conclusion, our study sheds new light on the genetic architecture of gout and adds to the growing body of evidence supporting the role of urate, but not other metabolic measures, including lipoprotein lipids and glucose, as a key risk factor for developing gout.

## Introduction

Previous genome-wide association studies (GWASs)^1^ have identified numerous loci associated, with gout, a common inflammatory joint disease caused by the crystallization of urate inside the joints^2^. These findings have provided valuable information about key gout-related risk factors, such as hyperuricemia and joint diseases^3–10^. However, the genetic factors underlying gout remain incompletely understood. In this study, we provide additional information about the genetic factors behind gout using GWAS and several downstream analyses. In addition, as gout is considered a disease affecting older people and males^2^, we also performed both age- and sex-stratified GWASs to identify possible age of onset or sex-specific loci. Importantly, because gout is frequently diagnosed with abnormal blood lipid and sugar levels^11^, we explored whether certain metabolic changes are systemically associated with gout risk alleles using data from a large nuclear magnetic resonance (NMR) metabolome GWAS^12^. Overall, our aim was to create a more comprehensive picture of gout as a disease by combining genetic and metabolic information, allowing us to gain insights into the underlying biological pathways.

## Materials & Methods

### Study populations

This work is part of the FinnGen research project^13^. The main goal of FinnGen is a better understanding of disease mechanisms by combining genomic and health data from up to over 500,000 Finns. The FinnGen project has the necessary ethical and prior permits for biobank research (Supplementary Note), and all participants have given their written consent to biobank research either in connection with sample donation or when participating in older research projects, the materials of which have been transferred to Finnish biobanks with the consent of Fimea, the National Supervisory Authority for Welfare and Health. The FinnGen data freeze R9 data used in the study contains 10,885 gout cases and 366,392 control cases (Table S1.1).

In the genome-wide meta-analysis, we utilised data from Estonian and UK biobanks. The Estonian biobank includes samples from 200,000 voluntary participants, representing more than 15% of the Estonian adult population. In the meta-analysis, there were 2,578 gout cases and 192,600 controls from the Estonian Biobank. The UK biobank material consists of samples collected during the years 2006-2010. Samples were collected from hundreds of thousands of people aged 40–69 from all over Great Britain. There were 4,509 gout patients and 415,619 controls in the UK biobank, adding up to 17,972 gout cases and 974,611 controls in the meta-analysis.

### Phenotype descriptions

*The International Classification of Diseases, Tenth Revision* (ICD-10), and *Ninth Revision* (ICD-9) codes were used to characterize phenotypes. The Hospital Discharge Registry and the Cause of Death Registry provided data for the codes. Patients with ICD-10 codes M10.0 and M10.9 and ICD-9 codes 274.0 and 274.9 were categorized as gout cases. Patients without a record of these ICD codes were categorized as controls.

In the age-stratified GWAS completed in FinnGen, the cases were further classified into two age groups: those who were diagnosed before the age of 50 years (988 cases, 366,329 controls, Table S2.1) and those who were diagnosed at the age of 50 or older (9897 cases, 366,329 controls, Table S2.2; Figure S1). If the patient had multiple diagnoses, only the initial diagnosis of gout was considered to determine the age of diagnosis. The control group was not stratified by age, and, thus, the same control group as in the original FinnGen GWAS was used in both age-stratified GWASs. In addition, we ran additional sex-stratified GWAS analyses comparing gout-affected males (8142 cases, Table S2.3) and females (2743 controls, Table S2.4) to same-sex controls (N=158,265 and N=208,27, respectively; Figure S2).

### Genotyping, imputation & quality control

Illumina and Affymetrix DNA microarrays were used to determine genotypes. Genotype data were quality controlled to exclude variants with a low Hardy-Weinberg equilibrium (HWE) p-value (<1×10^−6^), minor allele count (MAC) below three, and high missingness (cut-off 2%), as well as individuals with high genotype missingness (cut-off 5%), high levels of heterozygosity (±4 SD), non-Finnish ancestry, and individuals whose sex did not match the genotype data. Samples were pre-phased using Eagle 2.3.5, with the number of conditioning haplotypes set to 20,000. Beagle 4.1 was used for genotype imputation. The reference panel was Finnish SISu v3. The imputation protocol has been described at (dx.doi.org/10.17504/protocols.io.nmndc5e.) Finally, post-imputation quality control was carried out to exclude variants with imputation information values less than 0.6.

### GWAS and meta-analysis

An additive GWAS was performed using the Regenie program^14^. The GWAS was adjusted for age, sex, and the first 10 genetic principal components. Sex was not included as a covariate in the sex-stratified GWAS.

The METAL software^15^ was used to do an inverse variance-weighted fixed-effect meta-analysis of the GWAS results from the three populations. Variant data from the Estonian and UK biobanks were converted from the hg19 to the hg38 prior to the meta-analysis.

### Characterization of the association signals

We defined a locus as a window of 2 MB (± 1,000,000 bases from the association lead variant) containing at least one variant associated with gout at P<5×10^−8^. For the loci that had not been reported in association with gout in prior studies, we determined a potential candidate gene with a relevant biological function with the help of literature and databases (Genbank^16^, Uniprot^17^, GTEx-Portal^18^). With age- and sex-stratified GWASs that were completed in FinnGen, we used a 0.5% allele frequency filter to mitigate the risk of false positive findings.

### Functional annotations

Functional annotations of the results of the meta-analysis were completed using FUMA^19^. Functional settings were selected for the analysis, in which case the program uses functional information for mapping. Positional mapping was also performed, and for that, SNP markers were selected for the region of exons or introns, affecting post-transcriptional modifications, and involved in gene regulation. Optional options included filtering SNP markers based on CADD results, which provided additional information on the possible harmful effects of SNP markers. In addition, filtering of SNP markers was performed based on the RegulomeDB results, and in turn, information was obtained based on gene expression data and epigenomics about the possible functions of SNP markers affecting gene regulation. In the mapping, gene expression data was also utilized, and eQTL mapping was performed. For this, the relevant tissue types were selected, which were kidney, liver, and blood (GTEx v8), and the focus was only on genes involved in protein coding. A MAGMA analysis^20^ providing gene-level results was also performed in the run, which focuses on gene-level information, unlike GWAS, where associations are reported at the variant level. In the gene-set analysis MAGMA uses curated gene sets and GO annotations from MSigDB^21^. A 10kb gene window and selected GTEx v8 tissue variants were put into the analysis. Also, the HLA region was left out of the analysis.

### eQTL colocalizations

We further investigated colocalizations between gout association signals and gene expression: approximate Bayesian factor analyses were performed with the ‘coloc.abf’ function found in the ‘coloc’ R-library^22^. Colocalizations were investigated for a total of 39 genes. Candidate genes and genes closest to the association signal were selected for the analysis if the closest gene was a different gene than the candidate gene. For the analysis, we selected tissues that are relevant for gout, namely kidney cortex, liver, small intestine, and whole blood. Variant-gene expression associations (GTEx v8), for the analysis were downloaded (01/16/2023) from GTExportal (https://www.gtexportal.org/home/datasets). Colocalizations with posterior probabilities ≥ 0,8 for the variant were considered significant^23^.

### SNP-based heritability and genetic correlations

The LDSC software^24^ was used to calculate the SNP-based heritability estimate and genetic correlations. Heritability estimation was performed using the liability scale, with a sample prevalence of 0.018 and a population prevalence of 0.015 as estimated by Dehlin et al. (2020) for the Nordic adult population^25^. Genetic correlations were calculated between gout and 437 other phenotypes extracted from the GWAS database provided by the MRC Integrative Epidemiology Unit (IEU) (https://gwas.mrcieu.ac.uk/). We used a false discovery rate (FDR)-corrected p-value (pfdr) < 0.05 as the limit for correlations.

### Subgroup effect differences

In FinnGen, calculated whether there were difference in the effects of the lead variants at the gout-associated loci (Table S2.5) between the age- or sex-stratified subgroups, i.e., those diagnosed before the age of 50 years vs. those diagnosed after 50 years of age, and females vs. males. For this, we used a two-tailed test, using subgroup-specific effect estimates of the variants and the corresponding standard errors *((Effect_males-Effect_females)/ sqrt(standarderror_males*^*2*^*+standarderror_females*^*2*^)). A P-value <0.05 was considered the limit of a significant effect difference.

### Conditional analysis at 19q13.**33**

Since the locus on chromosome 19 had been connected to different candidate genes in previous studies^6,9^ and they were located very close to each other, we performed a conditional GWAS to see if the locus harbours multiple independent association signals. For that, the genotype of the gout-associated lead variant (rs150414818) from locus 19q13.33 was selected into the conditional GWAS as a covariate.

### Metabolic associations

To investigate the differences in metabolic associations between the gout risk variants and to determine metabolic changes occurring in association with higher gout risk, we compared the metabolic effects of the gout risk-increasing allele of each variant. Metabolic effects of the gout risk alleles were extracted from our recent metabolomics GWAS^12^: full summary statistics for the study have been made publicly available through the NHGRI-EBI GWAS catalogue (GCST90301941–GCST90302173) and https://www.phpc.cam.ac.uk/ceu/lipids-and-metabolites/. We selected 66 metabolic measures (Table S3), which provide a comprehensive view of the metabolic associations of gout variants. Data from the metabolomics GWAS^12^ were combined with inflammatory marker (https://gwas.mrcieu.ac.uk/files/ukb-d-30710_irnt/ukb-d-30710_irnt_report.html#diagnostics) and urate (https://gwas.mrcieu.ac.uk/files/ukb-d-30880_irnt/ukb-d-30880_irnt_report.html) data extracted from the MR-Base website^26,27^. For the six gout risk-associated variants not found in the metabolomics GWAS data, we estimated proxies using the LDproxy tool available in LDlink applications^28^. Unfortunately, no LD-based proxies were found for two variants; hence, we were not able to determine metabolic associations for rs200864767 and rs143321297. We scaled the metabolic associations relative to the largest absolute effect by dividing the original effects by the largest absolute effect estimate for each variant, thus obtaining effect estimates for all variants between -1 and 1. With the help of scaling, the association profiles better reflect biological and metabolic functions than the original effect estimates, which vary in magnitude^29^. A heatmap was constructed by hierarchical clustering using the scaled metabolic effect estimates of the variants.

## Results

We detected 32 genome-wide significant (P<5×10^−8^) loci associated with gout in the meta-analysis (Figure 1, Table S4, Figure S3). While most loci were reported in previous studies, the association near *CDH8* (*cadherin 8*, Table 1, Figure S4.1) achieved GWAS significance for association with gout for the first time. We estimated LDSC-derived SNP-based heritability to be 0.21 (standard error [SE]= 0.04), suggesting that genetic factors account for 21% of the variation in gout risk. Genomic inflation factor lambda (1.18) suggested minor inflation in the test statistics; given the intercept of 1.04, minor inflation is likely caused by a polygenic signal.

**Table 1.** Novel gout-associated loci. The table reports the lead variants of a novel gout associated locus (P<5×10^−8^, at least 1MB apart) identified in the genome-wide meta-analysis of 17,972 gout cases and 974,611 controls from FinnGen, the Estonian Biobank, and the UK Biobank (above) and a novel locus identified in a genome-wide association study of gout in a male subgroup of FinnGen participants (8,142 cases, 158,265 controls) (below). A full list of gout associated loci identified in the genome-wide meta-analysis can be seen in Table S4.

| Meta-analysis (FinnGen, Estonian Biobank, UK Biobank) |  |  |  |  |  |  |  |  |  |
| --- | --- | --- | --- | --- | --- | --- | --- | --- | --- |
| Locus | Gene | CHR:POS | rsid | EA | OA | p-value | Cohort | OR (95% CI) | EAF |
| 16q21 | CDH8 | 16:63007457 | rs186124051 | T | C | 9.76e-09 | Meta-analysis | 0.76 (0.66-0.86) | 0.02 |
|  |  |  |  |  |  |  | FinnGen | 0.73 (0.61-0.85) | 0.02 |
|  |  |  |  |  |  |  | Estonian Biobank | 0.82 (0.52-1.12) | 0.01 |
|  |  |  |  |  |  |  | UK Biobank | 0.79 (0.61-0.98) | 0.02 |
| Sex-stratified analysis (FinnGen) |  |  |  |  |  |  |  |  |  |
| Locus | Gene | CHR:POS | rsid | EA | OA | p-value | Sex | OR (95% CI) | EAF |
| 17p13.2 | SLC25A11 | 17:4914555 | rs139336593 | T | G | 4.14e-08 | Males | 0.78 (0.69-0.87) | 0.04 |
|  |  |  |  |  |  |  | Females | 1.02 (0.88-1.16) | 0.04 |

Gene, a plausible candidate the biological function of which is likely to explain the gout association; CHR:POS, chromosome, and position (genome build hg38); rsid, SNP markers identification number; EA, effect allele; OA, other allele; p-value, p-value in genome-wide meta-analysis; Cohort, study population or subgroup in sex-stratified analysis; OR (95CI), odds ratio and it's 95% confidence interval, EAF, effect allele frequency.

**Figure 1.**
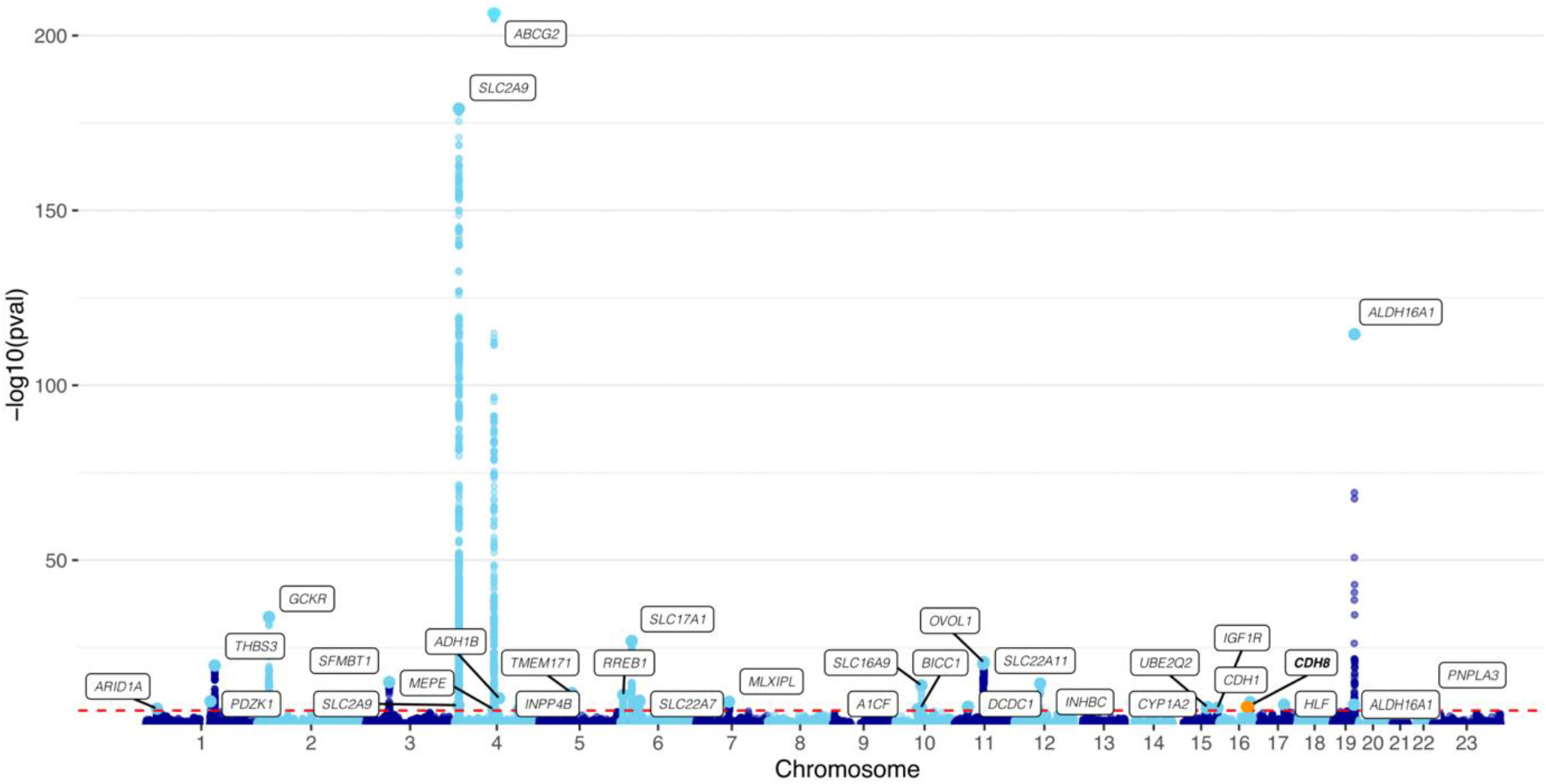
Manhattan plot of the gout associations the meta-analysis of 17,972 cases and 974,611 controls from FinnGen, Estonian Biobank and UK Biobank. A novel gout associated locus is highlighted in orange. Candidate genes at each locus are labelled

In the age- and sex-stratified analyses completed in FinnGen, we discovered a novel locus associated with gout in the male subgroup (Table 1, Figure S4.2) near *solute carrier family 25 member 11* (*SLC25A11*, P=4.14e-8). We also found significant sex differences in the effect size of the lead variant at this locus (P=0.001, Figure S5, Table S5) and four other loci near *ABCG2* (*ATP binding cassette subfamily G member 2), SLC2A9* (*solute carrier family 2 member 9*), *RREB1* (*ras responsive element binding protein 1*) and *SLC17A1* (*solute carrier family 17 member 1*). Significant differences in the effect sizes between age groups were observed for the lead variant located on chromosome 19 near *ALDH16A1* (*aldehyde dehydrogenase 16 family member A1*).

The conditional GWAS was adjusted for the rs150414818 (19:49465749:C:G, missense) genotype, no other genome-wide significant associations emerged on chromosome 19 (Figure S6). This suggests that the association with gout on chromosome 19 spans a larger area and is explained by the missense variant on *ALDH16A1*.

MAGMA gene-set analysis of the meta-analysed data suggested enrichment for pathways responsible for urate metabolism and transportation (Figure S7). In the MAGMA tissue expression analysis testing a positive relationship between tissue-specific gene expression profiles and disease-gene associations, only kidneys reached genome-wide significance (Figure S6). The gout association signal at 1q22 colocalized with the expression of *thrombospondin 3* (*THBS3)* in the small intestine (posterior probability for a single causal variant [PP]=0.997), liver (PP=0.997), and whole blood (PP=0.993). In addition, some other gout risk loci, and association signals colocalized with their candidate genes in certain tissues: gout association signal near *ARID1A* (*AT-rich interaction domain 1)* colocalized with its gene expression in whole blood (PP=0.913). In small intestine gout association signals at 1q21.1 and 4p16.1 colocalized with the expression of respective candidate genes *PDZK1* (*PDZ domain containing 1)* (PP=0.999) and *SLC2A9* (PP=0.812). In addition, the association signals near *SLC17A1* (PP=0.984) and *SLC22A11* (*solute carrier family 22 member 11)* (PP=0.947) gene expressions were colocalized in the liver, while *HLF* (*HLF transcription factor, PAR bZIP family member*) (PP=0.995) expression was colocalized with the gout association signal in the kidneys (Figure S8, Table S6).

We tested for genetic correlations (rg) of gout with other traits. The strongest positive correlations between gout and other traits were observed with M25:other joint disorders’ (Figure 2; rg=0.46) and blood urate level (rg=0.45). We also observed positive correlations with several measures of lipid metabolism, such as mono-unsaturated fatty acids (rg=0.43) and cholesterol-lowering medication (rg=0.43), triglycerides to total lipids ratio in IDL (rg=0.41), triglycerides in large VLDL (rg=0.41) and phospholipids in medium VLDL (rg=0.4). Positive correlations were also observed between gout and I20:Angina pectoris (rg=0.43) and childhood asthma (rg=0.43). The strongest of the negative correlations was total cholesterol in very large high-density lipoprotein (HDL) (rg=-0.50), and another significant negative correlation was the ratio of linoleic acid to total fatty acids (rg=-0.44). The significant correlations (false discovery rate-corrected p-value [pfdr<0.05]) of rg>0.4 or rg<-0.4 are shown in Figure 2, and a full list of results is shown in Table S7.

**Figure 2.**
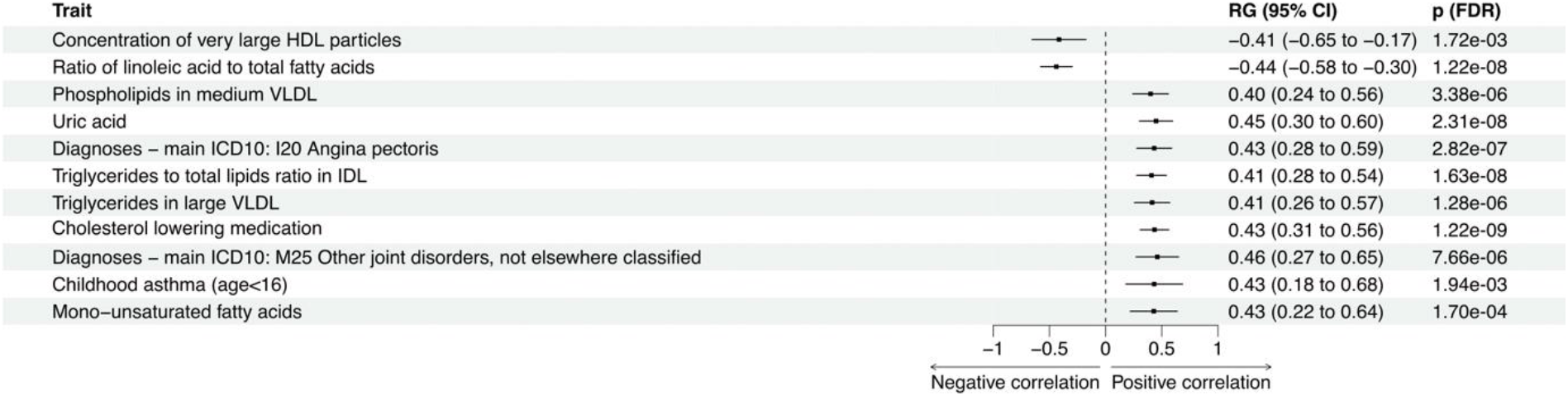
Genetic correlations between gout and other traits. Genetic correlations were calculated using LDSC-software^24^, gout summary statistics from the meta-analysis and, for other traits, data extracted from the GWAS database provided by the MRC Integrative Epidemiology Unit (IEU) (https://gwas.mrcieu.ac.uk/). Only the significant correlations (false discovery rate-corrected p-value [pFDR]<0.05) of rg>0.4 or rg<-0.4 are shown in the figure: genetic correlations between gout and all tested traits can be seen in Table S7. RG (95% CI), genetic correlation coefficient value and it’s 95% confidence intervals; p(FDR), false discovery rate-corrected p-value.

Figure 3 displays the metabolic associations of gout risk alleles. *GCKR rs1260326-T, MLX interacting protein like (MLXIPL) rs12531645-G*, and *ARID1A rs114165349-C* alleles were found to have widespread genome-wide significant metabolic associations. Otherwise, metabolic associations were very modest, and for other alleles, only individual metabolic measures reached genome-wide significance. Urate was the only one of the 66 metabolites examined in the study, the levels of which were consistently higher in association with nearly all gout risk-increasing alleles. However, we observed diverse association strengths in terms of varying effect size magnitudes between alleles. For most of the risk alleles, the effect size for urate was the largest among the studied metabolites, whereas for the alleles showing broader metabolic effects, i.e., *GCKR rs1260326-T, MLXIPL rs12531645-G, ARID1A rs114165349-C*, the effect sizes for urate were modest compared with the effect sizes for other metabolic measures. *CDH8 rs186124051-C* and *SLC25A11 rs139336593-G* were the only exceptions in terms of the effect direction on urate: for these alleles, the effect sizes for urate were negative, although non-significant.

**Figure 3.**
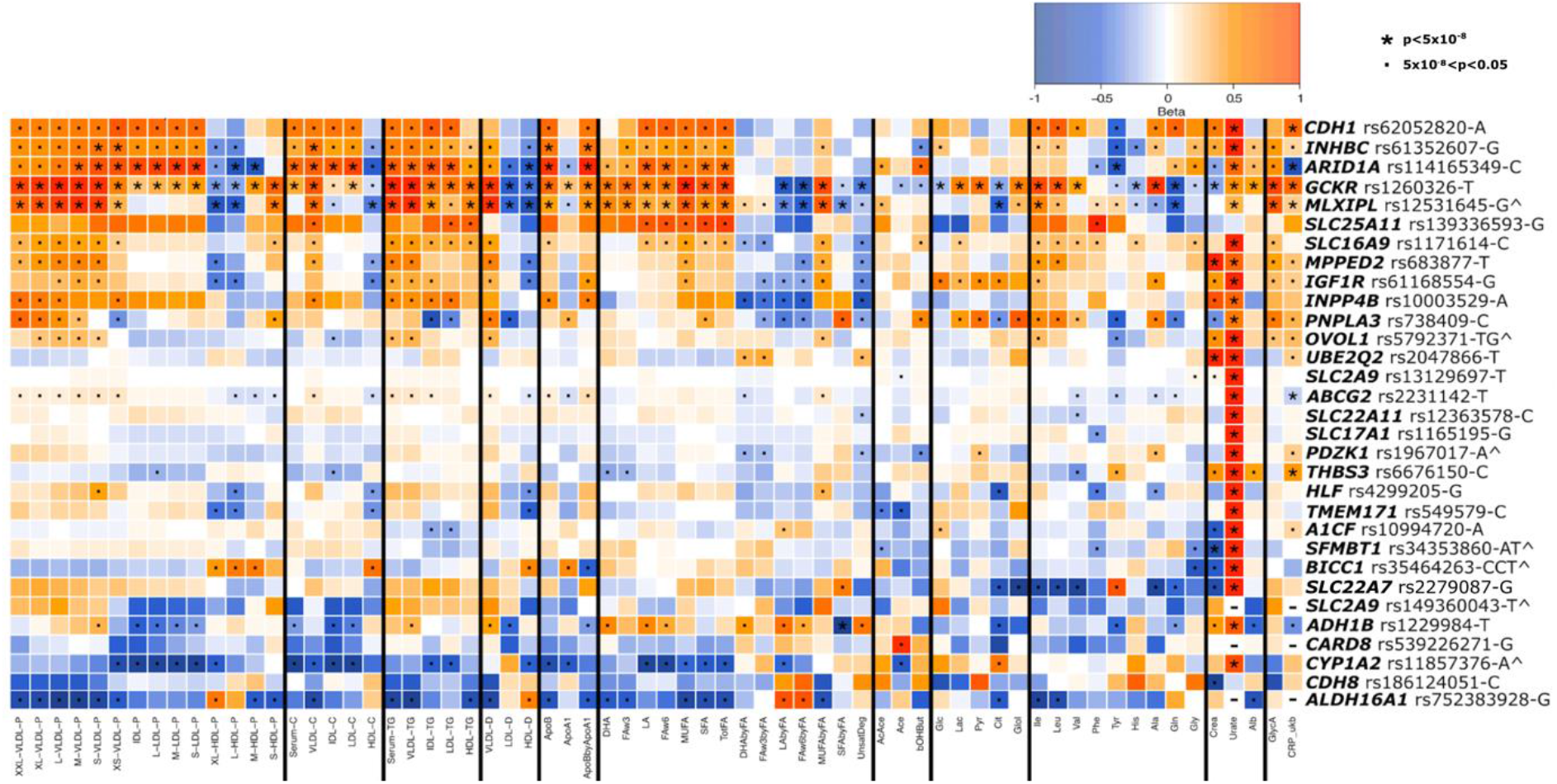
Metabolic association profiles of gout risk-increasing alleles. Metabolic effects are extracted from the metabolomics GWAS^30^, except for uric acid (Urate) and the inflammatory marker c-reactive protein (CRP), which were extracted from the MRBase-website^26,27^ (respective data ids: ukb-d-30880_irnt and ukb-d-30710_irnt). The effect sizes are scaled between -1 and 1 by dividing the original effect sizes by the largest absolute effect estimate of each variant. Genome-wide significant (p<5×10^−8^) associations are marked with an asterisk. Nominally significant associations (5×10^−8^<p<0.05) are marked with a dot. Variants that are LD-based proxies for the originally discovered gout-associated variants are marked with the ^ symbol (Table S8). Missing data are marked with a (-) symbol. The *SLC25A11* variant was associated with gout in a subgroup of men from FinnGen, but its metabolic associations are based on the whole sample of the metabolomics GWAS^30^. The meanings of metabolite abbreviations are listed in Table S3.

*GCKR rs1260326-T, MLXIPL rs12531645-G, ARID1A rs114165349-C* alleles were associated with higher levels of lipids in the bloodstream. *GCKR rs1260326-T* and *MLXIPL rs12531645-G* were highly similar in their metabolic associations and were very strongly positively associated with very low-density lipoproteins (VLDL) and triglyceride levels. These alleles showed positive associations also with all fatty acid measures, and these were the only alleles for which significant positive associations were observed for both inflammatory markers, C-reactive protein (CRP) and glycoprotein acetyls (GlycA). The association profile of *ARID1A rs114165349-C* was slightly different compared to the *GCKR* and *MLXIPL* alleles; this allele was particularly strongly associated with IDL- and LDL-particle levels and cholesterol measures, whereas its association with VLDL particles was limited to the smallest VLDL subfraction. Regarding associations with HDL particles, *ARID1A rs114165349-C* showed negative effects on particles within all HDL subfractions, whereas *GCKR rs1260326-T* and *MLXIPL rs12531645-G* showed positive effects on the particle concentrations within the two smallest HDL subfractions. Interestingly, a very strong negative association with CRP was observed for the *ARID1A rs114165349-C* allele.

## Discussion

In the genome-wide meta-analysis of 17,972 cases and 974,611 controls, we identified 32 gout-associated loci, one of which was novel. In addition, one novel locus was observed in the male subgroup in the sex-stratified analysis, adding up to two novel gout risk loci identified in the present study. Enrichments were observed in the pathways related to urate metabolism, and in the tissue expression analysis, the kidneys were the only tissue significantly associated with gout. We further discovered that gout risk-increasing alleles affected blood metabolic measures in diverse ways, reflecting the underlying molecular pathways. The metabolic effects of gout risk alleles were modest except for the three alleles that had strong and broad metabolic associations. The results support urate’s status as a remarkable metabolic risk factor, as we discovered that, aside from urate, the gout risk alleles were not systemically associated with any other circulating metabolic measures.

### Novel gout risk loci

The novel gout association at 16q21 is located near *CDH8*. It is a member of the cadherin gene family, which is vital for controlling cytoskeletal complexes and sustaining cell-cell contact during cell adhesion^31^. It has previously been nominally associated with chronic kidney disease/kidney failure^32^, making it a potential candidate for explaining the possible gout association. Blaschke et al. found that the *CDH8* gene is normally expressed throughout the fetal stage but not in adult kidneys and that dysregulation of the gene was found to cause morphological changes in the kidneys^33^. Interestingly, however, we did not observe a significant association of the *CDH8 rs186124051-C* variant with circulating urate concentration. Its non-significant effect size for urate association was marginally negative, suggesting that the gout association of this variant is due to factors independent of its effect on urate metabolism. We detected another novel gout association at 17p13.2 near *SLC25A11* that was associated with gout in the male subgroup. *SLC25A11* exchanges α-ketoglutarate and malate across the mitochondrial inner membrane, it is also involved in glutathione (GSH) metabolism^34,35^. Through the malate-aspartate shuttle, *SLC25A11* also has an important role in glycolysis and lactate metabolism, where it transfers reducing equivalents from the cytosol into the mitochondria. Cytosolic NAD^+^ levels must be regenerated to maintain glycolytic flux and for lactate conversion to pyruvate^36^. We observed that the *SLC25A11* allele showed negative associations with both glucose and lactate, suggesting that glycolytic flux and pyruvate conversion to lactate would be operating effectively. However, we observed a marginally negative urate association for the allele, supporting the interpretation that the gout association of this variant is due to factors independent of its effect on urate metabolism. Further studies are needed for both loci, and the latter locus requires replication in another sample to confirm its reliable association with gout.

### Sex and age-specific effects

We observed previously unreported significant sex differences in effect sizes for the lead variants near *SLC25A11, RREB1*, and *SLC17A1*. We further replicated the previously reported sex differences for the lead variants near *SLC2A9, ABCG2*, and *ALDH16A1*^6,37,38^. *ALDH16A1* rs150414818 (missense, Pro476Arg) has been found to disrupt the interaction between *ALDH16A1* and *HPRT1* (*hypoxanthine phosphoribosyltransferase 1*) and thus potentially lead to urate overproduction via the purine salvage pathway^39,40^. *HPRT1* is located on the X-chromosome and may thus contribute to explaining the sex differences in the effect sizes observed for *ALDH16A1* variants. In line with the result of a study investigating early-onset gout^41^, we found that the effect of the *ALDH16A1* variant was significantly larger in the subgroup of patients diagnosed with gout before the age of 50 years than in the subgroup of patients diagnosed after 50 years of age.

### Metabolic associations

Metabolic associations were generally modest, and association directions and magnitudes varied widely between gout risk alleles. Urate was the only metabolite for which the effect size was systemically in the same direction in association with the gout risk alleles, except for the *CHD8 rs186124051-C* and *SLC25A11 rs139336593-G*, for which the non-significant effect sizes were marginally negative. Regarding the other metabolites, the associations varied. For example, we found that some alleles, such as *GCKR rs1260326-T*, are associated with higher and some, including, *ALDH16A1 rs752383928-G*, with lower cholesterol and triglyceride measures, while a vast majority of the gout risk alleles did not show a significant association with these lipid measures. Although abnormal lipid and glucose concentrations are found in association with gout at the population level^42,43^, it seems that many of the molecular pathways leading to gout only affect the urate concentration in the bloodstream without broader metabolic effects. This was especially evident for urate transporters, such as *SLC2A9 rs13129697-T*, which was significantly positively associated only with circulating urate concentration but no other metabolic measures. Thus, lifestyle factors that predispose to both gout and compromised cardiovascular health likely play a central role in explaining population-level associations between gout and circulating lipids and glucose.

The metabolic effects of some of the gout risk alleles are also affected by lifestyle factors. For example, *alcohol dehydrogenase 1B* (*ADH1B*) plays a major role in ethanol catabolism (oxidizing ethanol into acetaldehyde)^44^ and affects the ratio of urate production and urate reabsorption during alcohol consumption^5^, potentially leading to gout in carriers of rs1229984-T. This is supported by Sandoval-Plata et al.’s finding that the *ADH1B* variant is found especially among gout patients who consume a lot of alcohol^5^. Other examples include *MLXIPL*, which is known to be induced in response to a high carbohydrate diet^45^, and *CYP1A2* the enzymatic activity of which is affected by smoking, medication, and some dietary constituents^46^. At the population level, the urate concentration-raising effects of the *ADH1B rs1229984-T* and *CYP1A2 rs11857376-A* alleles are possibly diluted because the population includes people with the variant who do not consume alcohol or smoke. Similarly, *MLXIPL rs12531645-G* metabolic associations may be weaker in individuals with a lower dietary carbohydrate intake. The alleles showing strong metabolic associations, such as *GCKR rs1260326-T* and *MLXIPL rs12531645-G*, have previously been associated with abnormal blood lipid and glucose levels^29,47,48^. These observations, combined with the observed genetic correlations with several measures of lipid metabolism, suggest a shared genetic basis between gout and abnormalities in blood lipid and glucose levels. Variations in both genes, *GCKR* and *MLXIPL*, have been found to increase the glycolytic flux, which can lead to increased urate production via the pentose-phosphate pathway and simultaneously reduce fatty acid oxidation and increase lipogenesis^47,49^.

Information obtained from genetic and large-scale metabolomic data can be used to identify potential or poorly suitable medical targets^50^. A good example of this is *ALDH16A1*: the gout risk-increasing allele *ALDH16A1 rs752383928-G* shows negative effects on circulating cholesterol and triglyceride levels, such as LDL cholesterol and total triglycerides, the key cardiovascular risk markers. This makes it an unsuitable target molecule for treating gout, as the lower risk of gout would be achieved at the cost of a higher risk of cardiovascular diseases. The same applies to the *CYP1A2 rs11857376-A* allele we observed in this study. This information can be very valuable in the pre-clinical phase and can help to predict whether it would be worth moving forward with large-scale trials^50–52^.

### Limitations

Our study has limitations. Even though potential genes have been systematically identified, there is little clarity regarding their causality. The relative prevalence of gout cases varied among the sample populations examined in the meta-analysis, indicating that there are likely discrepancies in how different biobanks can identify gout patients. Considering that our sample consists only of individuals of European ancestry, it would be of high value to replicate the findings in other ethnicities. Also, the novel gout locus observed in the sex-stratified GWAS analysis would benefit from replication in other populations as well.

## Conclusions

The novel gout risk loci we discovered add to the knowledge of the hereditary factors behind gout, and the metabolic profiling of the gout risk alleles aids in understanding the molecular pathways underlying gout. Considering the heterogeneous and overall weak metabolic effects of the gout risk alleles, our findings suggest that the population-level associations between gout and circulating lipids and glucose likely arise due to shared lifestyle risk factors and that circulating metabolic measures other than urate appear to lack relevance in the context of gout development.

## Data Availability

The individual-level data are available under restricted access for legal and ethical reasons. Formal approval for the researchers is required to access the data: please see https://www.finngen.fi/en/access_results for more details. Access to individual-level data and genotype data is managed by the Finnish Biobank Cooperative at the Fingenious portal [https://site.fingenious.fi/en/]). The metabolomics GWAS summary statistics used in this study have been made publicly available through the NHGRI-EBI GWAS catalogue (GCST90301941-GCST90302173) and https://www.phpc.cam.ac.uk/ceu/lipids-and-metabolites/. Summary statistics of this study will be made available through the NHGRI-EBI GWAS catalogue upon publication.

## Acknowledgements

E.S. was funded by Academy of Finland (grant number: 338229) and Orion Research Foundation sr. J.K. was funded by Sigrid Juselius foundation. The authors wish to acknowledge CSC – IT Center for Science, Finland, for computational resources. We also want to acknowledge the participants and investigators of FinnGen study. The FinnGen project is funded by two grants from Business Finland (HUS 4685/31/2016 and UH 4386/31/2016) and the following industry partners: AbbVie Inc., AstraZeneca UK Ltd, Biogen MA Inc., Bristol Myers Squibb (and Celgene Corporation & Celgene International II Sàrl), Genentech Inc., Merck Sharp & Dohme LCC, Pfizer Inc., GlaxoSmithKline Intellectual Property Development Ltd., Sanofi US Services Inc., Maze Therapeutics Inc., Janssen Biotech Inc, Novartis Pharma AG, and Boehringer Ingelheim International GmbH. Following biobanks are acknowledged for delivering biobank samples to FinnGen: Auria Biobank (www.auria.fi/biopankki), THL Biobank (www.thl.fi/biobank), Helsinki Biobank (www.helsinginbiopankki.fi), Biobank Borealis of Northern Finland (https://www.ppshp.fi/Tutkimus-ja-opetus/Biopankki/Pages/Biobank-Borealis-briefly-in-English.aspx), Finnish Clinical Biobank Tampere (www.tays.fi/en-US/Research_and_development/Finnish_Clinical_Biobank_Tampere), Biobank of Eastern Finland (www.ita-suomenbiopankki.fi/en), Central Finland Biobank (www.ksshp.fi/fi-FI/Potilaalle/Biopankki), Finnish Red Cross Blood Service Biobank (www.veripalvelu.fi/verenluovutus/biopankkitoiminta), Terveystalo Biobank (www.terveystalo.com/fi/Yritystietoa/Terveystalo-Biopankki/Biopankki/) and Arctic Biobank (https://www.oulu.fi/en/university/faculties-and-units/faculty-medicine/northern-finland-birth-cohorts-and-arctic-biobank). All Finnish Biobanks are members of BBMRI.fi infrastructure (www.bbmri.fi). Finnish Biobank Cooperative -FINBB (https://finbb.fi/) is the coordinator of BBMRI-ERIC operations in Finland. The Finnish biobank data can be accessed through the Fingenious^®^ services (https://site.fingenious.fi/en/) managed by FINBB.

## Notes

### Competing Interest Statement

The authors have declared no competing interest.

### Author Declarations

Patients and control subjects in FinnGen provided informed consent for biobank research, based on the Finnish Biobank Act. Alternatively, separate research cohorts, collected prior the Finnish Biobank Act came into effect (in September 2013) and start of FinnGen (August 2017), were collected based on study-specific consents and later transferred to the Finnish biobanks after approval by Fimea (Finnish Medicines Agency), the National Supervisory Authority for Welfare and Health. Recruitment protocols followed the biobank protocols approved by Fimea. The Coordinating Ethics Committee of the Hospital District of Helsinki and Uusimaa (HUS) statement number for the FinnGen study is Nr HUS/990/2017. The FinnGen study is approved by Finnish Institute for Health and Welfare (permit numbers: THL/2031/6.02.00/2017, THL/1101/5.05.00/2017, THL/341/6.02.00/2018, THL/2222/6.02.00/2018, THL/283/6.02.00/2019, THL/1721/5.05.00/2019 and THL/1524/5.05.00/2020), Digital and population data service agency (permit numbers: VRK43431/2017-3, VRK/6909/2018-3, VRK/4415/2019-3), the Social Insurance Institution (permit numbers: KELA 58/522/2017, KELA 131/522/2018, KELA 70/522/2019, KELA 98/522/2019, KELA 134/522/2019, KELA 138/522/2019, KELA 2/522/2020, KELA 16/522/2020), Findata permit numbers THL/2364/14.02/2020, THL/4055/14.06.00/2020,THL/3433/14.06.00/2020, THL/4432/14.06/2020, THL/5189/14.06/2020, THL/5894/14.06.00/2020, THL/6619/14.06.00/2020, THL/209/14.06.00/2021, THL/688/14.06.00/2021, THL/1284/14.06.00/2021, THL/1965/14.06.00/2021, THL/5546/14.02.00/2020, THL/2658/14.06.00/2021, THL/4235/14.06.00/202, Statistics Finland (permit numbers: TK-53-1041-17 and TK/143/07.03.00/2020 (earlier TK-53-90-20) TK/1735/07.03.00/2021, TK/3112/07.03.00/2021) and Finnish Registry for Kidney Diseases permission/extract from the meeting minutes on 4th July 2019.

