## Supplementary Materials for "Exploring metabolic changes in gout – Insights from a genetic study"

**Supplementary Note**. FinnGen DF9 Ethics statement
**Figure S1.** Manhattan plot from age-stratified GWAS
**Figure S2.** Manhattan plot from sex-stratified GWAS
**Figure S3.** Beta estimates of lead variants in every dataset used in the study
**Figure S4.1.** Locus 16q21 (*CDH8*)
**Figure S4.2.** Locus 17p13.2 (*SLC25A11*)
**Figure S5.** Differences in effect sizes observed in sex and age-stratified GWASs
**Figure S6.** Conditional analysis of the gout association near ALDH16A1
**Figure S7.** Results of FUMA gene-set and tissue expression analysis
**Figure S8.** eQTL colocalization plots

**Table S1.1.** Study populations
**Table S2.1.** Genome-wide significant ($p<5x10^{-8}$) lead variants associated with gout in under 50-year-olds
**Table S2.2.** Genome-wide significant ($p<5x10^{-8}$) lead variants associated with gout in 50-year-olds and older
**Table S2.3.** Genome-wide significant ($p<5x10^{-8}$) lead variants associated with gout in under males
**Table S2.4.** Genome-wide significant ($p<5x10^{-8}$) lead variants associated with gout in females
**Table S2.5.** Genome-wide significant ($p<5x10^{-8}$) lead variants associated with gout in FinnGen
**Table S3.** Unit of measurement for every metabolite analyzed in the study
**Table S4.** Lead variants of the gout associated loci ($P<5x10^{-8}$) identified in the genome-wide meta-analysis
**Table S5.** Calculation of effect differences between sexes and age groups
**Table S6.** eQTL colocalization results
**Table S7.** Genetic correlations
**Table S8.** List of LD-based proxies used in metabolic association profiles
**Table S9.** A list of FinnGen authors and their affiliations
**Table S10.** A list Estonian Biobank Research Team authors and their affiliations

**References**

**Supplementary note.** FinnGen DF9 Ethics statement

Patients and control subjects in FinnGen provided informed consent for biobank research, based on the Finnish Biobank Act. Alternatively, separate research cohorts, collected prior the Finnish Biobank Act came into effect (in September 2013) and start of FinnGen (August 2017), were collected based on study-specific consents and later transferred to the Finnish biobanks after approval by Fimea (Finnish Medicines Agency), the National Supervisory Authority for Welfare and Health. Recruitment protocols followed the biobank protocols approved by Fimea. The Coordinating Ethics Committee of the Hospital District of Helsinki and Uusimaa (HUS) statement number for the FinnGen study is Nr HUS/990/2017.

The FinnGen study is approved by Finnish Institute for Health and Welfare (permit numbers: THL/2031/6.02.00/2017, THL/1101/5.05.00/2017, THL/341/6.02.00/2018, THL/2222/6.02.00/2018, THL/283/6.02.00/2019, THL/1721/5.05.00/2019 and THL/1524/5.05.00/2020), Digital and population data service agency (permit numbers: VRK43431/2017-3, VRK/6909/2018-3, VRK/4415/2019-3), the Social Insurance Institution (permit numbers: KELA 58/522/2017, KELA 131/522/2018, KELA 70/522/2019, KELA 98/522/2019, KELA 134/522/2019, KELA 138/522/2019, KELA 2/522/2020, KELA 16/522/2020), Findata permit numbers THL/2364/14.02/2020, THL/4055/14.06.00/2020,,THL/3433/14.06.00/2020, THL/4432/14.06/2020, THL/5189/14.06/2020, THL/5894/14.06.00/2020, THL/6619/14.06.00/2020, THL/209/14.06.00/2021, THL/688/14.06.00/2021, THL/1284/14.06.00/2021, THL/1965/14.06.00/2021, THL/5546/14.02.00/2020, THL/2658/14.06.00/2021, THL/4235/14.06.00/202, Statistics Finland (permit numbers: TK-53-1041-17 and TK/143/07.03.00/2020 (earlier TK-53-90-20) TK/1735/07.03.00/2021, TK/3112/07.03.00/2021) and Finnish Registry for Kidney Diseases permission/extract from the meeting minutes on 4$^{th}$ July 2019.

The Biobank Access Decisions for FinnGen samples and data utilized in FinnGen Data Freeze 9 include: THL Biobank BB2017_55, BB2017_111, BB2018_19, BB_2018_34, BB_2018_67, BB2018_71, BB2019_7, BB2019_8, BB2019_26, BB2020_1, Finnish Red Cross Blood Service Biobank 7.12.2017, Helsinki Biobank HUS/359/2017, HUS/248/2020, Auria Biobank AB17-5154 and amendment #1 (August 17 2020), AB20-5926 and amendment #1 (April 23 2020) and it´s modification (Sep 22 2021), Biobank Borealis of Northern Finland_2017_1013, Biobank of Eastern Finland 1186/2018 and amendment 22 § /2020, Finnish Clinical Biobank Tampere MH0004 and amendments (21.02.2020 & 06.10.2020), Central Finland Biobank 1-2017, and Terveystalo Biobank STB 2018001 and amendment 25$^{th}$ Aug 2020.

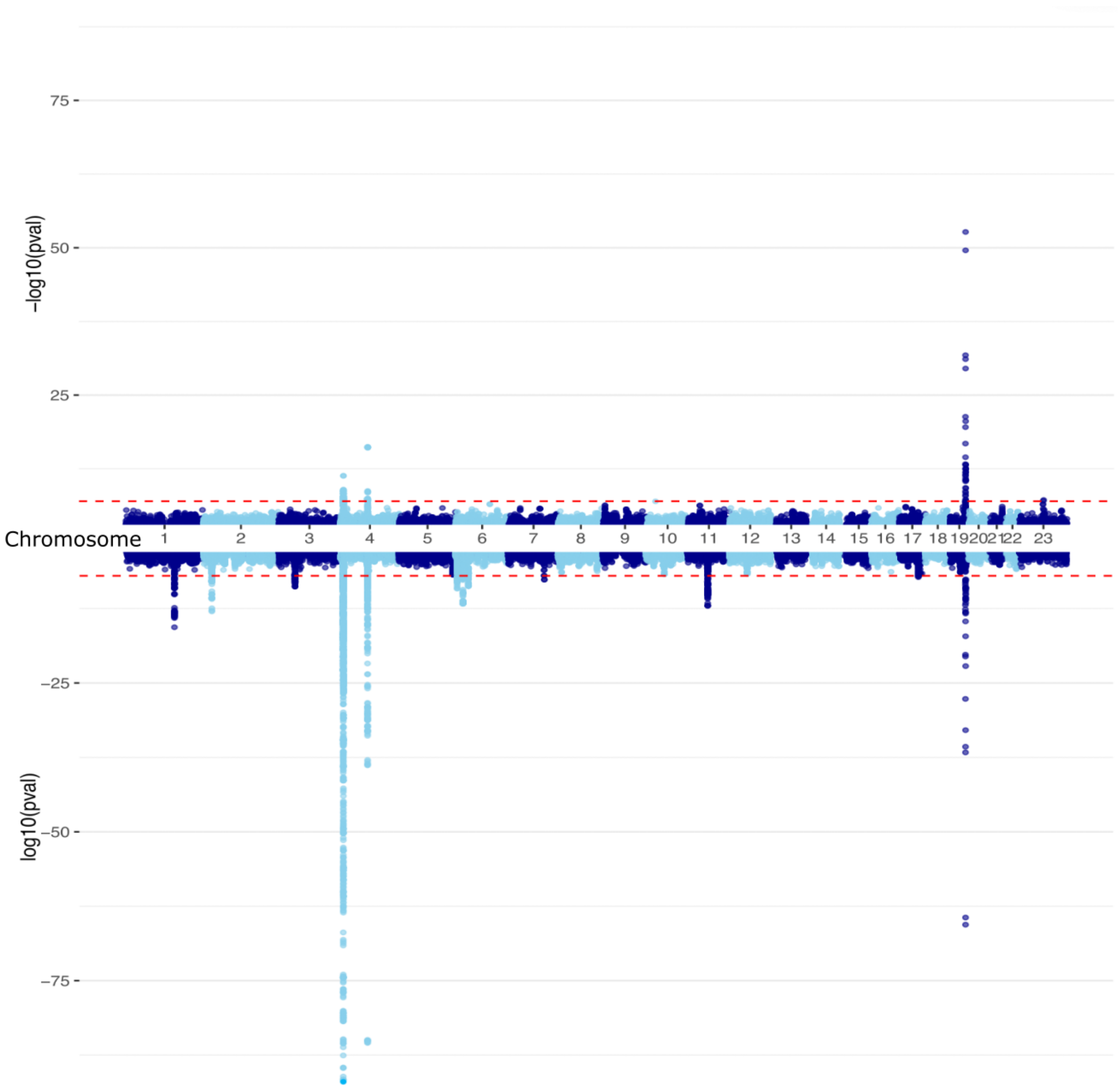


**Figure S1.** Manhattan plots from age-stratified GWASs. The GWAS conducted in individuals diagnosed before 50 years of age (top) included 988 cases and 366 392 controls. The GWAS conducted in individuals diagnosed 50 years of age or older (bottom) included 9897 cases and 366 292 controls. Both GWASs are based on FinnGen data. Case definitions are based on ICD-10 codes: M10.0 & M10.9 and ICD-9 codes 274.0 and 274.9. Both GWASs were adjusted for age, sex, and the first 10 genetic principal components.

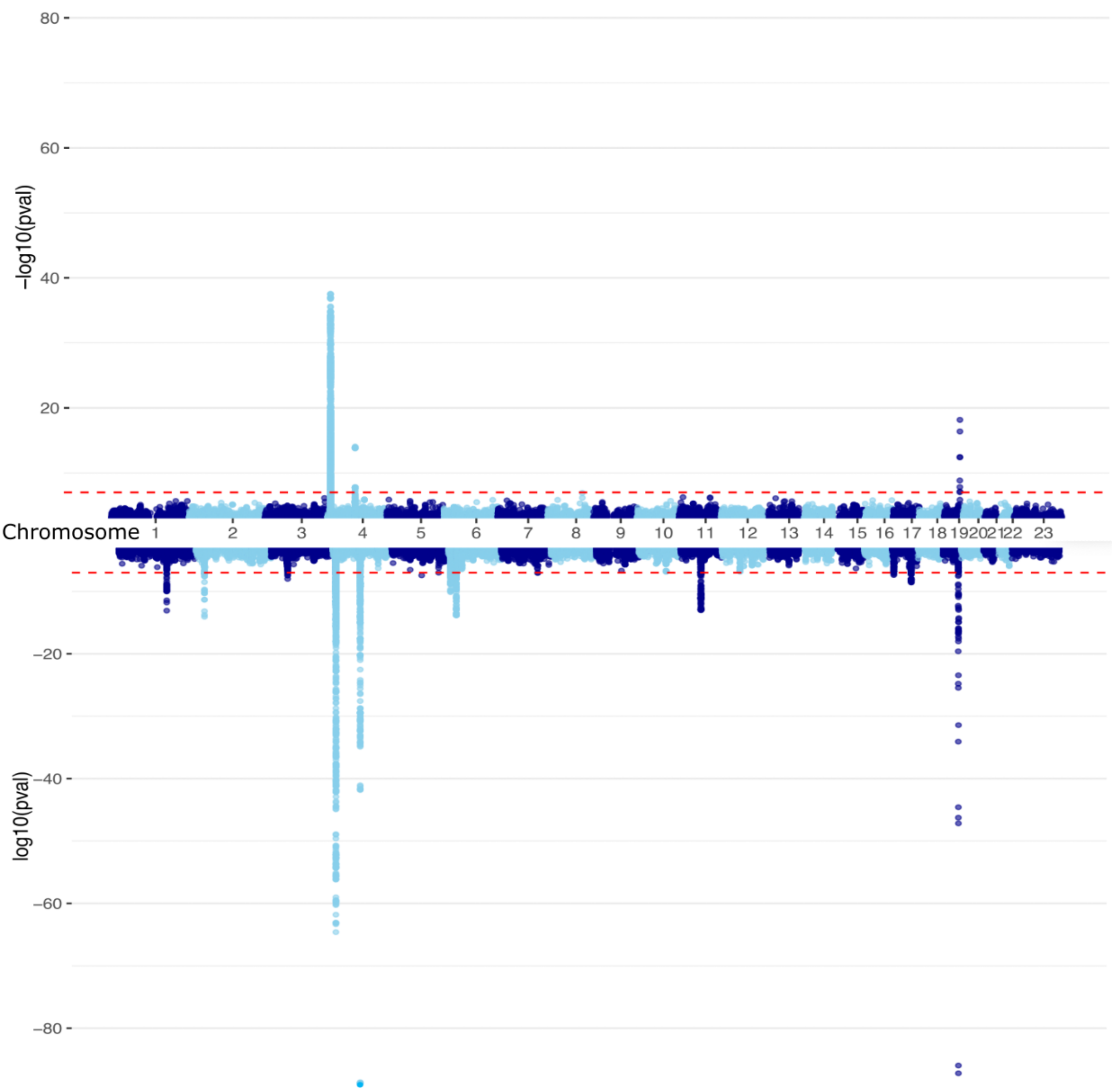


**Figure S2.** Manhattan plots from sex-stratified GWASs. The GWAS conducted in females (top) included 2743 cases and 208 127 female controls. The GWAS conducted in males (bottom) included 8142 cases and 158 265 controls. Both GWASs are based on FinnGen data. Case definitions are based on ICD-10 codes: M10.0 & M10.9 and ICD-9 codes 274.0 & 274.9. Both GWASs were adjusted for age and the first 10 genetic principal components.

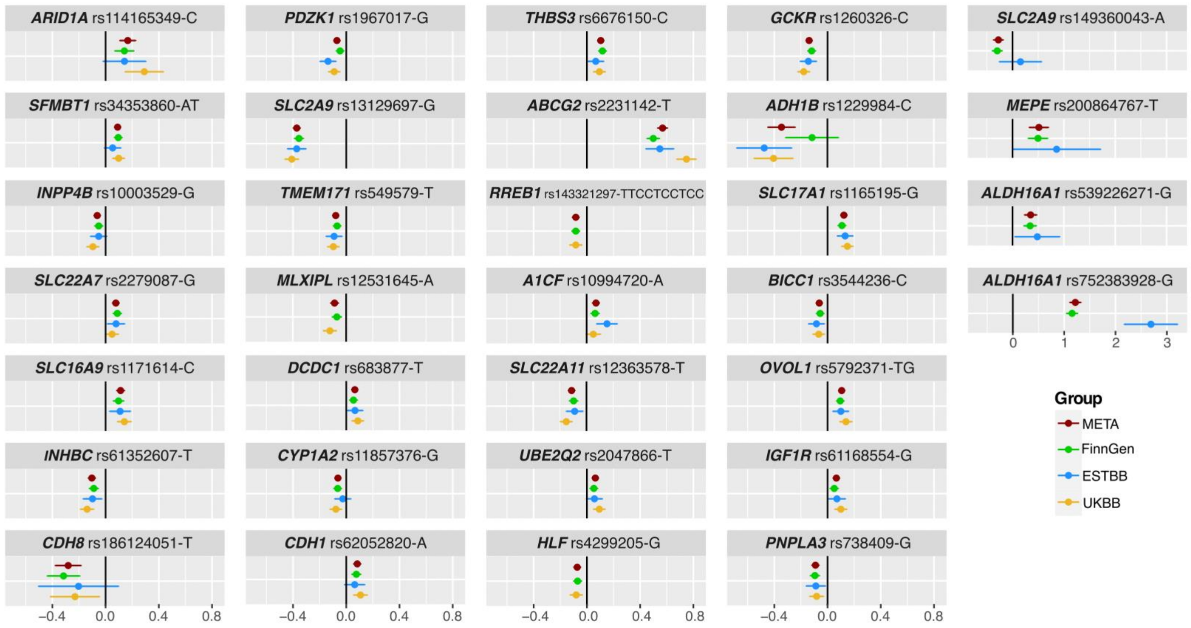


**Figure S3.** Forest plot of effect estimates of lead variants and their 95% confidence intervals. Variants are identified by candidate gene, rsid, and effect allele. The plot shows the effect estimates for each variant both in the meta-analysis (red) and in each individual dataset.

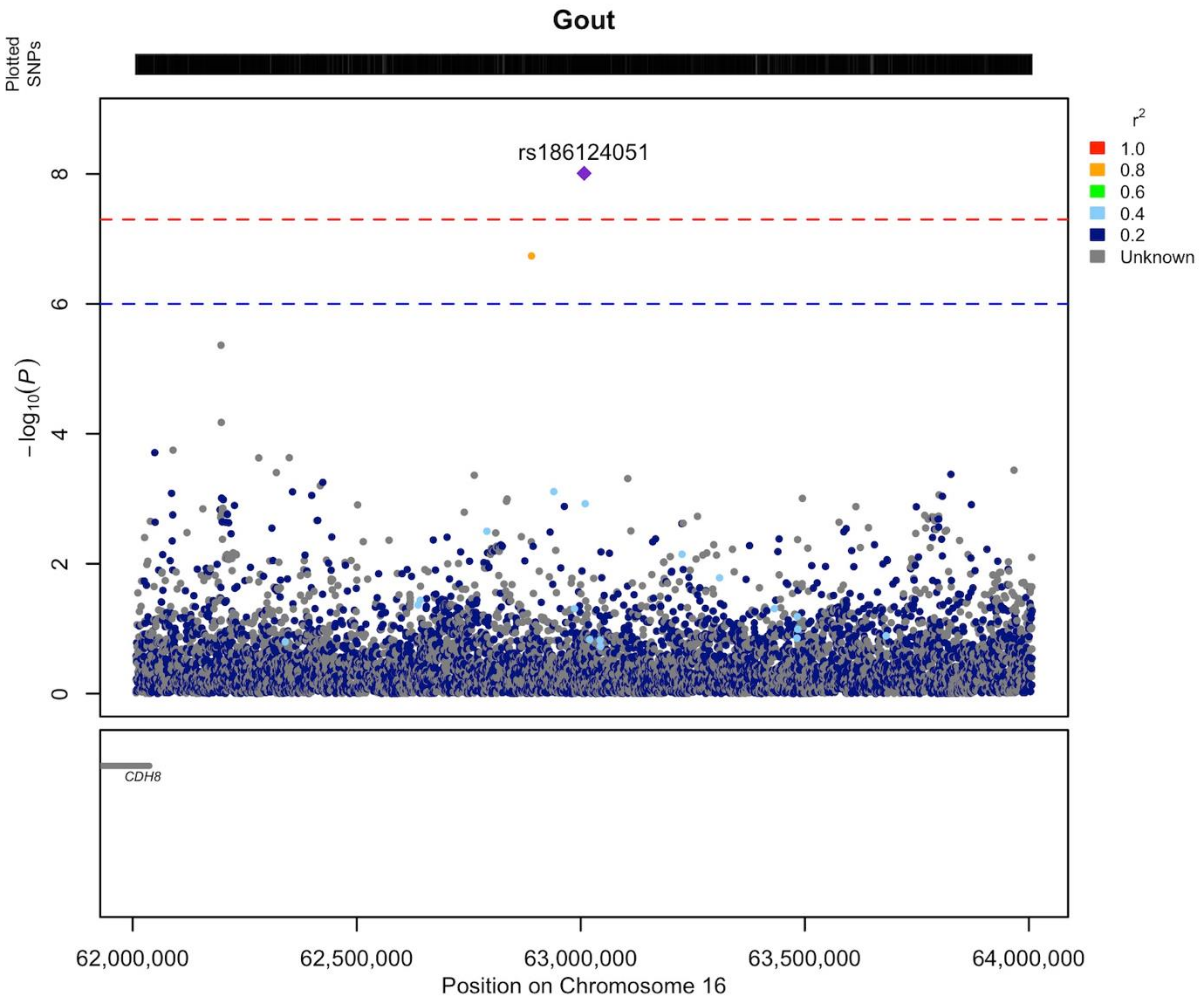


**Figure S4.1.** Regional association plot of novel gout association on chromosome 16 region 62.00-64.00Mb. The association is based on the results of a meta-analysis. The gene explaining the association of the locus is likely *CDH8* (*cadherin 8*). The LD information for the locus was calculated in FinnGen, and the plot was created using the R package Locuszooms (https://github.com/Geeketics/LocusZooms). Ensembl archive, version https://jul2023.archive.ensembl.org, was utilised to create the gene lists.

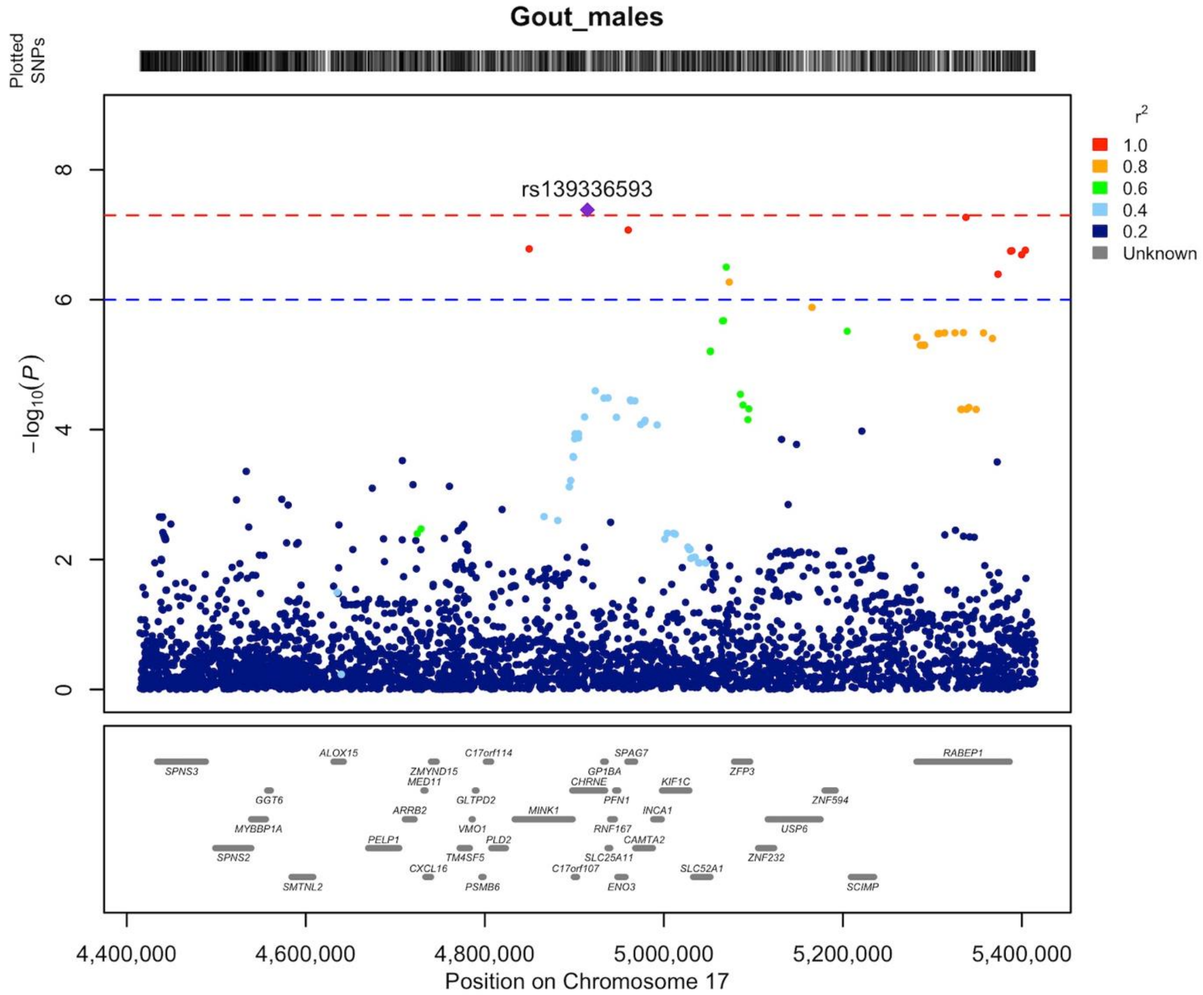


**Figure S4.2.** Regional association plot of novel gout association on chromosome 17 region 4.41-5.41Mb. Locus 17p13.2 was associated with gout in the male subgroup in FinnGen. The gene explaining the association of the locus is likely *SLC25A11* (*solute carrier family member 11*). The LD information for the locus was calculated in FinnGen, and the plot was created using the R package Locuszooms (https://github.com/Geeketics/LocusZooms). Ensembl archive, version https://jul2023.archive.ensembl.org, was utilised to create the gene lists.

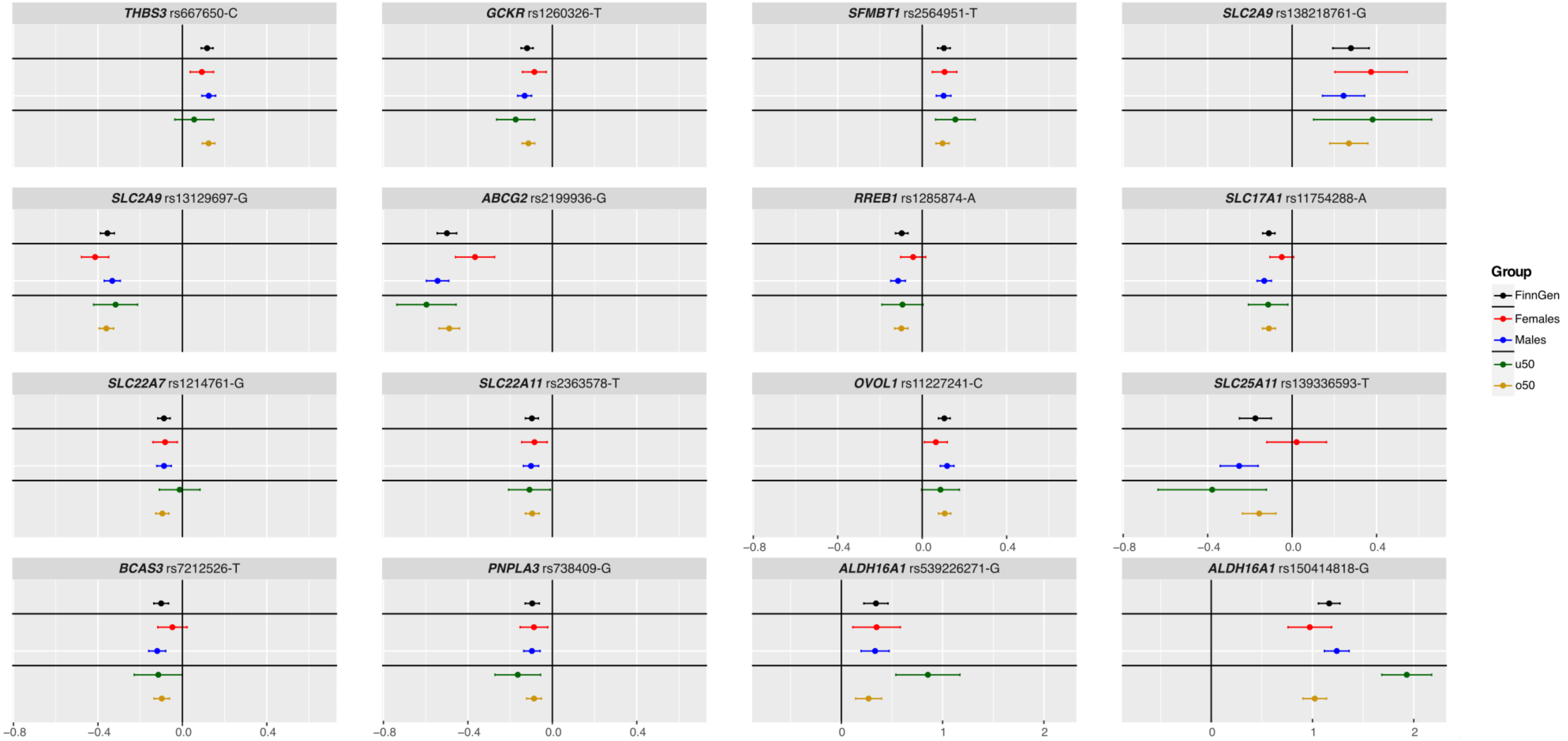


**Figure S5.** Comparison of the effect estimates of the lead variants discovered in the FinnGen and in age- and sex-stratified GWASs. The plot shows effect estimates and the corresponding 95% confidence intervals in each subgroup. Significant differences between the groups of interest (males vs. females, and under 50 years vs. over 50 years) are indicated with an asterisk (numerical representation is available Table S4).

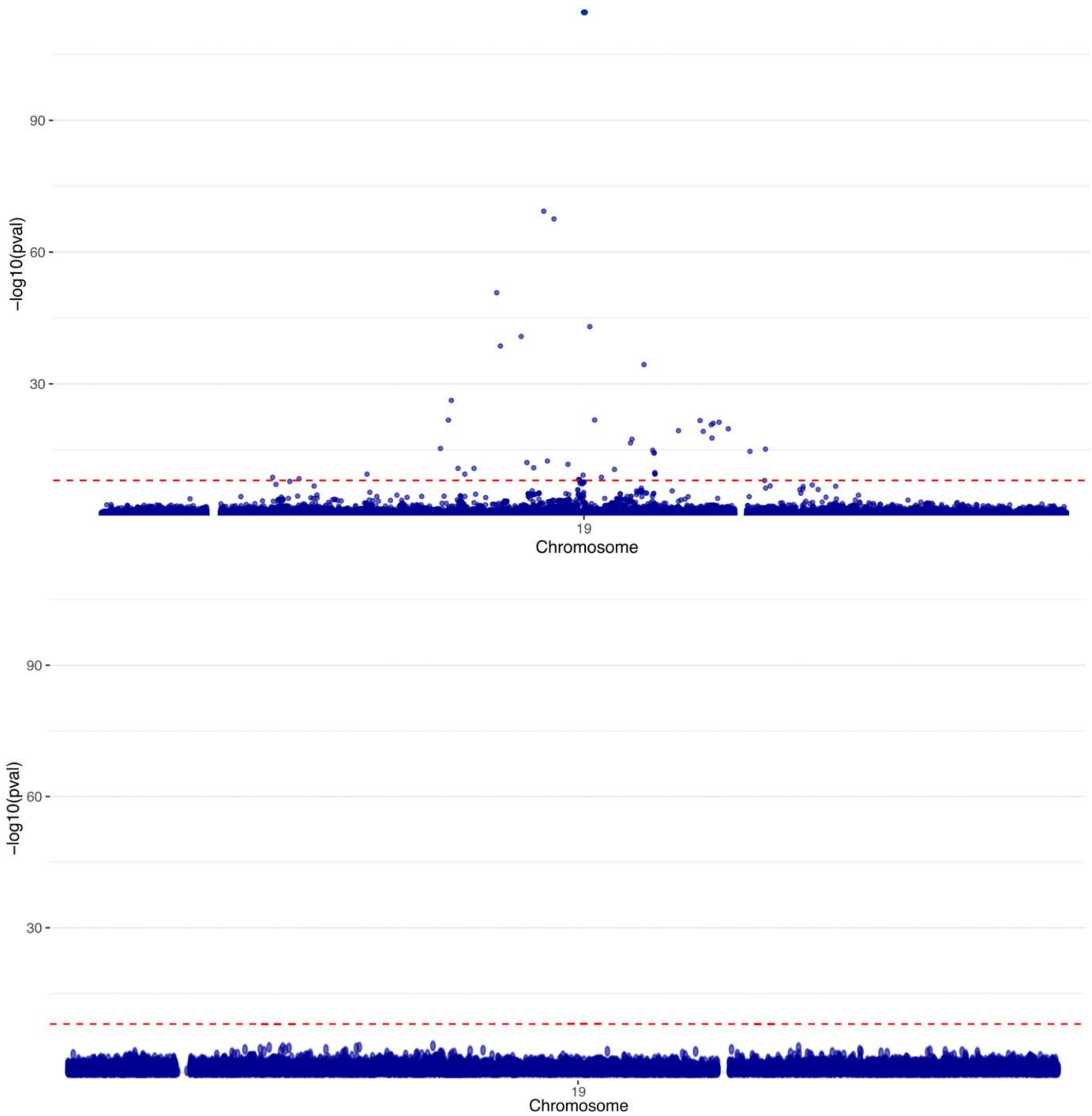


**Figure S6.** Conditional analysis of the gout association near *ALDH16A1*. The plot shows the association test results as obtained from the original GWAS in FinnGen (top) and from the conditional GWAS (bottom) that was adjusted for the rs150414818 (19:49465749:C:G, missense) genotype. The plot shows a ± 4MB window (47465749-51465749) around the rs150414818 variant.

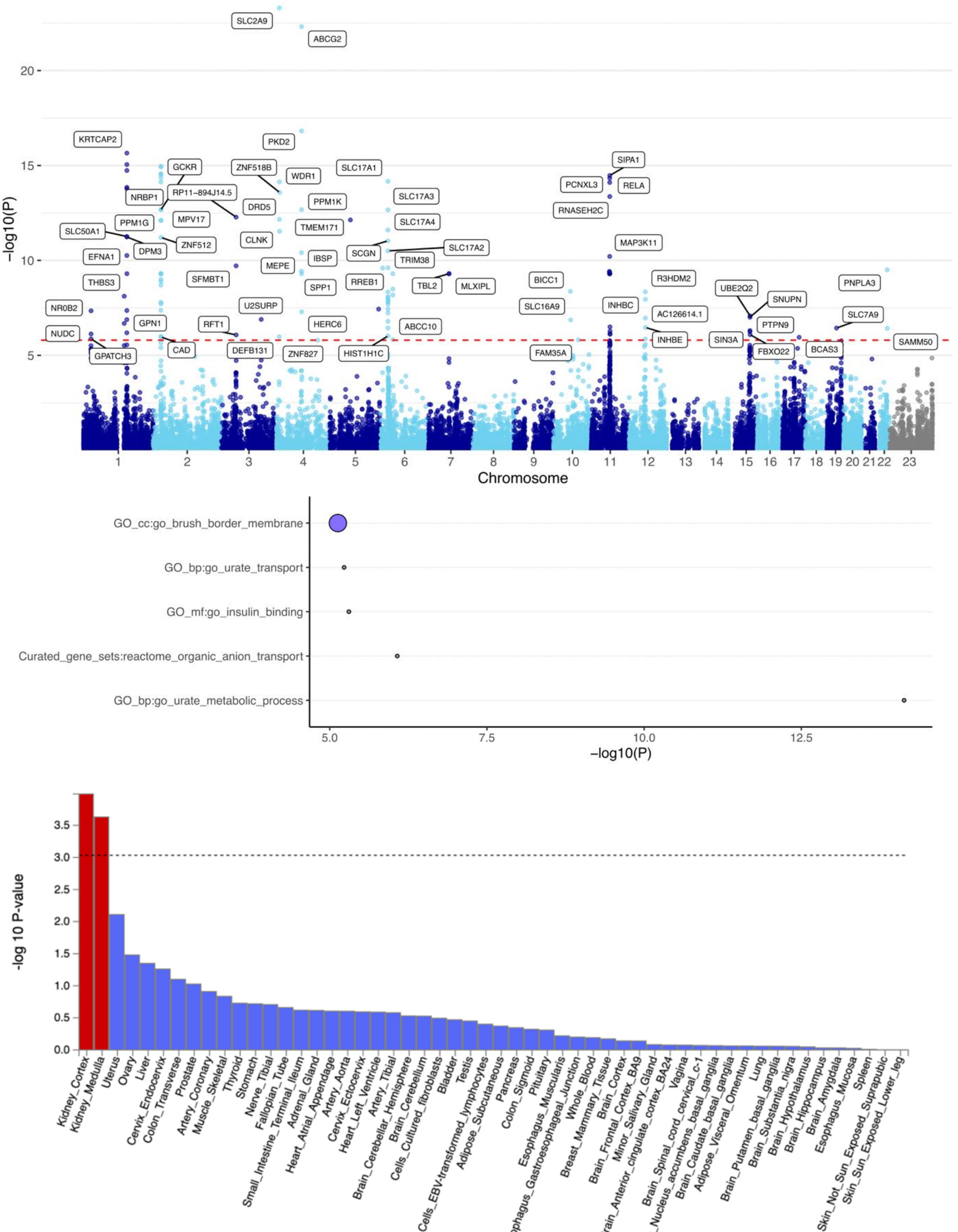


**Figure S7.** Top: Results of the MAGMA[1] gene-based test in a Manhattan plot; X-axis, chromosomes; y-axis, -log(p-value). Middle: MAGMA gene-set enrichment analysis; The plot shows significantly enriched pathways (pFDR < 0.05), curated gene sets, and GO-annotations ranked by p-value -log10(P). The size of the circles refers to the size of the gene set. Small grey <15 and purple 100-200 genes. Bottom: Results of tissue expression analysis: y-axis, -log 10 P-value; x-axis, tissues (GTEx Output-General Tissues). A dashed line indicates statistical significance above the threshold of $-\log_{10}(3.03)$ ($-\log_{10}(0.05/54)$) after Bonferroni correction for testing 54 tissues. Tissues with significant results are highlighted in red. Analysis was done using FUMA[2], and the gene-sets and GO annotations included in the analysis are from MSigDB[3].

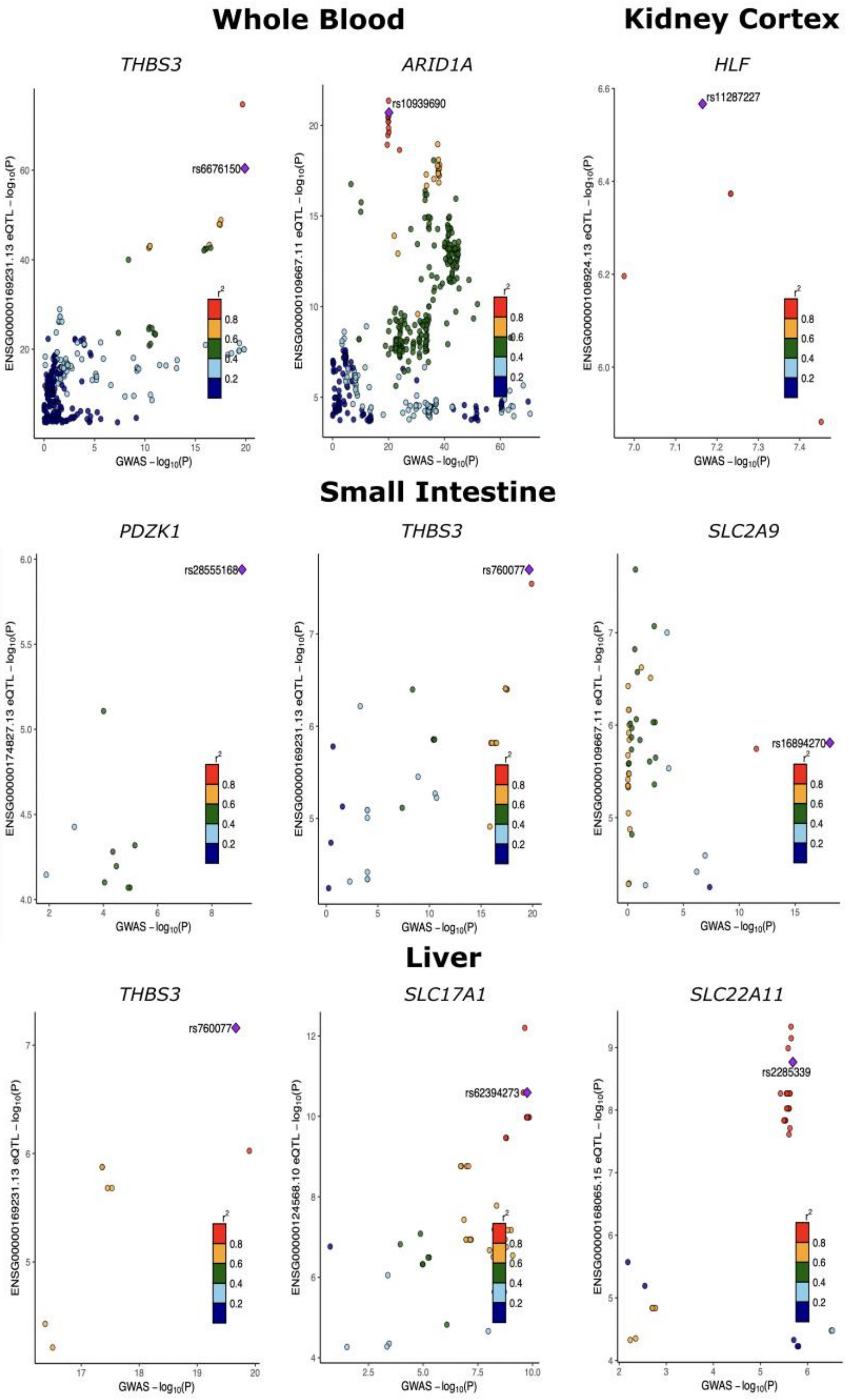


**Figure S8.** eQTL colocalizations between gout associations and gene expression. Colocalizations were investigated for a total of 39 genes. Candidate genes and genes closest to the association signal were selected for the analysis if the closest gene was a different gene than the candidate gene. Approximate Bayesian factor analyses were performed with the 'coloc.abf' function found in the 'coloc' R-library[4]. We focused on tissues that, based on our results and previous studies, are relevant for gout. Consequently, kidney cortex, liver, small intestine, and whole blood were selected for analysis. Variant-gene expression associations for the analysis were downloaded from GTExportal (https://www.gtexportal.org/home/datasets). Colocalizations with posterior probabilities ≥ 0,8 for the variant were considered significant[5].

**Table S1.1.** Study populations. The number of gout cases and controls in the study.

| Study population | Cases | Controls | Sample prevalence | Total |
|---|---|---|---|---|
| FinnGen | 10 885 | 366 392 | 2.9% | 377 277 |
| Estonian Biobank | 2 578 | 192 600 | 1.3% | 195 178 |
| UK Biobank | 4 509 | 415 619 | 1,1% | 420 128 |
| **Meta-analysis** | **17 972** | **974661** | **1.8%** | **992 583** |
| FinnGen subgroup analysis | | | | |
| Males | 8 142 | 158 265 | 4.9% | 166 407 |
| Females | 2 743 | 208 127 | 1.3% | 210 870 |
| Under 50-year-olds | 988 | 366 392 | 0.3% | 367 380 |
| Over 50-year-olds | 9 897 | 366 392 | 2.6% | 376 289 |

**Table S2.1.** Genome-wide significant ($p<5x10^{-8}$) lead variants associated with gout in patients diagnosed before 50 years of age. The table reports lead variants of the gout-associated loci (at least 1MB apart) identified in the GWAS of 988 gout cases and 366,392 controls. GWAS includes only gout cases diagnosed before the age of 50. If the patient had multiple diagnoses, only the initial diagnosis of gout was considered in the study.

| Locus | Candidate gene | CHR:POS | rsid | EA | OA | OR (95% CI) | p-value | Ref. |
|---|---|---|---|---|---|---|---|---|
| **4p16.1** | *SLC2A9* | 4:9919558 | rs34426626 | AC | C | 0.73 (0.64-0.82) | 4.89e-12 | [6–9] |
| **4q22.1** | *ABCG2* | 4:88128874 | rs74904971 | A | C | 1.82 (1.68-1.96) | 6.45e-17 | [7–9] |
| **19q13.33** | *ALDH16A1* | 19:49465749 | rs150414818 | G | C | 6.89 (6.65-7.14) | 2.10e-53 | [6] |

Candidate gene, a gene at a new locus the biological function of which is likely to explain the gout association; CHR: POS, chromosome, and position (genome build hg38); rsid, SNP markers identification number; EA, effect allele; OA, other allele; OR (95% CI), odds ratio and it's 95% confidence interval; Ref, reference article in which a gout association was observed +/- 1 Mb in the vicinity of the lead variant.

**Table S2.2.** Genome-wide significant ($p<5x10^{-8}$) lead variants associated with gout in patients diagnosed after 50 years of age. The table reports lead variants of the gout-associated loci (at least 1MB apart) identified in the GWAS of 9897 gout cases and 366,392 controls. GWAS includes only gout cases diagnosed after the age of 50. If the patient had multiple diagnoses, only the initial diagnosis of gout was considered in the study.

| Locus | Candidate gene | CHR:POS | rsid | EA | OA | OR (95% CI) | p-value | Ref. |
|---|---|---|---|---|---|---|---|---|
| **1q22** | *THBS3* | 1:155151361 | rs6676150 | C | G | 1.13 (1.10-1.16) | 2.29e-16 | [8,9] |
| **2p23.3** | *GCKR* | 2:27518370 | rs780094 | C | T | 0.89 (0.86-0.92) | 1.03e-13 | [6–9] |
| **3p21.1** | *SFMBT1* | 3:52944675 | rs2710332 | G | A | 1.10 (1.07-1.13) | 1.47e-09 | [7–9] |
| **4p16.1** | *SLC2A9* | 4:8827046 | rs138218761 | G | A | 1.31 (1.22-1.40) | 5.29e-09 | [6–9] |
| **4p16.1** | *SLC2A9* | 4:9925343 | rs13129697 | G | T | 0.70 (0.66-0.73) | 1.04e-92 | [6–9] |
| **4q22.1** | *ABCG2* | 4:88131171 | rs2231142 | T | G | 1.63 (1.58-1.68) | 3.66e-86 | [7–9] |
| **6p24.3** | *RREB1* | 6:7115300 | rs1285874 | A | T | 0.91 (0.87-0.94) | 7.28e-10 | [6–9] |
| **6p22.2** | *SLC17A1* | 6:25801091 | rs1165209 | A | G | 1.11 (1.08-1.14) | 2.02e-12 | [6–9] |
| **6p21.1** | *SLC22A7* | 6:43386693 | rs1214761 | G | A | 0.91 (0.88-0.94) | 1.82e-09 | [9] |
| **11q13.1** | *SLC22A11* | 11:64597394 | rs12363578 | T | C | 0.91 (0.88-0.94) | 1.03e-08 | [6–9] |
| **11q13.1** | *OVOL1* | 11:65622419 | rs11227241 | C | G | 1.11 (1.08-1.14) | 8.67e-13 | [8] |
| **19q13.33** | *ALDH16A1* | 19:49465749 | rs150414818 | G | C | 2.78 (2.66-2.90) | 2.53e-66 | [6] |

Candidate gene, a gene at a new locus the biological function of which is likely to explain the gout association; CHR: POS, chromosome and position (genome build hg38); rsid, SNP markers identification number; EA, effect allele; OA, other allele; OR (95% CI), odds ratio and it's 95% confidence interval; Ref, reference article in which a gout association was observed +/- 1 Mb in the vicinity of the lead variant.

**Table S2.3.** Genome-wide significant ($p<5x10^{-8}$) lead variants associated with gout in the male subgroup. The table reports lead variants of the gout-associated loci (at least 1MB apart) identified in the GWAS of 8142 gout cases and 158,265 same-sex controls. GWAS compares males to same-sex controls.

| Locus | Candidate gene | CHR:POS | rsid | EA | OA | OR (95% CI) | p-value | Ref. |
|---|---|---|---|---|---|---|---|---|
| **1q22** | *THBS3* | 1:155151361 | rs6676150 | C | G | 1.13 (1.10-1.17) | 8.20e-14 | [8,9] |
| **2p23.3** | *GCKR* | 2:27508073 | rs1260326 | C | T | 0.88 (0.84-0.91) | 8.46e-15 | [6–9] |
| **3p21.1** | *SFMBT1* | 3:53026433 | rs2564951 | T | C | 1.11 (1.07-1.14) | 1.00e-08 | [7–9] |
| **4p16.1** | *SLC2A9* | 4:9925343 | rs13129697 | G | T | 0.72 (0.68-0.76) | 2.49e-65 | [6–9] |
| **4q22.1** | *ABCG2* | 4:88131171 | rs2231142 | T | G | 1.72 (1.67-1.78) | 9.28e-90 | [7–9] |
| **6p24.3** | *RREB1* | 6:7109432 | rs3904600 | G | C | 0.90 (0.87-0.93) | 7.42e-11 | [6–9] |
| **6p22.2** | *SLC17A1* | 6:25817561 | rs1165153 | G | A | 1.14 (1.11-1.17) | 1.40e-14 | [6–9] |
| **11q13.1** | *SLC22A11* | 11:64597394 | rs12363578 | T | C | 0.90 (0.87-0.94) | 3.93e-08 | [6–9] |
| **11q13.1** | *OVOL1* | 11:65744586 | rs7110576 | G | T | 1.13 (1.10-1.16) | 1.00e-13 | [8] |
| **17p13.2** | ***SLC25A11*** | **17:4914555** | **rs139336593** | **T** | **G** | **0.78 (0.69-0.87)** | **4.14e-08** | **Novel** |
| **17q23.2** | *BCAS3* | 17:61170034 | rs11658810 | C | G | 0.89 (0.85-0.93) | 2.75e-09 | [10] |
| **19q13.33** | *ALDH16A1* | 19:49465749 | rs150414818 | G | C | 3.45 (3.33-3.58) | 5.90e-88 | [6] |

Candidate gene, a gene at a new locus the biological function of which is likely to explain the gout association; CHR: POS, chromosome and position (genome build hg38); rsid, SNP markers identification number; EA, effect allele; OA, other allele; OR (95% CI), odds ratio and it's 95% confidence interval; Ref, reference article in which a gout association was observed +/- 1 Mb in the vicinity of the lead variant.

**Table S2.4.** Genome-wide significant ($p<5x10^{-8}$) lead variants associated with gout in the female subgroup. The table reports lead variants of the gout-associated loci (at least 1MB apart) identified in the GWAS of 2743 gout cases and 208,127 same-sex controls. GWAS compares females to same-sex controls.

| Locus | Candidate gene | CHR:POS | rsid | EA | OA | OR (95% CI) | p-value | Ref. |
|---|---|---|---|---|---|---|---|---|
| **4p16.1** | *SLC2A9* | 4:9982917 | rs9994216 | T | G | 1.57 (1.50-1.64) | 2.80e-38 | [6–9] |
| **4q22.1** | *ABCG2* | 4:88124179 | rs2199936 | G | A | 0.69 (0.60-0.79) | 8.83e-15 | [7–9] |
| **19q13.33** | *ALDH16A1* | 19:49465749 | rs150414818 | G | C | 2.65 (2.43-2.86) | 6.94e-19 | [6] |

Candidate gene, a gene at a new locus the biological function of which is likely to explain the gout association; CHR: POS, chromosome and position (genome build hg38); rsid, SNP markers identification number; EA, effect allele; OA, other allele; OR (95% CI), odds ratio and it's 95% confidence interval; Ref, reference article in which a gout association was observed +/- 1 Mb in the vicinity of the lead variant.

**Table S2.5.** Genome-wide significant ($p<5x10^{-8}$) lead variants associated with gout in FinnGen. The table reports lead variants of the gout-associated loci (at least 1MB apart) identified in the GWAS of 10,885 gout cases and 366,392 controls.

| Locus | Candidate Gene | CHR:POS | rsid | EA | OA | OR (95% CI) | p-value | Ref. |
|---|---|---|---|---|---|---|---|---|
| **1q22** | *THBS3* | 1:155151361 | rs6676150 | C | G | 1.12 (1.10-1.15) | 3.99e-16 | 8,9 |
| **2p23.3** | *GCKR* | 2:27508073 | rs1260326 | C | T | 0.89 (0.86-0.92) | 2.45e-16 | 6–9 |
| **3p21.1** | *SFMBT1* | 3:53026433 | rs2564951 | T | C | 1.11 (1.08-1.14) | 1.32e-11 | 7–9 |
| **4p16.1** | *SLC2A9* | 4:8827046 | rs138218761 | G | A | 1.32 (1.24-1.41) | 2.14e-10 | 6–9 |
| **4p16.1** | *SLC2A9* | 4:9925343 | rs13129697 | G | T | 0.70 (0.67-0.73) | 1.12e-99 | 6–9 |
| **4q22.1** | *ABCG2* | 4:88124179 | rs2199936 | G | A | 0.61 (0.56–0.65) | 3.59e-100 | 7–9 |
| **6p24.3** | *RREB1* | 6:7115300 | rs1285874 | A | T | 0.91 (0.88-0.94) | 2.04e-10 | 6–9 |
| **6p22.2** | *SLC17A1* | 6:25776721 | rs11754288 | A | G | 0.90 (0.87-0.92) | 1.14e-13 | 6–9 |
| **6p21.1** | *SLC22A7* | 6:43386693 | rs1214761 | G | A | 0.92 (0.89-0.95) | 6.40e-09 | 9 |
| **11q13.1** | *SLC22A11* | 11:64597394 | rs12363578 | T | C | 0.91 (0.88-0.94) | 8.99e-10 | 6–9 |
| **11q13.1** | *OVOL1* | 11:65622419 | rs11227241 | C | G | 1.11 (1.08-1.14) | 1.48e-13 | 8 |
| **17q23.2** | *BCAS3* | 17:61172493 | rs7212526 | T | C | 0.90 (0.87-0.94) | 1.38e-08 | 10 |
| **19q13.33** | *ALDH16A1* | 19:48175672 | rs539226271 | G | T | 1.41 (1.29-1.53) | 1.93e-08 | 6,11 |
| **19q13.33** | *ALDH16A1* | 19:49465749 | rs150414818 | G | C | 3.20 (3.10-3.31) | 5.89e-103 | 6 |
| **22q13.31** | *PNPLA3* | 22:43928847 | rs738409 | G | C | 0.91 (0.88-0.94) | 2.34e-08 | 7–9 |

Candidate gene, a gene at a new locus the biological function of which is likely to explain the gout association; CHR: POS, chromosome and position (genome build hg38); rsid, SNP markers identification number; EA, effect allele; OA, other allele; OR (95% CI), odds ratio and it's 95% confidence interval; Ref, reference article in which a gout association was observed +/- 1 Mb in the vicinity of the lead variant.

**Table S3.** List of metabolic measures used in the study. The metabolic effects of the gout alleles were extracted from a metabolomics GWAS[12]. Except for C-reactive protein (CRP) (https://gwas.mrcieu.ac.uk/files/ukb-d-30710_irnt/ukb-d-30710_irnt_report.html#diagnostics) and urate (https://gwas.mrcieu.ac.uk/files/ukb-d-30880_irnt/ukb-d-30880_irnt_report.html) which were extracted from the MR-Base website[13,14]. The table lists the subset of 66 metabolic measures and analyzed in the present study and the respective units of measurements.

| Metabolite group | Abbreviation | Metabolite | Unit of measure |
|---|---|---|---|
| **Lipoprotein particles** | | | |
| | XXL-VLVL-P | Concentration of chylomicrons and very large VLDL particles | mmol/l |
| | XL-VLDL-P | Concentration of very large VLDL particles | |
| | L-VLDL-P | Concentration of large VLDL particles | |
| | M-VLDL-P | Concentration of medium VLDL particles | |
| | S-VLDL-P | Concentration of small VLDL particles | |
| | XS-VLDL-P | Concentration of very small VLDL particles | |
| | IDL-P | Concentration of IDL particles | |
| | L-LDL-P | Concentration of large LDL particles | |
| | M-LDL-P | Concentration of medium LDL particles | |
| | S-LDL-P | Concentration of small LDL particles | |
| | XL-HDL-P | Concentration of very large HDL particles | |
| | L-HDL-P | Concentration of large HDL particles | |
| | M-HDL-P | Concentration of medium HDL particles | |
| | S-HDL-P | Concentration of small HDL particles | |
| **Cholesterol** | | | |
| | Serum-C | Serum total cholesterol | mmol/l |
| | VLDL-C | Total cholesterol in VLDL | |
| | IDL-C | Total cholesterol in IDL | |
| | LDL-C | Total cholesterol in LDL | |
| | HDL-C | Total cholesterol in HDL | |
| **Triglycerides** | | | |
| | Serum-TG | Serum total triglycerides | mmol/l |
| | VLDL-TG | Total triglycerides in VLDL | |
| | IDL-TG | Total triglycerides in IDL | |
| | LDL-TG | Total triglycerides in LDL | |
| | HDL-TG | Total triglycerides in HDL | |
| **Lipoprotein particle sizes** | | | |
| | VLDL-D | VLDL diameter | nm |
| | LDL-D | LDL diameter | |
| | HDL-D | HDL diameter | |
| **Apolipoprotein concentrations** | | | |
| | ApoB | Apolipoprotein B | g/l |
| | ApoA1 | Apolipoprotein A1 | |
| | ApoBbyApoA1 | Ratio of Apolipoprotein B to Apolipoprotein A1 | |
| **Fatty acids** | | | |
| | DHA | Docohexaenoic acid | mmol/l |
| | FAw3 | Omega-3 fatty acids | |
| | LA | Linoleic acid | |
| | FAw6 | Omega-6 fatty acids | |
| | MUFA | Mono-unsaturated fatty acids | |
| | SFA | Saturated fatty acids | |
| | TotFA | | |
| **Relative proportions of fatty acids** | | | |
| | DHAbyFA | Ratio of docohexaenoic acid to fatty acids | % |
| | FAw3byFA | Ratio of omega-3 fatty acids to fatty acids | |
| | LAbyFA | Ratio of linoleic acid to fatty acids | |
| | Faw6byFA | Ratio of omega-6 fatty acids to fatty acids | |
| | MUFAbyFA | Ratio of mono-unsaturated fatty acids to fatty acids | |
| | SFAbyFA | Ratio of saturated fatty acids to fatty acids | |
| **Estimated degree of unsaturation of fatty acids** | | | |
| | UnsatDeg | Unsaturation level | % |
| **Fatty acid derivatives** | | | |
| | AcAce | Acetoacetate | mmol/l |
| | Ace | Acetate | |
| | bOHBut | 3-hydroxybutyrate | |
| **Glucose metabolism related measures** | | | |
| | Glu | Glucose | mmol/l |
| | Lac | Lactate | |
| | Pyr | Pyruvate | |
| | Cit | Citrate | |
| | Glol | Glyserol | |
| **Amino acids** | | | |

| | | | |
|---|---|---|---|
| | Ile | Isoleucine | mmol/l |
| | Leu | Leucine | |
| | Val | Valine | |
| | Phe | Phenylalanine | |
| | Tyr | Tyrosine | |
| | His | Histidine | |
| | Ala | Alanine | |
| | Gln | Glutamine | |
| | Gly | Glycine | |
| **Markers that potentially affect kidney function** | | | |
| | Crea | Creatinine | mmol/l & cu |
| | Urate | Urate | |
| | Alb | Albumin | |
| **Inflammatory markers** | | | |
| | GlycA | Glycoprotein acetyls | mmol/l |
| | CRP_ukb | C-reactive protein | |

**Table S4.** Lead variants of the gout-associated loci. The table reports lead variants of the gout associated loci ($P<5x10^{-8}$, at least 1MB apart) identified in the genome-wide meta-analysis of 17,972 gout cases and 974,611 controls from FinnGen, the Estonian Biobank, and the UK Biobank (top) and a novel locus identified in a genome-wide association study of gout in a male subgroup of FinnGen participants (8,142 cases, 158,265 controls) (bottom).

| Locus | Candidate gene | CHR: POS | rsid | EA | OA | OR | 95 % Cl | p-value | Het PVal | Fin Enric. | Ref. |
|---|---|---|---|---|---|---|---|---|---|---|---|
| **Meta-analysis** | | | | | | | | | | | |
| **1p36.11** | *ARID1A* | 1:26695422 | rs114165349 | C | G | 1,18 | 1,12–1,24 | 2.85e-08 | 0.1742 | 1.58 | 9 |
| **1q21.1** | *PDZK1* | 1:145711421 | rs1967017 | G | A | 0,93 | 0,90–0,95 | 2.28e-10 | 0.01407 | 1.01 | 8,9 |
| **1q22** | *THBS3* | 1:155151361 | rs6676150 | C | G | 1,11 | 1,08–1,13 | 1.26e-20 | 0.2735 | 0.96 | 8,9 |
| **2p23.3** | *GCKR* | 2:27508073 | rs1260326 | C | T | 0,87 | 0,85–0,89 | 1.75e-34 | 0.0776 | 1.07 | 6–9 |
| **3p21.1** | *SFMBT1* | 3:53026537 | rs34353860 | AT | A | 1,09 | 1,07–1,11 | 7.72e-16 | 0.4252 | 0.97 | 7–9 |
| **4p16.1** | *SLC2A9* | 4:8818656 | rs149360043 | A | T | 0,76 | 0,67–0,85 | 3.04e-09 | 0.0339 | 1.89 | 6–9 |
| **4p16.1** | *SLC2A9* | 4:9925343 | rs13129697 | G | T | 0,69 | 0,66–0,72 | 7.94e-180 | 0.2116 | 0.96 | 6–9 |
| **4q21.3** | *MEPE* | 4:87047227 | rs200864767 | T | C | 1,67 | 1,49–1,85 | 4.53e-08 | 0.4176 | NA | 8 |
| **4q22.1** | *ABCG2* | 4:88131171 | rs2231142 | T | G | 1,77 | 1,73–1,81 | 3.95e-207 | 6.76e-08 | 0.66 | 7–9 |
| **4q23** | *ADH1B* | 4:99318162 | rs1229984 | C | T | 0,71 | 0,61–0,81 | 3.00e-11 | 0.0248 | 1.04 | 8 |
| **4q31.21** | *INPP4B* | 4:143262580 | rs10003529 | G | A | 0,94 | 0,92–0,96 | 4.94e-08 | 0.2487 | NA | 9 |
| **5q13.2** | *TMEM171* | 5:73135630 | rs549579 | T | C | 0,92 | 0,90–0,94 | 1.03e-12 | 0.5035 | 0.92 | 7–9 |
| **6p24.3** | *RREB1* | 6:7108316 | rs143321297 | TTCCTCCTCC | T | 0,92 | 0,90–0,94 | 2.98e-12 | 0.9950 | NA | 6–9 |
| **6p22.2** | *SLC17A1* | 6:25814852 | rs1165195 | G | T | 1,13 | 1,11–1,15 | 1.33e-27 | 0.2822 | 1.17 | 6–9 |
| **6p21.1** | *SLC22A7* | 6:43331320 | rs2279087 | G | T | 1,08 | 1,06–1,10 | 1.54e-10 | 0.3786 | 0.91 | 9 |
| **7q11.23** | *MLXIPL* | 7:73609551 | rs12531645 | A | G | 0,92 | 0,89–0,95 | 2.97e-10 | 0.0725 | 0.80 | 7–9,15 |
| **10q11.23** | *A1CF* | 10:50852341 | rs10994720 | A | G | 1,07 | 1,04–1,10 | 4.60e-08 | 0.07445 | 1.33 | 9,15 |
| **10q21.1** | *BICC1* | 10:58514676 | rs35464263 | C | CCT | 0,94 | 0,92–0,96 | 1.38e-08 | 0.6887 | 1.14 | 9 |
| **10q21.2** | *SLC16A9* | 10:59709780 | rs1171614 | C | T | 1,12 | 1,09–1,15 | 6.57e-15 | 0.3993 | 1.10 | 6–9 |
| **11p14.1** | *DCDC1* | 11:30730731 | rs683877 | T | C | 1,07 | 1,05–1,09 | 7.38e-09 | 0.4596 | 0.89 | 9 |
| **11q13.1** | *SLC22A11* | 11:64597394 | rs12363578 | T | C | 0,89 | 0,87–0,91 | 5.02e-21 | 0.0903 | 0.67 | 6–9 |
| **11q13.1** | *OVOL1* | 11:65789645 | rs5792371 | TG | T | 1,11 | 1,09–1,13 | 1.53e-21 | 0.2746 | 1.30 | 8 |
| **12q13.3** | *INHBC* | 12:57445390 | rs61352607 | T | G | 0,90 | 0,87–0,93 | 1.86e-15 | 0.2459 | 1.04 | 6–9 |
| **15q24.1** | *CYP1A2* | 15:74760586 | rs11857376 | G | A | 0,94 | 0,92–0,96 | 2.00e-08 | 0.3813 | 0.83 | 16 |
| **15q24.2** | *UBE2Q2* | 15:75843853 | rs2047866 | T | C | 1,07 | 1,05–1,09 | 9.43e-09 | 0.2757 | 0.85 | 8,9 |
| **15q26.3** | *IGF1R* | 15:98743751 | rs61168554 | G | A | 1,07 | 1,05–1,09 | 1.53e-08 | 0.1881 | 0.85 | 9,17 |
| **16q21** | ***CDH8*** | **16:63007457** | **rs186124051** | **T** | **C** | **0,76** | **0,66–0,86** | **9.76e-09** | **0.6460** | **1.27** | **Novel** |
| **16q22.1** | *CDH1* | 16:69541335 | rs62052820 | A | G | 1,09 | 1,06–1,12 | 3.94e-10 | 0.5215 | 0.93 | 9 |
| **17q22** | *HLF* | 17:55288785 | rs4299205 | G | T | 0,93 | 0,91–0,95 | 2.00e-09 | 0.6854 | NA | 17 |
| **19q13.33** | *ALDH16A1* | 19:48175672 | rs539226271 | G | T | 1,42 | 1,30–1,53 | 2.01e-09 | 0.5381 | 2.84 | 6,11 |
| **19q13.33** | *ALDH16A1* | 19:49467772 | rs752383928 | G | C | 3,38 | 3,28–3,48 | 2.53e-115 | 1.08e-08 | 20.09 | 6 |
| **22q13.31** | *PNPLA3* | 22:43928847 | rs738409 | G | C | 0,91 | 0,88–0,94 | 7.93e-12 | 0.9143 | 0.98 | 7–9 |
| **Subgroup analysis: Males** | | | | | | | | | | | |
| **17p13.2** | ***SLC25A11**** | **17:4914555** | **rs139336593** | **T** | **G** | **0,78** | **0,69–0,87** | **4.14e-08** | **NA** | **3.8** | **Novel** |

Candidate gene, a gene at a new locus the biological function of which is likely to explain the gout association; CHR: POS, chromosome and position (genome build hg38); rsid; SNP markers identification number; EA, effect allele; OA, other allele; OR, odds ratio; 95% Cl, odds ratio 95% confidence interval; p-value; HetPVal, p-value of heterogeneity; Fin Enric., enrichment in Finns (calculated FIN AF/NFEE AF in the Genome Aggregation Database [gnomAD], FIN AF is the allele frequency in Finns and NFEE AF is the allele frequency in Europeans (does not include Finns or Estonians)); REF, reference article in which a gout association was observed +/- 1 Mb in the vicinity of the lead variant.
* The locus near *SLC25A11* has been associated with gout in a GWAS restricted to a subgroup consisting of males.

**Table S5.** Effect size differences between sexes and age groups. In FinnGen, we selected the observed lead variants (Table S2.1), also novel lead variant observed in the male subgroup was added to the analysis. For theses variants, we calculated whether there were differences in the effects of the variants between the subgroups. For this, we used a two-tailed test, using subgroup effect estimates and their standard errors *((Effect_males-Effect_females)/ sqrt(standarderror_males$^2$+standarderror_females$^2$))*.

| Lead variant | Candidate gene | Beta females | Beta males | Se females | Se males | Sex P_diff | Beta o50 | Beta u50 | Se o50 | Se u50 | Age P_diff |
|---|---|---|---|---|---|---|---|---|---|---|---|
| 1:155151361:G:C | *THBS3* | 0.092 | 0.125 | 0.028 | 0.017 | 0.32 | 0.124 | 0.056 | 0.015 | 0.047 | 0.16 |
| 2:27508073:T:C | *GCKR* | -0.084 | -0.131 | 0.029 | 0.017 | 0.16 | -0.113 | -0.174 | 0.015 | 0.046 | 0.21 |
| 3:53026433:C:T | *SFMBT1* | 0.106 | 0.101 | 0.030 | 0.018 | 0.89 | 0.096 | 0.157 | 0.016 | 0.048 | 0.23 |
| 4:8827046:A:G | *SLC2A9* | 0.374 | 0.244 | 0.087 | 0.051 | 0.20 | 0.269 | 0.381 | 0.046 | 0.143 | 0.45 |
| 4:9925343:T:G | *SLC2A9* | -0.413 | -0.332 | 0.033 | 0.019 | **0.03** | -0.360 | -0.316 | 0.018 | 0.053 | 0.44 |
| 4:88124179:A:G | *ABCG2* | -0.366 | -0.544 | 0.047 | 0.027 | **0.001** | -0.488 | -0.596 | 0.025 | 0.072 | 0.15 |
| 6:7115300:T:A | *RREB1* | -0.043 | -0.114 | 0.030 | 0.018 | **0.04** | -0.099 | -0.094 | 0.016 | 0.050 | 0.92 |
| 6:25776721:G:A | *SLC17A1* | -0.049 | -0.132 | 0.029 | 0.017 | **0.01** | -0.110 | -0.113 | 0.016 | 0.048 | 0.94 |
| 6:43386693:A:G | *SLC22A7* | -0.081 | -0.087 | 0.029 | 0.017 | 0.88 | -0.094 | -0.013 | 0.016 | 0.049 | 0.11 |
| 11:64597394:C:T | *SLC22A11* | -0.085 | -0.101 | 0.031 | 0.018 | 0.65 | -0.095 | -0.108 | 0.017 | 0.050 | 0.80 |
| 11:65622419:G:C | *OVOL1* | 0.065 | 0.118 | 0.028 | 0.016 | 0.10 | 0.106 | 0.086 | 0.015 | 0.046 | 0.68 |
| 17:4914555:G:T | *SLC25A11* | 0.021 | -0.251 | 0.072 | 0.046 | **0.001** | -0.156 | -0.378 | 0.040 | 0.131 | 0.10 |
| 17:61172493:C:T | *BCAS3* | -0.047 | -0.119 | 0.035 | 0.021 | 0.08 | -0.098 | -0.114 | 0.019 | 0.058 | 0.79 |
| 19:48175672:T:G | *ALDH16A1* | 0.348 | 0.334 | 0.119 | 0.070 | 0.92 | 0.269 | 0.855 | 0.065 | 0.161 | **0.0008** |
| 19:49465749:C:G | *ALDH16A1* | 0.973 | 1.240 | 0.110 | 0.062 | **0.03** | 1.022 | 1.930 | 0.059 | 0.125 | **6.18e-11** |
| 22:43928847:C:G | *PNPLA3* | -0.087 | -0.097 | 0.033 | 0.020 | 0.80 | -0.087 | -0.164 | 0.018 | 0.055 | 0.18 |

**Table S6.** Colocalization results. All gout risk loci for which the association signal colocalized with the respective candidate gene expression in the tissues included in the study. Approximate Bayesian factor analyses were performed with the 'coloc.abf' function found in the 'coloc' R-library[4]. Candidate genes and genes closest to the association signal were selected for the analysis if the closest gene was a different gene than the candidate gene. For the analysis, we selected tissues that are relevant for gout, namely kidney cortex, liver, small intestine, and whole blood. Variant-gene expression associations (GTEx v8), for the analysis were downloaded (01/16/2023) from GTExportal (https://www.gtexportal.org/home/datasets). Colocalizations with a posterior probability ≥ 0,8 for the variant were considered significant[5]. PP4, posterior probability 4

| Tissue | Gene | PP4 |
|---|---|---|
| **Whole blood** | ARID1A | 0.913 |
| **Whole blood** | THBS3 | 0.993 |
| **Small intestine** | PDZK1 | 0.999 |
| **Small intestine** | THBS3 | 0.997 |
| **Small intestine** | SLC2A9 | 0.812 |
| **Kidney cortex** | HLF | 0.995 |
| **Liver** | THBS3 | 0.997 |
| **Liver** | SLC17A1 | 0.984 |
| **Liver** | SLC22A11 | 0.947 |

**Table S7.** Genetic correlations were calculated using LDSC-software[18], and were calculated between gout and 437 other phenotypes extracted from the GWAS database provided by the MRC Integrative Epidemiology Unit (IEU). Rg, genetic correlation coefficient value; se, standard error of genetic correlation, p, p-value; $p^{FDR}$, false discovery rate corrected p-value.

| Trait | rg | se | p | $p^{FDR}$ |
|---|---|---|---|---|
| **Concentration of small VLDL particles** | 0.3279 | 0.069 | 1.98e-06 | 7.66e-06 |
| **Knee pain for 3+ months** | 0.0855 | 0.0864 | 0.323 | 0.392 |
| **Time spent driving** | 0.081 | 0.0377 | 0.032 | 0.054 |
| **Smoking status: Current** | 0.0981 | 0.0371 | 0.008 | 0.016 |
| **Cholesterol in small VLDL** | 0.0925 | 0.0445 | 0.038 | 0.062 |
| **Vascular/heart problems diagnosed by doctor: Stroke** | 0.3618 | 0.1019 | 4,00e-04 | 0.001 |
| **Vitamin and mineral supplements: Multivitamins +/- minerals** | -0.0771 | 0.0481 | 0.108 | 0.159 |
| **Arm fat mass (right)** | 0.3302 | 0.0525 | 3.15e-10 | 6.00e-09 |
| **Forced expiratory volume in 1-second (FEV1)** | -0.1135 | 0.0243 | 2.99e-06 | 1.08e-05 |
| **Mouth/teeth dental problems: Toothache** | -0.02 | 0.0612 | 0.744 | 0.804 |
| **Ratio of docosahexaenoic acid to total fatty acids** | -0.1859 | 0.0377 | 8.30e-07 | 3.66e-06 |
| **Phospholipids to total lipids ratio in small HDL** | 0.0568 | 0.0434 | 0.191 | 0.255 |
| **Vitamin and mineral supplements: None of the above** | 0.0931 | 0.0485 | 0.055 | 0.088 |
| **HDL cholesterol** | -0.2672 | 0.0478 | 2.31e-08 | 1.43e-07 |
| **Potassium in urine** | 0.1039 | 0.0335 | 0.002 | 0.004 |
| **Total lipids in very large VLDL** | 0.25 | 0.0414 | 1.58e-09 | 1.62e-08 |
| **Pulse wave peak to peak time** | -0.2709 | 0.0592 | 4.71e-06 | 1.66e-05 |
| **Amyotrophic lateral sclerosis** | -0.0813 | 0.0712 | 0.254 | 0.324 |
| **LDL cholesterol** | -0.1377 | 0.0728 | 0.059 | 0.093 |
| **Types of transport used (excluding work): Cycle** | -0.1532 | 0.0501 | 0.002 | 0.005 |
| **Caudate volume** | 0.0092 | 0.0841 | 0.913 | 0.940 |
| **Type 2 diabetes** | 0.322 | 0.0641 | 5.01e-07 | 2.30e-06 |
| **Exposure to tobacco smoke at home** | 0.258 | 0.0542 | 1.96e-06 | 7.66e-06 |
| **Age at menopause (last menstrual period)** | -0.109 | 0.0331 | 0.001 | 0.002 |
| **Intelligence** | -0.1539 | 0.0259 | 2.92e-09 | 2.58-08 |
| **Alcohol intake versus 10 years previously** | 0.1871 | 0.0391 | 1.66e-06 | 6.65e-06 |
| **Triglycerides in HDL** | 0.2542 | 0.0476 | 9.49e-08 | 5.23e-07 |
| **Frequency of depressed mood in last 2 weeks** | 0.1525 | 0.0386 | 7.65e-05 | 0.0002 |
| **Total cholesterol** | 0.047 | 0.0456 | 0.303 | 0.374 |
| **Putamen volume** | 0.0103 | 0.0694 | 0.882 | 0.913 |
| **Phospholipids in small HDL** | 0.1308 | 0.0399 | 0.001 | 0.003 |
| **Ratio of polyunsaturated fatty acids to monounsaturated fatty acids** | -0.3463 | 0.0505 | 7.01e-12 | 4.71e-10 |
| **Mouth/teeth dental problems: Dentures** | 0.1178 | 0.0406 | 0.004 | 0.008 |
| **Glycoprotein acetyls** | 0.2534 | 0.0405 | 3.81e-10 | 6.00e-09 |
| **Neuroticism score** | 0.0787 | 0.0273 | 0.004 | 0.008 |
| **ICD10: H25 Senile cataract** | 0.1072 | 0.1028 | 0.297 | 0.369 |
| **Phospholipids in medium VLDL** | 0.4003 | 0.0809 | 7.53e-07 | 3.39e-06 |
| **Ever had stillbirth, spontaneous miscarriage or termination** | 0.1059 | 0.0542 | 0.051 | 0.081 |
| **Concentration of very large HDL particles** | -0.3316 | 0.0536 | 6.26e-10 | 8.62e-09 |
| **Illness, injury, bereavement, stress in last 2 years: Serious illness, injury or assault of a close relative** | 0.0562 | 0.0497 | 0.258 | 0.325 |
| **Medication for pain relief, constipation, heartburn: Paracetamol** | 0.1638 | 0.0376 | 1.35e-05 | 4.25e-05 |
| **Hearing difficulty/problems with background noise** | 0.05 | 0.0308 | 0.105 | 0.155 |
| **Mineral and other dietary supplements: Glucosamine** | -0.0785 | 0.0458 | 0.086 | 0.131 |
| **Triglycerides to total lipids ratio in very small VLDL** | 0.378 | 0.0655 | 7.88e-09 | 5.79e-08 |
| **Nervous feelings** | -0.0606 | 0.0287 | 0.035 | 0.058 |
| **Concentration of IDL particles** | 0.0768 | 0.0921 | 0.404 | 0.482 |
| **Ever unenthusiastic/disinterested for a whole week** | 0.105 | 0.0469 | 0.025 | 0.044 |
| **ICD10: M24 Other specific joint derangements** | 0.2737 | 0.1186 | 0.021 | 0.037 |
| **Bring up phlegm/sputum/mucus on most days** | 0.0831 | 0.0732 | 0.256 | 0.324 |
| **Fractured bone site(s): Ankle** | 0.1822 | 0.0994 | 0.067 | 0.104 |
| **Arm fat percentage (right)** | 0.3282 | 0.051 | 1.27e-10 | 3.30e-09 |
| **Alcohol intake frequency.** | 0.1388 | 0.0367 | 2,00e-04 | 0.0005 |
| **Femoral neck bone mineral density** | 0.0376 | 0.0469 | 0.423 | 0.501 |
| **Coronary artery disease** | 0.3311 | 0.0428 | 9.58e-15 | 2.11e-12 |
| **Uric acid** | 0.4487 | 0.0753 | 2.51e-09 | 2.31e-08 |
| **Total cholesterol in HDL** | -0.3526 | 0.0753 | 2.81e-06 | 1.03e-05 |
| **Tinnitus: No, never** | -0.066 | 0.0459 | 0.151 | 0.208 |
| **Triglycerides in very small VLDL** | 0.2961 | 0.0697 | 2.17e-05 | 6.59e-05 |
| **Total lipids in medium HDL** | -0.1007 | 0.0462 | 0.029 | 0.050 |
| **Serum total cholesterol** | -0.0342 | 0.1001 | 0.732 | 0.793 |
| **ICD10: N81 Female genital prolapse** | 0.118 | 0.0711 | 0.097 | 0.145 |
| **Exposure to tobacco smoke outside home** | 0.1965 | 0.0406 | 1.30e-06 | 5.31e-06 |
| **Qualifications: CSEs or equivalent** | 0.1908 | 0.0433 | 1.04e-05 | 3.46e-05 |
| **Concentration of very small VLDL particles** | 0.0355 | 0.0476 | 0.455 | 0.533 |

| Trait | rg | se | p | $p^{FDR}$ |
|---|---|---|---|---|
| **Age at menarche** | -0.1461 | 0.0339 | 1.61e-05 | 5.03e-05 |
| **Free cholesterol to total lipids ratio in medium LDL** | -0.3321 | 0.0508 | 6.17e-11 | 2.24e-09 |
| **Time spent watching television (TV)** | 0.1904 | 0.0273 | 2.93e-12 | 3.23e-10 |
| **Used an inhaler for chest within last hour** | 0.1486 | 0.1011 | 0.142 | 0.199 |
| **ICD10: Z80 Family history of malignant neoplasm** | -0.017 | 0.1622 | 0.917 | 0.941 |
| **Depressive symptoms** | 0.1368 | 0.0573 | 0.017 | 0.031 |
| **Current tobacco smoking** | 0.1171 | 0.0337 | 5,00e-04 | 0.001 |
| **Medication for cholesterol, blood pressure, diabetes, or take exogenous hormones: Insulin** | 0.3011 | 0.0995 | 0.003 | 0.006 |
| **Childhood intelligence** | -0.1474 | 0.0618 | 0.017 | 0.031 |
| **Phospholipids in very small VLDL** | 0.1024 | 0.0835 | 0.220 | 0.289 |
| **VLDL cholesterol** | 0.1057 | 0.0406 | 0.009 | 0.018 |
| **College completion** | -0.1933 | 0.0522 | 2,00e-04 | 0.0005 |
| **Phospholipids to total lipids ratio in large HDL** | 0.3656 | 0.0617 | 3.05e-09 | 2.63e-08 |
| **Concentration of chylomicrons and extremely large VLDL particles** | 0.2764 | 0.0464 | 2.59e-09 | 2.33e-08 |
| **Total lipids in small VLDL** | 0.1736 | 0.034 | 3.23e-07 | 1.55e-06 |
| **ICD10: N20 Calculus of kidney and ureter** | 0.1382 | 0.0699 | 0.048 | 0.078 |
| **Cerebral aneurysm** | 0.1474 | 0.0804 | 0.067 | 0.104 |
| **Lung cancer** | 0.0085 | 0.115 | 0.941 | 0.958 |
| **ICD10: D25 Leiomyoma of uterus** | 0.1087 | 0.0796 | 0.172 | 0.234 |
| **Type of tobacco previously smoked: Cigars or pipes** | -0.1157 | 0.1141 | 0.310 | 0.379 |
| **Number of children fathered** | 0.1363 | 0.0463 | 0.003 | 0.007 |
| **ICD10: I20 Angina pectoris** | 0.4324 | 0.0793 | 4.98e-08 | 2.82e-07 |
| **Average weekly intake of other alcoholic drinks** | 0.3176 | 0.3151 | 0.313 | 0.382 |
| **Major depressive disorder (ICD-10 coded)** | 0.2 | 0.0673 | 0.003 | 0.007 |
| **Total lipids in small HDL** | 0.1295 | 0.0395 | 0.001 | 0.002 |
| **ICD10: I10 Essential (primary) hypertension** | 0.4102 | 0.1537 | 0.008 | 0.015 |
| **Triglycerides to total lipids ratio in small VLDL** | 0.3386 | 0.0736 | 4.15e-06 | 1.48e-05 |
| **ICD10: M67 Other disorders of synovium and tendon** | 0.1717 | 0.1895 | 0.365 | 0.437 |
| **Neuroticism** | 0.06 | 0.0284 | 0.035 | 0.058 |
| **Concentration of small HDL particles** | 0.2699 | 0.1358 | 0.047 | 0.076 |
| **Total phospholipids in lipoprotein particles** | -0.0757 | 0.0507 | 0.135 | 0.190 |
| **Medication for pain relief, constipation, heartburn: Aspirin** | 0.3943 | 0.0695 | 1.41e-08 | 9.28e-08 |
| **Cholesterol in very small VLDL** | -0.1498 | 0.0597 | 0.012 | 0.022 |
| **Back pain for 3+ months** | 0.1528 | 0.0629 | 0.015 | 0.028 |
| **ICD10: R14 Flatulence and related conditions** | -0.1357 | 0.118 | 0.250 | 0.323 |
| **Mineral and other dietary supplements: Iron** | -0.1374 | 0.0807 | 0.089 | 0.134 |
| **Phospholipids to total lipids ratio in small LDL** | -0.2021 | 0.0448 | 6.47e-06 | 2.23e-05 |
| **Cholesteryl esters to total lipids ratio in chylomicrons and extremely large VLDL** | -0.1338 | 0.0649 | 0.039 | 0.064 |
| **Forced vital capacity (FVC), Best measure** | -0.124 | 0.0254 | 1.08e-06 | 4.59e-06 |
| **Cholesterol in large HDL** | -0.3126 | 0.0499 | 3.63e-10 | 6.00e-09 |
| **Reason for glasses/contact lenses: Other eye condition** | 0.111 | 0.1753 | 0.527 | 0.602 |
| **Fractured bone site(s): Arm** | 0.1421 | 0.1296 | 0.273 | 0.342 |
| **Impedance of arm (left)** | -0.1813 | 0.0384 | 2.28e-06 | 8.60e-06 |
| **Neo-conscientiousness** | 0.1336 | 0.1463 | 0.361 | 0.434 |
| **Triglycerides in medium HDL** | 0.2568 | 0.0486 | 1.30e-07 | 6.92e-07 |
| **Extreme waist-to-hip ratio** | 0.1512 | 0.0951 | 0.112 | 0.162 |
| **Remnant cholesterol (non-HDL, non-LDL -cholesterol)** | -0.0773 | 0.059 | 0.190 | 0.255 |
| **Duration of vigorous activity** | 0.0783 | 0.0379 | 0.039 | 0.063 |
| **Why reduced smoking: None of the above** | -0.0208 | 0.1472 | 0.887 | 0.916 |
| **Types of physical activity in last 4 weeks: None of the above** | 0.2933 | 0.0576 | 3.54e-07 | 1.68e-06 |
| **Free cholesterol** | -0.1026 | 0.1335 | 0.442 | 0.520 |
| **Qualifications: O levels/GCSEs or equivalent** | -0.2081 | 0.0334 | 4.51e-10 | 6.63e-09 |
| **Concentration of VLDL particles** | 0.1548 | 0.0385 | 5.76e-05 | 0.0002 |
| **Mineral and other dietary supplements: Selenium** | 0.0363 | 0.0693 | 0.601 | 0.672 |
| **Total lipids in IDL** | 0.0404 | 0.0917 | 0.659 | 0.736 |
| **ICD10: D12 Benign neoplasm of colon, rectum, anus and anal canal** | 0.0582 | 0.0511 | 0.255 | 0.324 |
| **Albumin** | -0.0018 | 0.0966 | 0.985 | 0.990 |
| **Degree of unsaturation** | -0.2752 | 0.0516 | 9.83e-08 | 5.35e-07 |
| **Total cholesterol in IDL** | -0.0319 | 0.0915 | 0.728 | 0.792 |
| **Eczema** | 0.1019 | 0.0872 | 0.243 | 0.315 |
| **HOMA-B** | 0.2616 | 0.0612 | 1.93e-05 | 5.90e-05 |
| **Free cholesterol in large LDL** | -0.2768 | 0.0786 | 4,00e-04 | 0.001 |
| **Triglycerides to total lipids ratio in IDL** | 0.4084 | 0.0677 | 1.63e-09 | 1.63e-08 |
| **Arm fat-free mass (left)** | 0.1962 | 0.0398 | 8.24e-07 | 3.66e-06 |
| **ICD10: I30 Acute pericarditis** | -0.27 | 0.1384 | 0.051 | 0.082 |
| **Cholesteryl esters in HDL** | -0.2717 | 0.0479 | 1.40e-08 | 9.28e-08 |
| **Cholesterol to total lipids ratio in medium HDL** | -0.3241 | 0.0559 | 6.86e-09 | 5.13e-08 |
| **Average diameter for LDL particles** | -0.3528 | 0.0597 | 3.44e-09 | 2.92e-08 |
| **Fractured bone site(s): Leg** | 0.0563 | 0.1391 | 0.686 | 0.760 |

| Trait | rg | se | p | $p^{FDR}$ |
|---|---|---|---|---|
| **Number of days/week of moderate physical activity 10+ minutes** | 0.0035 | 0.0358 | 0.922 | 0.943 |
| **Worry too long after embarrassment** | -0.0719 | 0.0271 | 0.008 | 0.016 |
| **ICD10: K21 Gastro-oesophageal reflux disease** | 0.2267 | 0.0797 | 0.004 | 0.009 |
| **Creatinine (enzymatic) in urine** | 0.1754 | 0.0311 | 1.66e-08 | 1.04e-07 |
| **Phospholipids in large HDL** | -0.2682 | 0.0456 | 4.14e-09 | 3.45e-08 |
| **Distance between home and job workplace** | -0.1287 | 0.0983 | 0.191 | 0.255 |
| **Total lipids in very small VLDL** | 0.0385 | 0.0448 | 0.390 | 0.466 |
| **Schizophrenia** | -0.0447 | 0.027 | 0.098 | 0.147 |
| **Vitamin and mineral supplements: Vitamin C** | -0.1017 | 0.0701 | 0.147 | 0.204 |
| **Cholesteryl esters to total lipids ratio in large HDL** | -0.3565 | 0.062 | 8.94e-09 | 6.36e-08 |
| **Frequency of stair climbing in last 4 weeks** | -0.0909 | 0.0492 | 0.065 | 0.102 |
| **Body fat percentage** | 0.3193 | 0.0484 | 4.18e-11 | 1.68e-09 |
| **Trunk fat-free mass** | 0.1343 | 0.0336 | 6.38e-05 | 0.0002 |
| **Pallidum volume** | 0.0152 | 0.0844 | 0.857 | 0.896 |
| **3-hydroxybutyrate** | 0.0052 | 0.111 | 0.962 | 0.976 |
| **Townsend deprivation index at recruitment** | 0.1375 | 0.0543 | 0.011 | 0.021 |
| **ICD10: H40 Glaucoma** | 0.0364 | 0.1007 | 0.718 | 0.785 |
| **Cholesteryl esters in large LDL** | -0.1558 | 0.0716 | 0.030 | 0.051 |
| **Birth weight** | -0.075 | 0.0291 | 0.010 | 0.019 |
| **Concentration of small LDL particles** | 0.0104 | 0.0537 | 0.846 | 0.888 |
| **Description of average fatty acid chain length, not actual carbon number** | -0.2609 | 0.0944 | 0.006 | 0.012 |
| **Cigarettes smoked per day** | 0.1679 | 0.0427 | 8.35e-05 | 0.0002 |
| **Total lipids in very small VLDL** | 0.1785 | 0.0789 | 0.024 | 0.041 |
| **Number of full brothers** | 0.1225 | 0.0652 | 0.060 | 0.095 |
| **Age started oral contraceptive pill** | -0.1902 | 0.0468 | 4.82e-05 | 0.0001 |
| **Infant head circumference** | -0.0857 | 0.0755 | 0.257 | 0.324 |
| **Trunk fat percentage** | 0.2948 | 0.0458 | 1.25e-10 | 3.30e-09 |
| **Cigarettes per Day** | 0.1679 | 0.0427 | 8.32e-05 | 0.0002 |
| **Total cholesterol in large LDL** | -0.006 | 0.0956 | 0.950 | 0.965 |
| **Vascular/heart problems diagnosed by doctor: Heart attack** | 0.3153 | 0.0578 | 4.79e-08 | 2.78e-07 |
| **Mean time to correctly identify matches** | -0.018 | 0.0276 | 0.515 | 0.591 |
| **Pain type(s) experienced in last month: Neck or shoulder pain** | 0.1621 | 0.0361 | 7.12e-06 | 2.43e-05 |
| **Qualifications: College or University degree** | -0.199 | 0.0266 | 7.95e-14 | 1.17e-11 |
| **Pain type(s) experienced in last month: Facial pain** | 6.52e-05 | 0.0866 | 0.999 | 0.999 |
| **Ischemic stroke** | 0.2376 | 0.0546 | 1.34e-05 | 4.25e-05 |
| **Fractured bone site(s): Spine** | 0.1197 | 0.2017 | 0.553 | 0.630 |
| **Phospholipids to total lipids ratio in chylomicrons and extremely large VLDL** | 0.3169 | 0.0666 | 1.98e-06 | 7.66e-06 |
| **Total lipids in large HDL** | -0.3585 | 0.071 | 4.44e-07 | 2.08e-06 |
| **Free cholesterol in HDL** | -0.2443 | 0.0473 | 2.44e-07 | 1.25e-06 |
| **Cholesteryl esters to total lipids ratio in medium HDL** | -0.3072 | 0.0533 | 8.36e-09 | 6.04e-08 |
| **Transport type for commuting to job workplace: Cycle** | -0.1032 | 0.0409 | 0.012 | 0.022 |
| **Other polyunsaturated fatty acids than 18:2** | 0.0618 | 0.1057 | 0.559 | 0.635 |
| **Medication for cholesterol, blood pressure, diabetes, or take exogenous hormones: None of the above** | -0.3647 | 0.0525 | 3.87e-12 | 3.41e-10 |
| **Hypermetropia** | 0.0758 | 0.0667 | 0.256 | 0.324 |
| **Triglycerides to total lipids ratio in small HDL** | 0.3094 | 0.0504 | 8.06e-10 | 9.87e-09 |
| **Lung adenocarcinoma** | 0.1062 | 0.1142 | 0.353 | 0.425 |
| **Total lipids in medium VLDL** | 0.1282 | 0.0383 | 8,00e-04 | 0.002 |
| **ICD10: I84 Haemorrhoids** | 0.093 | 0.0707 | 0.188 | 0.254 |
| **Triglycerides in large VLDL** | 0.4135 | 0.0803 | 2.59e-07 | 1.28e-06 |
| **Cholesterol in very large VLDL** | 0.2105 | 0.0368 | 1.03e-08 | 7.13e-08 |
| **ICD10: K62 Other diseases of anus and rectum** | 0.3255 | 0.1105 | 0.003 | 0.007 |
| **Total cholesterol in medium LDL** | 0.0292 | 0.1024 | 0.775 | 0.826 |
| **Cholesteryl esters to total lipids ratio in large LDL** | -0.0955 | 0.0534 | 0.074 | 0.113 |
| **Concentration of very large VLDL particles** | 0.2558 | 0.0414 | 6.71e-10 | 8.70e-09 |
| **Phospholipids in VLDL** | 0.1849 | 0.0362 | 3.21e-07 | 1.55e-06 |
| **Falls in the last year** | 0.16 | 0.0462 | 5,00e-04 | 0.001 |
| **Creatinine** | 0.1476 | 0.0918 | 0.108 | 0.159 |
| **ICD10: M17 Gonarthrosis [arthrosis of knee]** | 0.3077 | 0.0693 | 9.07e-06 | 3.05e-05 |
| **Transport type for commuting to job workplace: Car/motor vehicle** | 0.1562 | 0.0601 | 0.009 | 0.018 |
| **Phospholipids in large HDL** | -0.3495 | 0.0719 | 1.16e-06 | 4.86e-06 |
| **Leg fat mass (right)** | 0.3329 | 0.0514 | 9.74e-11 | 2.86e-09 |
| **Diastolic blood pressure** | 0.1791 | 0.0312 | 9.41e-09 | 6.59e-08 |
| **Cholesteryl esters to total lipids ratio in small VLDL** | -0.2423 | 0.0725 | 8,00e-04 | 0.002 |
| **Free cholesterol in medium LDL** | -0.1845 | 0.0794 | 0.020 | 0.036 |
| **Why reduced smoking: Illness or ill health** | 0.2369 | 0.1577 | 0.133 | 0.188 |
| **Headaches for 3+ months** | -0.014 | 0.0565 | 0.805 | 0.851 |
| **Alcohol drinker status: Never** | 0.0193 | 0.0516 | 0.708 | 0.777 |
| **Total fatty acids** | 0.1446 | 0.0356 | 4.76e-05 | 0.0001 |
| **Cholesteryl esters in large VLDL** | 0.1695 | 0.0362 | 2.78e-06 | 1.02e-05 |

| Trait | rg | se | p | $p^{FDR}$ |
|---|---|---|---|---|
| **Happiness** | 0.0669 | 0.0436 | 0.125 | 0.178 |
| **Total triglycerides** | 0.2532 | 0.041 | 6.69e-10 | 8.70e-09 |
| **Number of treatments/medications taken** | 0.3916 | 0.0463 | 2.65e-17 | 1.17e-14 |
| **Total lipids in small VLDL** | 0.3087 | 0.0704 | 1.17e-05 | 3.73e-05 |
| **ICD10: R35 Polyuria** | -0.0267 | 0.131 | 0.839 | 0.883 |
| **Miserableness** | 0.0976 | 0.0367 | 0.008 | 0.015 |
| **Average diameter for HDL particles** | -0.2875 | 0.0473 | 1.24e-09 | 1.40e-08 |
| **Length of working week for main job** | 0.0739 | 0.0563 | 0.190 | 0.255 |
| **Target heart rate achieved** | -0.0291 | 0.0743 | 0.695 | 0.766 |
| **Concentration of medium LDL particles** | 0.0809 | 0.0988 | 0.413 | 0.491 |
| **Anorexia Nervosa** | -0.0179 | 0.0599 | 0.766 | 0.820 |
| **ICD10: K35 Acute appendicitis** | 0.1583 | 0.1379 | 0.251 | 0.324 |
| **Job involves mainly walking or standing** | 0.1474 | 0.0343 | 1.75e-05 | 5.40e-05 |
| **Vitamin and mineral supplements: Vitamin A** | -0.0501 | 0.0883 | 0.571 | 0.643 |
| **Sleeplessness / insomnia** | 0.1482 | 0.0279 | 1.12e-07 | 6.01e-07 |
| **ICD10: K44 Diaphragmatic hernia** | 0.2796 | 0.0932 | 0.003 | 0.006 |
| **Citrate** | -0.0967 | 0.0848 | 0.254 | 0.324 |
| **Glutamine** | -0.2141 | 0.0452 | 2.16e-06 | 8.23e-06 |
| **Tense / highly strung** | 0.0912 | 0.0315 | 0.004 | 0.008 |
| **ICD10: N40 Hyperplasia of prostate** | -0.0027 | 0.0734 | 0.971 | 0.980 |
| **Cholesteryl esters in very large HDL** | -0.3449 | 0.0549 | 3.36e-10 | 6.00e-09 |
| **Wheeze or whistling in the chest in last year** | 0.2694 | 0.0435 | 6.16e-10 | 8.62e-09 |
| **Free cholesterol in very small VLDL** | -0.0094 | 0.0495 | 0.849 | 0.890 |
| **Alcohol usually taken with meals** | -0.1904 | 0.0405 | 2.65e-06 | 9.82e-06 |
| **Triglycerides in small VLDL** | 0.2486 | 0.0397 | 3.70e-10 | 6.00e-09 |
| **ICD10: S52 Fracture of forearm** | 0.047 | 0.0725 | 0.517 | 0.592 |
| **Ever stopped smoking for 6+ months** | 0.1509 | 0.1231 | 0.220 | 0.289 |
| **Handedness (chirality/laterality): Use both right and left hands equally** | 0.1257 | 0.0873 | 0.150 | 0.2085 |
| **Cataract** | 0.0589 | 0.0867 | 0.497 | 0.574 |
| **ICD10: J33 Nasal polyp** | 0.0461 | 0.1214 | 0.704 | 0.774 |
| **Daytime dozing / sleeping (narcolepsy)** | 0.1013 | 0.0385 | 0.009 | 0.017 |
| **Adopted as a child** | 0.0983 | 0.0611 | 0.108 | 0.159 |
| **Medication for pain relief, constipation, heartburn: Ibuprofen (e.g. Nurofen)** | 0.1082 | 0.0379 | 0.004 | 0.009 |
| **ICD10: K40 Inguinal hernia** | 0.0194 | 0.055 | 0.724 | 0.790 |
| **Doctor diagnosed hayfever or allergic rhinitis** | -5.44e-05 | 0.0393 | 0.999 | 0.999 |
| **Omega-6 fatty acids** | 0.1195 | 0.1279 | 0.350 | 0.423 |
| **Number of depression episodes** | -0.0145 | 0.0877 | 0.869 | 0.903 |
| **Waist-to-hip ratio** | 0.3239 | 0.0587 | 3.43e-08 | 2.042e-07 |
| **Leg fat-free mass (right)** | 0.1867 | 0.0408 | 4.76e-06 | 1.67e-05 |
| **Omega-3 fatty acids** | 0.0875 | 0.0415 | 0.035 | 0.058 |
| **Illness, injury, bereavement, stress in last 2 years: Death of a close relative** | 0.0809 | 0.0604 | 0.181 | 0.245 |
| **Illness, injury, bereavement, stress in last 2 years: Marital separation/divorce** | 0.0664 | 0.0872 | 0.446 | 0.524 |
| **HbA1c** | 0.0789 | 0.0511 | 0.122 | 0.175 |
| **Chronic kidney disease** | 0.3319 | 0.1114 | 0.003 | 0.006 |
| **Hand grip strength (right)** | -0.0718 | 0.0285 | 0.012 | 0.022 |
| **Vitamin and mineral supplements: Folic acid or Folate (Vit B9)** | 0.0353 | 0.109 | 0.746 | 0.804 |
| **Cholesterol lowering medication** | 0.435 | 0.065 | 2.22e-11 | 1.22e-09 |
| **Free cholesterol to total lipids ratio in small LDL** | -0.3203 | 0.0564 | 1.36e-08 | 9.23e-08 |
| **Mean diameter for VLDL particles** | 0.3898 | 0.0725 | 7.76e-08 | 4.33e-07 |
| **Handedness (chirality/laterality): Left-handed** | 0.0018 | 0.0628 | 0.977 | 0.9845 |
| **Free cholesterol to total lipids ratio in medium HDL** | -0.3028 | 0.0544 | 2.65e-08 | 1.62e-07 |
| **Reason for reducing amount of alcohol drunk: Illness or ill health** | 0.3665 | 0.1123 | 0.001 | 0.003 |
| **Underlying (primary) cause of death: ICD10: J84.1 Other interstitial pulmonary diseases with fibrosis** | -0.0189 | 0.1812 | 0.917 | 0.941 |
| **Total cholesterol in very large HDL** | -0.4956 | 0.211 | 0.019 | 0.034 |
| **Primary biliary cirrhosis** | 0.0864 | 0.0847 | 0.308 | 0.378 |
| **Smoking behaviors : Smoking cessation** | -0.107 | 0.0665 | 0.108 | 0.159 |
| **Phospholipids to total lipids ratio in very large HDL** | -0.2748 | 0.0468 | 4.30e-09 | 3.52e-08 |
| **Number of older siblings** | 0.2011 | 0.096 | 0.036 | 0.060 |
| **ICD10: I48 Atrial fibrillation and flutter** | 0.1438 | 0.0518 | 0.006 | 0.011 |
| **Pain type(s) experienced in last month: Knee pain** | 0.2067 | 0.0431 | 1.60e-06 | 6.47e-06 |
| **Oral contraceptive pill or minipill** | -0.1977 | 0.1094 | 0.071 | 0.109 |
| **Apolipoprotein A-I** | -0.1976 | 0.0911 | 0.030 | 0.051 |
| **Total cholesterol in medium HDL** | -0.151 | 0.0969 | 0.119 | 0.172 |
| **Total lipids in medium LDL** | 0.0176 | 0.0566 | 0.756 | 0.813 |
| **Mean platelet volume** | 0.0389 | 0.0278 | 0.163 | 0.223 |
| **Concentration of large HDL particles** | -0.3528 | 0.0701 | 4.89e-07 | 2.27e-06 |
| **ICD10: R10 Abdominal and pelvic pain** | 0.3476 | 0.0998 | 5,00e-04 | 0.001 |
| **Emphysema/chronic bronchitis** | 0.2063 | 0.052 | 7.31e-05 | 0.0002 |
| **Impedance of whole body** | -0.1822 | 0.0387 | 2.49e-06 | 9.29e-06 |

| Trait | rg | se | p | $p^{FDR}$ |
|---|---|---|---|---|
| **Blood pressure medication** | 0.3486 | 0.0509 | 7.48e-12 | 4.71e-10 |
| **Sodium in urine** | 0.1921 | 0.0503 | 1,00e-04 | 0.0003 |
| **Free cholesterol to total lipids ratio in medium VLDL** | -0.325 | 0.0662 | 9.31e-07 | 4.06e-06 |
| **Phosphatidylcholines** | -0.073 | 0.0481 | 0.129 | 0.182 |
| **Alanine** | 0.1274 | 0.0969 | 0.189 | 0.254 |
| **Acetoacetate** | 0.0227 | 0.0949 | 0.811 | 0.856 |
| **Ever had prostate specific antigen (PSA) test** | -0.0566 | 0.0442 | 0.200 | 0.266 |
| **Time from waking to first cigarette** | -0.2566 | 0.0653 | 8.41e-05 | 0.0002 |
| **Cholesteryl esters in small HDL** | 0.0458 | 0.044 | 0.300 | 0.369 |
| **Concentration of very large HDL particles** | -0.4148 | 0.1225 | 7,00e-04 | 0.002 |
| **Ratio of triglycerides to phosphoglycerides** | 0.2874 | 0.0458 | 3.59e-10 | 6.00e-09 |
| **Concentration of very large VLDL particles** | 0.3479 | 0.0785 | 9.45e-06 | 3.16e-05 |
| **Total lipids in very large HDL** | -0.3269 | 0.0524 | 4.51e-10 | 6.63e-09 |
| **Forced expiratory volume in 1-second (FEV1), predicted** | -0.026 | 0.0328 | 0.427 | 0.504 |
| **Circulating leptin levels** | 0.3956 | 0.0829 | 1.84e-06 | 7.317e-06 |
| **Mean diameter for HDL particles** | -0.3718 | 0.0761 | 1.03e-06 | 4.45e-06 |
| **Concentration of very small VLDL particles** | 0.1998 | 0.0761 | 0.009 | 0.017 |
| **ICD10: F43 Reaction to severe stress and adjustment disorders** | 0.4601 | 0.4143 | 0.267 | 0.335 |
| **Total fatty acids** | 0.339 | 0.1294 | 0.009 | 0.017 |
| **Morning/evening person (chronotype)** | -0.0587 | 0.0232 | 0.011 | 0.021 |
| **Triglycerides in very large HDL** | -0.0547 | 0.0801 | 0.495 | 0.572 |
| **Apolipoprotein B** | 0.1714 | 0.0966 | 0.076 | 0.116 |
| **Number of operations, self-reported** | 0.2271 | 0.0518 | 1.14e-05 | 3.70e-05 |
| **Total lipids in very large VLDL** | 0.3308 | 0.0739 | 7.53e-06 | 2.55e-05 |
| **Frequency of unenthusiasm / disinterest in last 2 weeks** | 0.1727 | 0.0394 | 1.15e-05 | 3.71e-05 |
| **Fracture resulting from simple fall** | 0.0812 | 0.0771 | 0.293 | 0.366 |
| **Operation code: bilateral oophorectomy** | 0.251 | 0.0517 | 1.18e-06 | 4.93e-06 |
| **Total lipids in VLDL** | 0.2013 | 0.0364 | 3.29e-08 | 1.99e-07 |
| **Apolipoprotein B** | 0.0397 | 0.0349 | 0.255 | 0.324 |
| **ICD10: N32 Other disorders of bladder** | 0.0808 | 0.0848 | 0.341 | 0.413 |
| **Acetoacetate** | 0.205 | 0.0624 | 0.001 | 0.002 |
| **Average weekly champagne plus white wine intake** | -0.1322 | 0.0469 | 0.005 | 0.010 |
| **Number of cigarettes currently smoked daily (current cigarette smokers)** | 0.165 | 0.08 | 0.039 | 0.064 |
| **Mouth/teeth dental problems: Mouth ulcers** | 0.0304 | 0.0426 | 0.475 | 0.553 |
| **Weight change compared with 1 year ago** | 0.0754 | 0.0489 | 0.123 | 0.175 |
| **Total lipids in large VLDL** | 0.2247 | 0.0387 | 6.24e-09 | 4.82e-08 |
| **ICD10: O75 Other complications of labour and delivery not elsewhere classified** | 0.0091 | 0.1113 | 0.935 | 0.954 |
| **ICD10: R11 Nausea and vomiting** | 0.2588 | 0.1623 | 0.111 | 0.162 |
| **ICD10: M54 Dorsalgia** | 0.3007 | 0.0661 | 5.40e-06 | 1.88e-05 |
| **Total lipids in large VLDL** | 0.3719 | 0.0723 | 2.66e-07 | 1.31e-06 |
| **Blood clot in the leg (DVT)** | 0.2643 | 0.0601 | 1.11e-05 | 3.65e-05 |
| **Triglycerides in VLDL** | 0.2466 | 0.0411 | 2.02e-09 | 1.94e-08 |
| **Cholesteryl esters in small LDL** | 0.0149 | 0.0528 | 0.778 | 0.827 |
| **Cholesterol to total lipids ratio in very large HDL** | 0.066 | 0.0434 | 0.128 | 0.182 |
| **Maximum workload during fitness test** | -0.3084 | 0.0754 | 4.31e-05 | 0.0001 |
| **Phospholipids to total lipids ratio in large LDL** | -0.0616 | 0.0506 | 0.224 | 0.293 |
| **Total lipids in lipoprotein particles** | -0.0083 | 0.0499 | 0.868 | 0.903 |
| **Guilty feelings** | 0.0787 | 0.0345 | 0.023 | 0.040 |
| **Free cholesterol to total lipids ratio in chylomicrons and extremely large VLDL** | -0.188 | 0.0635 | 0.003 | 0.007 |
| **Mineral and other dietary supplements: Zinc** | -0.1116 | 0.0529 | 0.035 | 0.058 |
| **Average number of methylene groups in a fatty acid chain** | 0.3726 | 0.2656 | 0.161 | 0.221 |
| **Free cholesterol in chylomicrons and extremely large VLDL** | 0.2744 | 0.0454 | 1.46e-09 | 1.57e-08 |
| **ICD10: H26.9 Cataract, unspecified** | 0.0989 | 0.068 | 0.146 | 0.203 |
| **Had major operations** | 0.3032 | 0.0966 | 0.002 | 0.004 |
| **Cholesterol esters in large VLDL** | 0.0149 | 0.0933 | 0.873 | 0.906 |
| **Cholesterol esters in medium HDL** | -0.165 | 0.1065 | 0.121 | 0.174 |
| **Smoking status: Never** | -0.1347 | 0.0343 | 8.48e-05 | 0.0002 |
| **Phospholipids in medium HDL** | -0.0525 | 0.0936 | 0.575 | 0.647 |
| **Work/job satisfaction** | 0.034 | 0.0643 | 0.597 | 0.670 |
| **Former alcohol drinker** | 0.0784 | 0.0614 | 0.201 | 0.267 |
| **Triglycerides in medium VLDL** | 0.2086 | 0.0369 | 1.63e-08 | 1.041e-07 |
| **Fractured/broken bones in last 5 years** | 0.0607 | 0.0381 | 0.111 | 0.162 |
| **ICD10: Z09 Follow-up examination after treatment for conditions other than malignant neoplasms** | 0.2634 | 0.1108 | 0.017 | 0.031 |
| **Cholesteryl esters in medium VLDL** | -0.2479 | 0.0792 | 0.002 | 0.004 |
| **Pain type(s) experienced in last month: Headache** | 0.0508 | 0.0276 | 0.066 | 0.104 |
| **Concentration of HDL particles** | -0.1274 | 0.0476 | 0.008 | 0.015 |
| **Mineral and other dietary supplements: Calcium** | -0.2251 | 0.0575 | 9.14e-05 | 0.0002 |
| **Cholesterol esters in large HDL** | -0.3819 | 0.0741 | 2.59e-07 | 1.28e-06 |
| **Whole body fat mass** | 0.3181 | 0.0501 | 2.16e-10 | 5.02e-09 |

| Trait | rg | se | p | $p^{FDR}$ |
|---|---|---|---|---|
| **Noisy workplace** | 0.1859 | 0.0583 | 0.0014 | 0.003 |
| **Forced vital capacity (FVC)** | -0.1311 | 0.0254 | 2.54e-07 | 1.28e-06 |
| **Wears glasses or contact lenses** | -0.1057 | 0.0451 | 0.019 | 0.034 |
| **Apolipoprotein A1** | -0.1567 | 0.0461 | 7,00e-04 | 0.002 |
| **Concentration of large HDL particles** | -0.302 | 0.0493 | 8.76e-10 | 1.04e-08 |
| **ICD10: J34 Other disorders of nose and nasal sinuses** | 0.1698 | 0.1671 | 0.310 | 0.379 |
| **ICD10: M25 Other joint disorders, not elsewhere classified** | 0.4601 | 0.0968 | 2.00e-06 | 7.66e-06 |
| **Cholesterol esters in medium VLDL** | 0.276 | 0.0693 | 6.75e-05 | 0.0002 |
| **Leucine** | 0.3169 | 0.1324 | 0.017 | 0.030 |
| **Ever taken oral contraceptive pill** | -0.112 | 0.0534 | 0.036 | 0.060 |
| **Ever used hormone-replacement therapy (HRT)** | 0.0993 | 0.0385 | 0.010 | 0.019 |
| **Eye problems/disorders: Injury or trauma resulting in loss of vision** | 0.1551 | 0.1134 | 0.171 | 0.234 |
| **Ever depressed for a whole week** | 0.0225 | 0.0395 | 0.570 | 0.643 |
| **Offspring birth weight** | -0.0621 | 0.0528 | 0.239 | 0.311 |
| **Sensitivity / hurt feelings** | 0.0888 | 0.0272 | 0.001 | 0.003 |
| **ICD10: S76 Injury of muscle and tendon at hip and thigh level** | -0.0426 | 0.1384 | 0.758 | 0.813 |
| **Pulse rate, automated reading** | 0.0794 | 0.025 | 0.002 | 0.003 |
| **Cholesterol to total lipids ratio in IDL** | -0.3897 | 0.0668 | 5.31e-09 | 4.18e-08 |
| **Ratio of linoleic acid to total fatty acids** | -0.4385 | 0.0719 | 1.05e-09 | 1.22e-08 |
| **Phospholipids in medium LDL** | 0.0686 | 0.1015 | 0.499 | 0.575 |
| **Phospholipids in IDL** | 0.0762 | 0.0975 | 0.435 | 0.513 |
| **Myopia** | -0.09 | 0.0335 | 0.007 | 0.014 |
| **Squamous cell lung cancer** | -0.0041 | 0.0937 | 0.965 | 0.976 |
| **Free cholesterol in large VLDL** | 0.2296 | 0.0383 | 2.03e-09 | 1.94e-08 |
| **Lactate** | 0.1923 | 0.0579 | 9,00e-04 | 0.002 |
| **Phospholipids in medium VLDL** | 0.071 | 0.0431 | 0.100 | 0.149 |
| **Ever had hysterectomy (womb removed)** | 0.2982 | 0.0616 | 1.28e-06 | 5.26e-06 |
| **Cholesterol to total lipids ratio in very large VLDL** | -0.3193 | 0.0608 | 1.49e-07 | 7.84e-07 |
| **Alcohol drinker status: Current** | -0.1055 | 0.0479 | 0.028 | 0.048 |
| **Ferritin** | 0.037 | 0.091 | 0.684 | 0.760 |
| **Glucose** | 0.0376 | 0.0646 | 0.560 | 0.635 |
| **Blood clot in the lung** | 0.2375 | 0.0718 | 9,00e-04 | 0.002 |
| **Childhood asthma (age<16)** | 0.4323 | 0.1288 | 8,00e-04 | 0.002 |
| **Triglycerides in large HDL** | 0.0835 | 0.0424 | 0.049 | 0.079 |
| **Diagnoses - main ICD10: E04 Other non-toxic goitre** | 0.0409 | 0.0969 | 0.673 | 0.750 |
| **Frequency of tiredness / lethargy in last 2 weeks** | 0.1733 | 0.0411 | 2.44e-05 | 7.30e-05 |
| **Phospholipids in medium HDL** | -0.0555 | 0.0464 | 0.231 | 0.301 |
| **Trunk fat mass** | 0.3008 | 0.0476 | 2.62e-10 | 5.77e-09 |
| **Types of physical activity in last 4 weeks: Other exercises** | -0.1521 | 0.0388 | 9.03e-05 | 0.0002 |
| **Total lipids in HDL** | -0.1988 | 0.0453 | 1.13e-05 | 3.69e-05 |
| **Cholesteryl esters to total lipids ratio in IDL** | -0.3985 | 0.0686 | 6.34e-09 | 4.82e-08 |
| **ICD10: I25 Chronic ischaemic heart disease** | 0.3664 | 0.0596 | 7.79e-10 | 9.82e-09 |
| **Ratio of bisallylic groups to total fatty acids** | -0.3065 | 0.0852 | 3,00e-04 | 0.0008 |
| **Impedance of arm (right)** | -0.1763 | 0.0378 | 3.03e-06 | 1.08e-05 |
| **Concentration of large VLDL particles** | 0.3683 | 0.0735 | 5.37e-07 | 2.44e-06 |
| **Phospholipids to total lipids ratio in very large VLDL** | 0.0146 | 0.057 | 0.798 | 0.846 |
| **Ever smoked** | 0.0838 | 0.0326 | 0.010 | 0.019 |
| **ICD10: K20 Oesophagitis** | 0.2307 | 0.0858 | 0.007 | 0.015 |
| **Length of menstrual cycle** | 0.1692 | 0.0526 | 0.001 | 0.003 |
| **Triglycerides in small HDL** | 0.2986 | 0.0475 | 3.34e-10 | 6.00e-09 |
| **Polyunsaturated fatty acids** | -0.0175 | 0.0438 | 0.689 | 0.762 |
| **Snoring** | -0.1401 | 0.0371 | 2,00e-04 | 0.0005 |
| **Loud music exposure frequency** | 0.0316 | 0.046 | 0.493 | 0.572 |
| **Duration of moderate activity** | 0.0553 | 0.0385 | 0.151 | 0.208 |
| **Weight** | 0.2803 | 0.0478 | 4.60e-09 | 3.69e-08 |
| **Phospholipids in very large VLDL** | 0.2443 | 0.0408 | 2.21e-09 | 2.08e-08 |
| **Number of days/week of vigorous physical activity 10+ minutes** | 0.0136 | 0.0394 | 0.760 | 0.792 |
| **Pain type(s) experienced in last month: Stomach or abdominal pain** | 0.139 | 0.0425 | 0.001 | 0.003 |
| **Hippocampus volume** | -0.1442 | 0.0799 | 0.071 | 0.109 |
| **multiple sclerosis** | 0.0102 | 0.0355 | 0.774 | 0.826 |
| **Total cholesterol minus HDL-C** | -0.1102 | 0.0662 | 0.096 | 0.144 |
| **Total lipids in small HDL** | 0.2188 | 0.1197 | 0.068 | 0.105 |
| **Non-accidental death in close genetic family** | 0.2319 | 0.0833 | 0.005 | 0.011 |
| **Heel bone mineral density (BMD) T-score, automated** | 0.0646 | 0.0247 | 0.009 | 0.017 |
| **Diastolic blood pressure, automated reading** | 0.2777 | 0.0418 | 2.95e-11 | 1.30-09 |
| **Triglycerides in small LDL** | 0.2524 | 0.04 | 2.80e-10 | 5.89e-09 |
| **ICD10: M06.99 Rheumatoid arthritis, unspecified** | 0.1121 | 0.1097 | 0.307 | 0.378 |
| **Number of pregnancy terminations** | -0.0525 | 0.0503 | 0.297 | 0.369 |
| **Phospholipids to total lipids ratio in small VLDL** | -0.3674 | 0.0703 | 1.75e-07 | 9.07e-07 |
| **Basal metabolic rate** | 0.1998 | 0.041 | 1.08e-06 | 4.59e-06 |

| Trait | rg | se | p | $p^{FDR}$ |
|---|---|---|---|---|
| **Total lipids in large HDL** | -0.2884 | 0.0477 | 1.52e-09 | 1.60e-08 |
| **3-Hydroxybutyrate** | 0.1134 | 0.0507 | 0.025 | 0.044 |
| **Triglycerides in chylomicrons and extremely large VLDL** | 0.2651 | 0.0483 | 3.91e-08 | 2.30e-07 |
| **Mono-unsaturated fatty acids** | 0.4286 | 0.1068 | 5.95e-05 | 0.0002 |
| **Time spent using computer** | -0.0598 | 0.0248 | 0.016 | 0.029 |
| **Free cholesterol to total lipids ratio in large LDL** | -0.3568 | 0.0557 | 1.55e-10 | 3.79e-09 |
| **Free cholesterol in medium HDL** | -0.1699 | 0.0451 | 2,00e-04 | 0.0005 |
| **Ratio of saturated fatty acids to total fatty acids** | 0.2434 | 0.0668 | 3,00e-04 | 0.0008 |
| **Subjective well being** | -0.0848 | 0.0538 | 0.115 | 0.166 |
| **Mouth/teeth dental problems: None of the above** | -0.1114 | 0.0459 | 0.015 | 0.028 |
| **Average weekly fortified wine intake** | -0.1687 | 0.0582 | 0.004 | 0.008 |
| **Free cholesterol in very large HDL** | -0.3246 | 0.0535 | 1.30e-09 | 1.43e-08 |
| **Qualifications: NVQ or HND or HNC or equivalent** | 0.1233 | 0.0576 | 0.033 | 0.055 |
| **Forced expiratory volume in 1-second (FEV1), Best measure** | -0.0993 | 0.0235 | 2.45e-05 | 7.30e-05 |
| **Triglycerides in large LDL** | 0.2692 | 0.0414 | 7.55e-11 | 2.38e-09 |
| **Cholesteryl esters to total lipids ratio in medium VLDL** | -0.3494 | 0.0618 | 1.58e-08 | 1.02e-07 |
| **Difference in height between adolescence and adulthood** | -0.3076 | 0.0852 | 3,00e-04 | 0.0008 |
| **Leucine** | 0.2846 | 0.0672 | 2.26e-05 | 6.81e-05 |
| **Valine** | 0.2484 | 0.0614 | 5.27e-05 | 0.0002 |
| **Phospholipids in very large VLDL** | 0.3352 | 0.078 | 1.72e-05 | 5.34e-05 |
| **Concentration of medium HDL particles** | -0.1403 | 0.0458 | 0.002 | 0.005 |
| **Tinnitus: Yes, but not now, but have in the past** | 0.1518 | 0.0765 | 0.047 | 0.077 |
| **Chest pain or discomfort walking normally** | 0.3452 | 0.0517 | 2.55e-11 | 1.25e-09 |
| **Job involves heavy manual or physical work** | 0.1636 | 0.03 | 4.95e-08 | 2.82-07 |
| **Phosphoglycerides** | -0.0344 | 0.0472 | 0.465 | 0.543 |
| **ICD10: K30 Dyspepsia** | 0.1142 | 0.0912 | 0.211 | 0.278 |
| **Cholesterol in large LDL** | -0.1912 | 0.0713 | 0.007 | 0.015 |

**Table S8.** List of LD-based proxies used in metabolic association profiles of gout risk-increasing alleles. We estimated proxies using the LDproxy tool available in LDlink applications[19] for the gout-associated variants that were not found in the metabolomics data. CHR:POS (hg19), chromosome and (genome build hg19), Proxy (hg19), chromosome and position of the used LD-based proxy (genome build hg19), Proxy rsid, SNP markers identification marker, $R^2$ squared correlation that has been used to measure LD between the original lead variant and proxy in Europeans.

| Identifier | CHR:POS **(hg19)** | Proxy **(hg19)** | **Proxy rsid** | Correlative alleles | $R^2$ **(EUR)** |
|---|---|---|---|---|---|
| ***PDZK1*** rs1967017-A | 1:145723643 | 1:145723120 | rs10910845 | A=C, G=T | 1 |
| ***SFMBT1*** rs34353860-AT | 3:53060553 | 3:53062661 | rs9847710 | AT=T, A=C | 0.90 |
| ***MLXIPL*** rs12531645-G | 7:73023881 | 7:73035857 | rs7800944 | G=T, A=C | 0.97 |
| ***BICC1*** rs35464263-CCT | 10:60274436 | 10:60291548 | rs1593678 | CCT=G, C=A | 1 |
| ***OVOL1*** rs5792371-TG | 11:65557116 | 11:65555458 | rs4930319 | TG=C, T=G | 0.99 |
| ***CYP1A2*** rs11857376-A | 15:75052927 | 15:75052495 | rs12903896 | A=C, G=T | 0.56 |

**Table S9.** A list of FinnGen authors and their affiliations

| Full Name | Affiliation | Role 1 | Role 2 |
|---|---|---|---|
| **Aarno Palotie** | Institute for Molecular Medicine Finland (FIMM), HiLIFE, University of Helsinki, Helsinki, Finland; Broad Institute of MIT and Harvard; Massachusetts General Hospital | **Steering Committee** | **Steering Committee** |
| **Mark Daly** | Institute for Molecular Medicine Finland (FIMM), HiLIFE, University of Helsinki, Helsinki, Finland; Broad Institute of MIT and Harvard; Massachusetts General Hospital | **Steering Committee** | **Steering Committee** |
| **Bridget Riley-Gills** | Abbvie, Chicago, IL, United States | **Steering Committee** | **Pharmaceutical companies** |
| **Howard Jacob** | Abbvie, Chicago, IL, United States | **Steering Committee** | **Pharmaceutical companies** |
| **Dirk Paul** | Astra Zeneca, Cambridge, United Kingdom | **Steering Committee** | **Pharmaceutical companies** |
| **Slavé Petrovski** | Astra Zeneca, Cambridge, United Kingdom | **Steering Committee** | **Pharmaceutical companies** |
| **Heiko Runz** | Biogen, Cambridge, MA, United States | **Steering Committee** | **Pharmaceutical companies** |
| **Sally John** | Biogen, Cambridge, MA, United States | **Steering Committee** | **Pharmaceutical companies** |
| **George Okafo** | Boehringer Ingelheim, Ingelheim am Rhein, Germany | **Steering Committee** | **Pharmaceutical companies** |
| **Nathan Lawless** | Boehringer Ingelheim, Ingelheim am Rhein, Germany | **Steering Committee** | **Pharmaceutical companies** |
| **Heli Salminen-Mankonen** | Boehringer Ingelheim, Ingelheim am Rhein, Germany | **Steering Committee** | **Pharmaceutical companies** |
| **Robert Plenge** | Bristol Myers Squibb, New York, NY, United States | **Steering Committee** | **Pharmaceutical companies** |
| **Joseph Maranville** | Bristol Myers Squibb, New York, NY, United States | **Steering Committee** | **Pharmaceutical companies** |
| **Mark McCarthy** | Genentech, San Francisco, CA, United States | **Steering Committee** | **Pharmaceutical companies** |
| **Margaret G. Ehm** | GlaxoSmithKline, Collegeville, PA, United States | **Steering Committee** | **Pharmaceutical companies** |
| **Kirsi Auro** | GlaxoSmithKline, Espoo, Finland | **Steering Committee** | **Pharmaceutical companies** |
| **Simonne Longerich** | Merck, Kenilworth, NJ, United States | **Steering Committee** | **Pharmaceutical companies** |
| **Anders Mälarstig** | Pfizer, New York, NY, United States | **Steering Committee** | **Pharmaceutical companies** |
| **Katherine Klinger** | Translational Sciences, Sanofi R&D, Framingham, MA, USA | **Steering Committee** | **Pharmaceutical companies** |
| **Clement Chatelain** | Translational Sciences, Sanofi R&D, Framingham, MA, USA | **Steering Committee** | **Pharmaceutical companies** |
| **Matthias Gossel** | Translational Sciences, Sanofi R&D, Framingham, MA, USA | **Steering Committee** | **Pharmaceutical companies** |
| **Karol Estrada** | Maze Therapeutics, San Francisco, CA, United States | **Steering Committee** | **Pharmaceutical companies** |
| **Robert Graham** | Maze Therapeutics, San Francisco, CA, United States | **Steering Committee** | **Pharmaceutical companies** |
| **Robert Yang** | Janssen Biotech, Beerse, Belgium | **Steering Committee** | **Pharmaceutical companies** |
| **Chris O´Donnell** | Novartis Institutes for BioMedical Research, Cambridge, MA, United States | **Steering Committee** | **Pharmaceutical companies** |
| **Tomi P. Mäkelä** | HiLIFE, University of Helsinki, Finland, Finland | **Steering Committee** | **University of Helsinki & Biobanks** |
| **Jaakko Kaprio** | Institute for Molecular Medicine Finland (FIMM), HiLIFE, University of Helsinki, Helsinki, Finland | **Steering Committee** | **University of Helsinki & Biobanks** |
| **Petri Virolainen** | Auria Biobank / University of Turku / Hospital District of Southwest Finland, Turku, Finland | **Steering Committee** | **University of Helsinki & Biobanks** |
| **Antti Hakanen** | Auria Biobank / University of Turku / Hospital District of Southwest Finland, Turku, Finland | **Steering Committee** | **University of Helsinki & Biobanks** |

| Full Name | Affiliation | Role 1 | Role 2 |
|---|---|---|---|
| **Terhi Kilpi** | THL Biobank / Finnish Institute for Health and Welfare (THL), Helsinki, Finland | Steering Committee | **University of Helsinki & Biobanks** |
| **Markus Perola** | THL Biobank / Finnish Institute for Health and Welfare (THL), Helsinki, Finland | Steering Committee | **University of Helsinki & Biobanks** |
| **Jukka Partanen** | Finnish Red Cross Blood Service / Finnish Hematology Registry and Clinical Biobank, Helsinki, Finland | Steering Committee | **University of Helsinki & Biobanks** |
| **Anne Pitkäranta** | Helsinki Biobank / Helsinki University and Hospital District of Helsinki and Uusimaa, Helsinki | Steering Committee | **University of Helsinki & Biobanks** |
| **Taneli Raivio** | Helsinki Biobank / Helsinki University and Hospital District of Helsinki and Uusimaa, Helsinki | Steering Committee | **University of Helsinki & Biobanks** |
| **Jani Tikkanen** | Northern Finland Biobank Borealis / University of Oulu / Northern Ostrobothnia Hospital District, Oulu, Finland | Steering Committee | **University of Helsinki & Biobanks** |
| **Raisa Serpi** | Northern Finland Biobank Borealis / University of Oulu / Northern Ostrobothnia Hospital District, Oulu, Finland | Steering Committee | **University of Helsinki & Biobanks** |
| **Tarja Laitinen** | Finnish Clinical Biobank Tampere / University of Tampere / Pirkanmaa Hospital District, Tampere, Finland | Steering Committee | **University of Helsinki & Biobanks** |
| **Veli-Matti Kosma** | Biobank of Eastern Finland / University of Eastern Finland / Northern Savo Hospital District, Kuopio, Finland | Steering Committee | **University of Helsinki & Biobanks** |
| **Jari Laukkanen** | Central Finland Biobank / University of Jyväskylä / Central Finland Health Care District, Jyväskylä, Finland | Steering Committee | **University of Helsinki & Biobanks** |
| **Marco Hautalahti** | FINBB - Finnish biobank cooperative | Steering Committee | **University of Helsinki & Biobanks** |
| **Outi Tuovila** | Business Finland, Helsinki, Finland | Steering Committee | **Other Experts/ Non-Voting Members** |
| **Raimo Pakkanen** | Business Finland, Helsinki, Finland | Steering Committee | **Other Experts/ Non-Voting Members** |
| **Jeffrey Waring** | Abbvie, Chicago, IL, United States | Scientific Committee | **Pharmaceutical companies** |
| **Bridget Riley-Gillis** | Abbvie, Chicago, IL, United States | Scientific Committee | **Pharmaceutical companies** |
| **Fedik Rahimov** | Abbvie, Chicago, IL, United States | Scientific Committee | **Pharmaceutical companies** |
| **Ioanna Tachmazidou** | Astra Zeneca, Cambridge, United Kingdom | Scientific Committee | **Pharmaceutical companies** |
| **Chia-Yen Chen** | Biogen, Cambridge, MA, United States | Scientific Committee | **Pharmaceutical companies** |
| **Heiko Runz** | Biogen, Cambridge, MA, United States | Scientific Committee | **Pharmaceutical companies** |
| **Zhihao Ding** | Boehringer Ingelheim, Ingelheim am Rhein, Germany | Scientific Committee | **Pharmaceutical companies** |
| **Marc Jung** | Boehringer Ingelheim, Ingelheim am Rhein, Germany | Scientific Committee | **Pharmaceutical companies** |
| **Shameek Biswas** | Bristol Myers Squibb, New York, NY, United States | Scientific Committee | **Pharmaceutical companies** |
| **Rion Pendergrass** | Genentech, San Francisco, CA, United States | Scientific Committee | **Pharmaceutical companies** |
| **Margaret G. Ehm** | GlaxoSmithKline, Collegeville, PA, United States | Scientific Committee | **Pharmaceutical companies** |
| **David Pulford** | GlaxoSmithKline, Stevenage, United Kingdom | Scientific Committee | **Pharmaceutical companies** |
| **Neha Raghavan** | Merck, Kenilworth, NJ, United States | Scientific Committee | **Pharmaceutical companies** |
| **Adriana Huertas-Vazquez** | Merck, Kenilworth, NJ, United States | Scientific Committee | **Pharmaceutical companies** |
| **Jae-Hoon Sul** | Merck, Kenilworth, NJ, United States | Scientific Committee | **Pharmaceutical companies** |
| **Anders Mälarstig** | Pfizer, New York, NY, United States | Scientific Committee | **Pharmaceutical companies** |
| **Xinli Hu** | Pfizer, New York, NY, United States | Scientific Committee | **Pharmaceutical companies** |

| Full Name | Affiliation | Role 1 | Role 2 |
|---|---|---|---|
| **Åsa Hedman** | Pfizer, New York, NY, United States | Scientific Committee | **Pharmaceutical companies** |
| **Katherine Klinger** | Translational Sciences, Sanofi R&D, Framingham, MA, USA | Scientific Committee | **Pharmaceutical companies** |
| **Robert Graham** | Maze Therapeutics, San Francisco, CA, United States | Scientific Committee | **Pharmaceutical companies** |
| **Manuel Rivas** | Maze Therapeutics, San Francisco, CA, United States | Scientific Committee | **Pharmaceutical companies** |
| **Dawn Waterworth** | Janssen Research & Development, LLC, Spring House, PA, United States | Scientific Committee | **Pharmaceutical companies** |
| **Nicole Renaud** | Novartis Institutes for BioMedical Research, Cambridge, MA, United States | Scientific Committee | **Pharmaceutical companies** |
| **Ma´en Obeidat** | Novartis Institutes for BioMedical Research, Cambridge, MA, United States | Scientific Committee | **Pharmaceutical companies** |
| **Samuli Ripatti** | Institute for Molecular Medicine Finland (FIMM), HiLIFE, University of Helsinki, Helsinki, Finland | Scientific Committee | **University of Helsinki & Biobanks** |
| **Johanna Schleutker** | Auria Biobank / Univ. of Turku / Hospital District of Southwest Finland, Turku, Finland | Scientific Committee | **University of Helsinki & Biobanks** |
| **Markus Perola** | THL Biobank / Finnish Institute for Health and Welfare (THL), Helsinki, Finland | Scientific Committee | **University of Helsinki & Biobanks** |
| **Mikko Arvas** | Finnish Red Cross Blood Service / Finnish Hematology Registry and Clinical Biobank, Helsinki, Finland | Scientific Committee | **University of Helsinki & Biobanks** |
| **Olli Carpén** | Helsinki Biobank / Helsinki University and Hospital District of Helsinki and Uusimaa, Helsinki | Scientific Committee | **University of Helsinki & Biobanks** |
| **Reetta Hinttala** | Northern Finland Biobank Borealis / University of Oulu / Northern Ostrobothnia Hospital District, Oulu, Finland | Scientific Committee | **University of Helsinki & Biobanks** |
| **Johannes Kettunen** | Northern Finland Biobank Borealis / University of Oulu / Northern Ostrobothnia Hospital District, Oulu, Finland | Scientific Committee | **University of Helsinki & Biobanks** |
| **Arto Mannermaa** | Biobank of Eastern Finland / University of Eastern Finland / Northern Savo Hospital District, Kuopio, Finland | Scientific Committee | **University of Helsinki & Biobanks** |
| **Katriina Aalto-Setälä** | Faculty of Medicine and Health Technology, Tampere University, Tampere, Finland | Scientific Committee | **University of Helsinki & Biobanks** |
| **Mika Kähönen** | Finnish Clinical Biobank Tampere / University of Tampere / Pirkanmaa Hospital District, Tampere, Finland | Scientific Committee | **University of Helsinki & Biobanks** |
| **Jari Laukkanen** | Central Finland Biobank / University of Jyväskylä / Central Finland Health Care District, Jyväskylä, Finland | Scientific Committee | **University of Helsinki & Biobanks** |
| **Johanna Mäkelä** | FINBB - Finnish biobank cooperative | Scientific Committee | **University of Helsinki & Biobanks** |
| **Reetta Kälviäinen** | Northern Savo Hospital District, Kuopio, Finland | Clinical Groups | **Neurology Group** |
| **Valtteri Julkunen** | Northern Savo Hospital District, Kuopio, Finland | Clinical Groups | **Neurology Group** |
| **Hilkka Soininen** | Northern Savo Hospital District, Kuopio, Finland | Clinical Groups | **Neurology Group** |
| **Anne Remes** | Northern Ostrobothnia Hospital District, Oulu, Finland | Clinical Groups | **Neurology Group** |
| **Mikko Hiltunen** | University of Eastern Finland, Kuopio, Finland | Clinical Groups | **Neurology Group** |
| **Jukka Peltola** | Pirkanmaa Hospital District, Tampere, Finland | Clinical Groups | **Neurology Group** |
| **Minna Raivio** | Hospital District of Helsinki and Uusimaa, Helsinki, Finland | Clinical Groups | **Neurology Group** |
| **Pentti Tienari** | Hospital District of Helsinki and Uusimaa, Helsinki, Finland | Clinical Groups | **Neurology Group** |
| **Juha Rinne** | Hospital District of Southwest Finland, Turku, Finland | Clinical Groups | **Neurology Group** |
| **Roosa Kallionpää** | Hospital District of Southwest Finland, Turku, Finland | Clinical Groups | **Neurology Group** |
| **Juulia Partanen** | Institute for Molecular Medicine Finland, HiLIFE, University of Helsinki, Finland | Clinical Groups | **Neurology Group** |

| Full Name | Affiliation | Role 1 | Role 2 |
|---|---|---|---|
| **Ali Abbasi** | Abbvie, Chicago, IL, United States | Clinical Groups | **Neurology Group** |
| **Adam Ziemann** | Abbvie, Chicago, IL, United States | Clinical Groups | **Neurology Group** |
| **Nizar Smaoui** | Abbvie, Chicago, IL, United States | Clinical Groups | **Neurology Group** |
| **Anne Lehtonen** | Abbvie, Chicago, IL, United States | Clinical Groups | **Neurology Group** |
| **Susan Eaton** | Biogen, Cambridge, MA, United States | Clinical Groups | **Neurology Group** |
| **Heiko Runz** | Biogen, Cambridge, MA, United States | Clinical Groups | **Neurology Group** |
| **Sanni Lahdenperä** | Biogen, Cambridge, MA, United States | Clinical Groups | **Neurology Group** |
| **Shameek Biswas** | Bristol Myers Squibb, New York, NY, United States | Clinical Groups | **Neurology Group** |
| **Natalie Bowers** | Genentech, San Francisco, CA, United States | Clinical Groups | **Neurology Group** |
| **Edmond Teng** | Genentech, San Francisco, CA, United States | Clinical Groups | **Neurology Group** |
| **Rion Pendergrass** | Genentech, San Francisco, CA, United States | Clinical Groups | **Neurology Group** |
| **Fanli Xu** | GlaxoSmithKline, Brentford, United Kingdom | Clinical Groups | **Neurology Group** |
| **David Pulford** | GlaxoSmithKline, Stevenage, United Kingdom | Clinical Groups | **Neurology Group** |
| **Kirsi Auro** | GlaxoSmithKline, Espoo, Finland | Clinical Groups | **Neurology Group** |
| **Laura Addis** | GlaxoSmithKline, Brentford, United Kingdom | Clinical Groups | **Neurology Group** |
| **John Eicher** | GlaxoSmithKline, Brentford, United Kingdom | Clinical Groups | **Neurology Group** |
| **Qingqin S Li** | Janssen Research & Development, LLC, Titusville, NJ 08560, United States | Clinical Groups | **Neurology Group** |
| **Karen He** | Janssen Research & Development, LLC, Spring House, PA, United States | Clinical Groups | **Neurology Group** |
| **Ekaterina Khramtsova** | Janssen Research & Development, LLC, Spring House, PA, United States | Clinical Groups | **Neurology Group** |
| **Neha Raghavan** | Merck, Kenilworth, NJ, United States | Clinical Groups | **Neurology Group** |
| **Martti Färkkilä** | Hospital District of Helsinki and Uusimaa, Helsinki, Finland | Clinical Groups | **Gastroenterology Group** |
| **Jukka Koskela** | Hospital District of Helsinki and Uusimaa, Helsinki, Finland | Clinical Groups | **Gastroenterology Group** |
| **Sampsa Pikkarainen** | Hospital District of Helsinki and Uusimaa, Helsinki, Finland | Clinical Groups | **Gastroenterology Group** |
| **Airi Jussila** | Pirkanmaa Hospital District, Tampere, Finland | Clinical Groups | **Gastroenterology Group** |
| **Katri Kaukinen** | Pirkanmaa Hospital District, Tampere, Finland | Clinical Groups | **Gastroenterology Group** |
| **Timo Blomster** | Northern Ostrobothnia Hospital District, Oulu, Finland | Clinical Groups | **Gastroenterology Group** |
| **Mikko Kiviniemi** | Northern Savo Hospital District, Kuopio, Finland | Clinical Groups | **Gastroenterology Group** |
| **Markku Voutilainen** | Hospital District of Southwest Finland, Turku, Finland | Clinical Groups | **Gastroenterology Group** |
| **Mark Daly** | Institute for Molecular Medicine, Finland (FIMM), HiLIFE, University of Helsinki, Helsinki, Finland; Broad Institute of MIT and Harvard; Massachusetts General Hospital | Clinical Groups | **Gastroenterology Group** |
| **Ali Abbasi** | Abbvie, Chicago, IL, United States | Clinical Groups | **Gastroenterology Group** |
| **Jeffrey Waring** | Abbvie, Chicago, IL, United States | Clinical Groups | **Gastroenterology Group** |
| **Nizar Smaoui** | Abbvie, Chicago, IL, United States | Clinical Groups | **Gastroenterology Group** |
| **Fedik Rahimov** | Abbvie, Chicago, IL, United States | Clinical Groups | **Gastroenterology Group** |
| **Anne Lehtonen** | Abbvie, Chicago, IL, United States | Clinical Groups | **Gastroenterology Group** |
| **Tim Lu** | Genentech, San Francisco, CA, United States | Clinical Groups | **Gastroenterology Group** |

| Full Name | Affiliation | Role 1 | Role 2 |
|---|---|---|---|
| **Natalie Bowers** | Genentech, San Francisco, CA, United States | Clinical Groups | **Gastroenterology Group** |
| **Rion Pendergrass** | Genentech, San Francisco, CA, United States | Clinical Groups | **Gastroenterology Group** |
| **Linda McCarthy** | GlaxoSmithKline, Brentford, United Kingdom | Clinical Groups | **Gastroenterology Group** |
| **Amy Hart** | Janssen Research & Development, LLC, Spring House, PA, United States | Clinical Groups | **Gastroenterology Group** |
| **Meijian Guan** | Janssen Research & Development, LLC, Spring House, PA, United States | Clinical Groups | **Gastroenterology Group** |
| **Jason Miller** | Merck, Kenilworth, NJ, United States | Clinical Groups | **Gastroenterology Group** |
| **Kirsi Kalpala** | Pfizer, New York, NY, United States | Clinical Groups | **Gastroenterology Group** |
| **Melissa Miller** | Pfizer, New York, NY, United States | Clinical Groups | **Gastroenterology Group** |
| **Xinli Hu** | Pfizer, New York, NY, United States | Clinical Groups | **Gastroenterology Group** |
| **Kari Eklund** | Hospital District of Helsinki and Uusimaa, Helsinki, Finland | Clinical Groups | **Rheumatology Group** |
| **Antti Palomäki** | Hospital District of Southwest Finland, Turku, Finland | Clinical Groups | **Rheumatology Group** |
| **Pia Isomäki** | Pirkanmaa Hospital District, Tampere, Finland | Clinical Groups | **Rheumatology Group** |
| **Laura Pirilä** | Hospital District of Southwest Finland, Turku, Finland | Clinical Groups | **Rheumatology Group** |
| **Oili Kaipiainen-Seppänen** | Northern Savo Hospital District, Kuopio, Finland | Clinical Groups | **Rheumatology Group** |
| **Johanna Huhtakangas** | Northern Ostrobothnia Hospital District, Oulu, Finland | Clinical Groups | **Rheumatology Group** |
| **Nina Mars** | Institute for Molecular Medicine Finland (FIMM), HiLIFE, University of Helsinki, Helsinki, Finland | Clinical Groups | **Rheumatology Group** |
| **Ali Abbasi** | Abbvie, Chicago, IL, United States | Clinical Groups | **Rheumatology Group** |
| **Jeffrey Waring** | Abbvie, Chicago, IL, United States | Clinical Groups | **Rheumatology Group** |
| **Fedik Rahimov** | Abbvie, Chicago, IL, United States | Clinical Groups | **Rheumatology Group** |
| **Apinya Lertratanakul** | Abbvie, Chicago, IL, United States | Clinical Groups | **Rheumatology Group** |
| **Nizar Smaoui** | Abbvie, Chicago, IL, United States | Clinical Groups | **Rheumatology Group** |
| **Anne Lehtonen** | Abbvie, Chicago, IL, United States | Clinical Groups | **Rheumatology Group** |
| **Coralie Viollet** | AstraZeneca, Cambridge, United Kingdom | Clinical Groups | **Rheumatology Group** |
| **Marla Hochfeld** | Bristol Myers Squibb, New York, NY, United States | Clinical Groups | **Rheumatology Group** |
| **Natalie Bowers** | Genentech, San Francisco, CA, United States | Clinical Groups | **Rheumatology Group** |
| **Rion Pendergrass** | Genentech, San Francisco, CA, United States | Clinical Groups | **Rheumatology Group** |
| **Jorge Esparza Gordillo** | GlaxoSmithKline, Brentford, United Kingdom | Clinical Groups | **Rheumatology Group** |
| **Kirsi Auro** | GlaxoSmithKline, Espoo, Finland | Clinical Groups | **Rheumatology Group** |
| **Dawn Waterworth** | Janssen Research & Development, LLC, Spring House, PA, United States | Clinical Groups | **Rheumatology Group** |
| **Fabiana Farias** | Merck, Kenilworth, NJ, United States | Clinical Groups | **Rheumatology Group** |
| **Kirsi Kalpala** | Pfizer, New York, NY, United States | Clinical Groups | **Rheumatology Group** |
| **Nan Bing** | Pfizer, New York, NY, United States | Clinical Groups | **Rheumatology Group** |
| **Xinli Hu** | Pfizer, New York, NY, United States | Clinical Groups | **Rheumatology Group** |
| **Tarja Laitinen** | Pirkanmaa Hospital District, Tampere, Finland | Clinical Groups | **Pulmonology Group** |
| **Margit Pelkonen** | Northern Savo Hospital District, Kuopio, Finland | Clinical Groups | **Pulmonology Group** |
| **Paula Kauppi** | Hospital District of Helsinki and Uusimaa, Helsinki, Finland | Clinical Groups | **Pulmonology Group** |

| Full Name | Affiliation | Role 1 | Role 2 |
|---|---|---|---|
| **Hannu Kankaanranta** | University of Gothenburg, Gothenburg, Sweden/ Seinäjoki Central Hospital, Seinäjoki, Finland/ Tampere University, Tampere, Finland | Clinical Groups | **Pulmonology Group** |
| **Terttu Harju** | Northern Ostrobothnia Hospital District, Oulu, Finland | Clinical Groups | **Pulmonology Group** |
| **Riitta Lahesmaa** | Hospital District of Southwest Finland, Turku, Finland | Clinical Groups | **Pulmonology Group** |
| **Nizar Smaoui** | Abbvie, Chicago, IL, United States | Clinical Groups | **Pulmonology Group** |
| **Coralie Viollet** | AstraZeneca, Cambridge, United Kingdom | Clinical Groups | **Pulmonology Group** |
| **Susan Eaton** | Biogen, Cambridge, MA, United States | Clinical Groups | **Pulmonology Group** |
| **Hubert Chen** | Genentech, San Francisco, CA, United States | Clinical Groups | **Pulmonology Group** |
| **Rion Pendergrass** | Genentech, San Francisco, CA, United States | Clinical Groups | **Pulmonology Group** |
| **Natalie Bowers** | Genentech, San Francisco, CA, United States | Clinical Groups | **Pulmonology Group** |
| **Joanna Betts** | GlaxoSmithKline, Brentford, United Kingdom | Clinical Groups | **Pulmonology Group** |
| **Kirsi Auro** | GlaxoSmithKline, Espoo, Finland | Clinical Groups | **Pulmonology Group** |
| **Rajashree Mishra** | GlaxoSmithKline, Brentford, United Kingdom | Clinical Groups | **Pulmonology Group** |
| **Majd Mouded** | Novartis, Basel, Switzerland | Clinical Groups | **Pulmonology Group** |
| **Debby Ngo** | Novartis, Basel, Switzerland | Clinical Groups | **Pulmonology Group** |
| **Teemu Niiranen** | Finnish Institute for Health and Welfare (THL), Helsinki, Finland | Clinical Groups | **Cardiometabolic Diseases Group** |
| **Felix Vaura** | Finnish Institute for Health and Welfare (THL), Helsinki, Finland | Clinical Groups | **Cardiometabolic Diseases Group** |
| **Veikko Salomaa** | Finnish Institute for Health and Welfare (THL), Helsinki, Finland | Clinical Groups | **Cardiometabolic Diseases Group** |
| **Kaj Metsärinne** | Hospital District of Southwest Finland, Turku, Finland | Clinical Groups | **Cardiometabolic Diseases Group** |
| **Jenni Aittokallio** | Hospital District of Southwest Finland, Turku, Finland | Clinical Groups | **Cardiometabolic Diseases Group** |
| **Mika Kähönen** | Pirkanmaa Hospital District, Tampere, Finland | Clinical Groups | **Cardiometabolic Diseases Group** |
| **Jussi Hernesniemi** | Pirkanmaa Hospital District, Tampere, Finland | Clinical Groups | **Cardiometabolic Diseases Group** |
| **Daniel Gordin** | Hospital District of Helsinki and Uusimaa, Helsinki, Finland | Clinical Groups | **Cardiometabolic Diseases Group** |
| **Juha Sinisalo** | Hospital District of Helsinki and Uusimaa, Helsinki, Finland | Clinical Groups | **Cardiometabolic Diseases Group** |
| **Marja-Riitta Taskinen** | Hospital District of Helsinki and Uusimaa, Helsinki, Finland | Clinical Groups | **Cardiometabolic Diseases Group** |
| **Tiinamaija Tuomi** | Hospital District of Helsinki and Uusimaa, Helsinki, Finland | Clinical Groups | **Cardiometabolic Diseases Group** |
| **Timo Hiltunen** | Hospital District of Helsinki and Uusimaa, Helsinki, Finland | Clinical Groups | **Cardiometabolic Diseases Group** |
| **Jari Laukkanen** | Central Finland Health Care District, Jyväskylä, Finland | Clinical Groups | **Cardiometabolic Diseases Group** |
| **Amanda Elliott** | Institute for Molecular Medicine Finland (FIMM), HiLIFE, University of Helsinki, Helsinki, Finland; Broad Institute, Cambridge, MA, USA and Massachusetts General Hospital, Boston, MA, USA | Clinical Groups | **Cardiometabolic Diseases Group** |
| **Mary Pat Reeve** | Institute for Molecular Medicine Finland (FIMM), HiLIFE, University of Helsinki, Helsinki, Finland | Clinical Groups | **Cardiometabolic Diseases Group** |
| **Sanni Ruotsalainen** | Institute for Molecular Medicine Finland (FIMM), HiLIFE, University of Helsinki, Helsinki, Finland | Clinical Groups | **Cardiometabolic Diseases Group** |
| **Dirk Paul** | Astra Zeneca, Cambridge, United Kingdom | Clinical Groups | **Cardiometabolic Diseases Group** |
| **Natalie Bowers** | Genentech, San Francisco, CA, United States | Clinical Groups | **Cardiometabolic Diseases Group** |
| **Rion Pendergrass** | Genentech, San Francisco, CA, United States | Clinical Groups | **Cardiometabolic Diseases Group** |
| **Audrey Chu** | GlaxoSmithKline, Brentford, United Kingdom | Clinical Groups | **Cardiometabolic Diseases Group** |

| Full Name | Affiliation | Role 1 | Role 2 |
|---|---|---|---|
| **Kirsi Auro** | GlaxoSmithKline, Espoo, Finland | Clinical Groups | **Cardiometabolic Diseases Group** |
| **Dermot Reilly** | Janssen Research & Development, LLC, Boston, MA, United States | Clinical Groups | **Cardiometabolic Diseases Group** |
| **Mike Mendelson** | Novartis, Boston, MA, United States | Clinical Groups | **Cardiometabolic Diseases Group** |
| **Jaakko Parkkinen** | Pfizer, New York, NY, United States | Clinical Groups | **Cardiometabolic Diseases Group** |
| **Melissa Miller** | Pfizer, New York, NY, United States | Clinical Groups | **Cardiometabolic Diseases Group** |
| **Tuomo Meretoja** | Hospital District of Helsinki and Uusimaa, Helsinki, Finland | Clinical Groups | **Oncology Group** |
| **Heikki Joensuu** | Hospital District of Helsinki and Uusimaa, Helsinki, Finland | Clinical Groups | **Oncology Group** |
| **Olli Carpén** | Hospital District of Helsinki and Uusimaa, Helsinki, Finland | Clinical Groups | **Oncology Group** |
| **Johanna Mattson** | Hospital District of Helsinki and Uusimaa, Helsinki, Finland | Clinical Groups | **Oncology Group** |
| **Eveliina Salminen** | Hospital District of Helsinki and Uusimaa, Helsinki, Finland | Clinical Groups | **Oncology Group** |
| **Annika Auranen** | Pirkanmaa Hospital District , Tampere, Finland | Clinical Groups | **Oncology Group** |
| **Peeter Karihtala** | Northern Ostrobothnia Hospital District, Oulu, Finland | Clinical Groups | **Oncology Group** |
| **Päivi Auvinen** | Northern Savo Hospital District, Kuopio, Finland | Clinical Groups | **Oncology Group** |
| **Klaus Elenius** | Hospital District of Southwest Finland, Turku, Finland | Clinical Groups | **Oncology Group** |
| **Johanna Schleutker** | Hospital District of Southwest Finland, Turku, Finland | Clinical Groups | **Oncology Group** |
| **Esa Pitkänen** | Institute for Molecular Medicine Finland (FIMM), HiLIFE, University of Helsinki, Helsinki, Finland | Clinical Groups | **Oncology Group** |
| **Nina Mars** | Institute for Molecular Medicine Finland (FIMM), HiLIFE, University of Helsinki, Helsinki, Finland | Clinical Groups | **Oncology Group** |
| **Mark Daly** | Institute for Molecular Medicine Finland (FIMM), HiLIFE, University of Helsinki, Helsinki, Finland; Broad Institute of MIT and Harvard; Massachusetts General Hospital | Clinical Groups | **Oncology Group** |
| **Relja Popovic** | Abbvie, Chicago, IL, United States | Clinical Groups | **Oncology Group** |
| **Jeffrey Waring** | Abbvie, Chicago, IL, United States | Clinical Groups | **Oncology Group** |
| **Bridget Riley-Gillis** | Abbvie, Chicago, IL, United States | Clinical Groups | **Oncology Group** |
| **Anne Lehtonen** | Abbvie, Chicago, IL, United States | Clinical Groups | **Oncology Group** |
| **Margarete Fabre** | AstraZeneca, Cambridge, United Kingdom | Clinical Groups | **Oncology Group** |
| **Jennifer Schutzman** | Genentech, San Francisco, CA, United States | Clinical Groups | **Oncology Group** |
| **Natalie Bowers** | Genentech, San Francisco, CA, United States | Clinical Groups | **Oncology Group** |
| **Rion Pendergrass** | Genentech, San Francisco, CA, United States | Clinical Groups | **Oncology Group** |
| **Diptee Kulkarni** | GlaxoSmithKline, Brentford, United Kingdom | Clinical Groups | **Oncology Group** |
| **Kirsi Auro** | GlaxoSmithKline, Espoo, Finland | Clinical Groups | **Oncology Group** |
| **Alessandro Porello** | Janssen Research & Development, LLC, Spring House, PA, United States | Clinical Groups | **Oncology Group** |
| **Andrey Loboda** | Merck, Kenilworth, NJ, United States | Clinical Groups | **Oncology Group** |
| **Heli Lehtonen** | Pfizer, New York, NY, United States | Clinical Groups | **Oncology Group** |
| **Stefan McDonough** | Pfizer, New York, NY, United States | Clinical Groups | **Oncology Group** |
| **Sauli Vuoti** | Janssen-Cilag Oy, Espoo, Finland | Clinical Groups | **Oncology Group** |
| **Kai Kaarniranta** | Northern Savo Hospital District, Kuopio, Finland; Department of Molecular Genetics, University of Lodz, Lodz, Poland | Clinical Groups | **Opthalmology Group** |

| Full Name | Affiliation | Role 1 | Role 2 |
|---|---|---|---|
| **Joni A Turunen** | Helsinki University Hospital and University of Helsinki, Helsinki, Finland; Eye Genetics Group, Folkhälsan Research Center, Helsinki, Finland | Clinical Groups | **Opthalmology Group** |
| **Terhi Ollila** | Hospital District of Helsinki and Uusimaa, Helsinki, Finland | Clinical Groups | **Opthalmology Group** |
| **Hannu Uusitalo** | Pirkanmaa Hospital District, Tampere, Finland | Clinical Groups | **Opthalmology Group** |
| **Juha Karjalainen** | Institute for Molecular Medicine Finland (FIMM), HiLIFE, University of Helsinki, Helsinki, Finland | Clinical Groups | **Opthalmology Group** |
| **Esa Pitkänen** | Institute for Molecular Medicine Finland (FIMM), HiLIFE, University of Helsinki, Helsinki, Finland | Clinical Groups | **Opthalmology Group** |
| **Mengzhen Liu** | Abbvie, Chicago, IL, United States | Clinical Groups | **Opthalmology Group** |
| **Heiko Runz** | Biogen, Cambridge, MA, United States | Clinical Groups | **Opthalmology Group** |
| **Stephanie Loomis** | Biogen, Cambridge, MA, United States | Clinical Groups | **Opthalmology Group** |
| **Erich Strauss** | Genentech, San Francisco, CA, United States | Clinical Groups | **Opthalmology Group** |
| **Natalie Bowers** | Genentech, San Francisco, CA, United States | Clinical Groups | **Opthalmology Group** |
| **Hao Chen** | Genentech, San Francisco, CA, United States | Clinical Groups | **Opthalmology Group** |
| **Rion Pendergrass** | Genentech, San Francisco, CA, United States | Clinical Groups | **Opthalmology Group** |
| **Kaisa Tasanen** | Northern Ostrobothnia Hospital District, Oulu, Finland | Clinical Groups | **Dermatology Group** |
| **Laura Huilaja** | Northern Ostrobothnia Hospital District, Oulu, Finland | Clinical Groups | **Dermatology Group** |
| **Katariina Hannula-Jouppi** | Hospital District of Helsinki and Uusimaa, Helsinki, Finland | Clinical Groups | **Dermatology Group** |
| **Teea Salmi** | Pirkanmaa Hospital District, Tampere, Finland | Clinical Groups | **Dermatology Group** |
| **Sirkku Peltonen** | Hospital District of Southwest Finland, Turku, Finland | Clinical Groups | **Dermatology Group** |
| **Leena Koulu** | Hospital District of Southwest Finland, Turku, Finland | Clinical Groups | **Dermatology Group** |
| **Nizar Smaoui** | Abbvie, Chicago, IL, United States | Clinical Groups | **Dermatology Group** |
| **Fedik Rahimov** | Abbvie, Chicago, IL, United States | Clinical Groups | **Dermatology Group** |
| **Anne Lehtonen** | Abbvie, Chicago, IL, United States | Clinical Groups | **Dermatology Group** |
| **David Choy** | Genentech, San Francisco, CA, United States | Clinical Groups | **Dermatology Group** |
| **Rion Pendergrass** | Genentech, San Francisco, CA, United States | Clinical Groups | **Dermatology Group** |
| **Dawn Waterworth** | Janssen Research & Development, LLC, Spring House, PA, United States | Clinical Groups | **Dermatology Group** |
| **Kirsi Kalpala** | Pfizer, New York, NY, United States | Clinical Groups | **Dermatology Group** |
| **Ying Wu** | Pfizer, New York, NY, United States | Clinical Groups | **Dermatology Group** |
| **Pirkko Pussinen** | Hospital District of Helsinki and Uusimaa, Helsinki, Finland | Clinical Groups | **Odontology Group** |
| **Aino Salminen** | Hospital District of Helsinki and Uusimaa, Helsinki, Finland | Clinical Groups | **Odontology Group** |
| **Tuula Salo** | Hospital District of Helsinki and Uusimaa, Helsinki, Finland | Clinical Groups | **Odontology Group** |
| **David Rice** | Hospital District of Helsinki and Uusimaa, Helsinki, Finland | Clinical Groups | **Odontology Group** |
| **Pekka Nieminen** | Hospital District of Helsinki and Uusimaa, Helsinki, Finland | Clinical Groups | **Odontology Group** |
| **Ulla Palotie** | Hospital District of Helsinki and Uusimaa, Helsinki, Finland | Clinical Groups | **Odontology Group** |
| **Maria Siponen** | Northern Savo Hospital District, Kuopio, Finland | Clinical Groups | **Odontology Group** |
| **Liisa Suominen** | Northern Savo Hospital District, Kuopio, Finland | Clinical Groups | **Odontology Group** |
| **Päivi Mäntylä** | Northern Savo Hospital District, Kuopio, Finland | Clinical Groups | **Odontology Group** |

| Full Name | Affiliation | Role 1 | Role 2 |
|---|---|---|---|
| **Ulvi Gursoy** | Hospital District of Southwest Finland, Turku, Finland | Clinical Groups | **Odontology Group** |
| **Vuokko Anttonen** | Northern Ostrobothnia Hospital District, Oulu, Finland | Clinical Groups | **Odontology Group** |
| **Kirsi Sipilä** | Research Unit of Oral Health Sciences Faculty of Medicine, University of Oulu, Oulu, Finland; Medical Research Center, Oulu, Oulu University Hospital and University of Oulu, Oulu, Finland | Clinical Groups | **Odontology Group** |
| **Rion Pendergrass** | Genentech, San Francisco, CA, United States | Clinical Groups | **Odontology Group** |
| **Hannele Laivuori** | Institute for Molecular Medicine Finland (FIMM), HiLIFE, University of Helsinki, Helsinki, Finland | Clinical Groups | **Women's Health and Reproduction Group** |
| **Venla Kurra** | Pirkanmaa Hospital District, Tampere, Finland | Clinical Groups | **Women's Health and Reproduction Group** |
| **Laura Kotaniemi-Talonen** | Pirkanmaa Hospital District, Tampere, Finland | Clinical Groups | **Women's Health and Reproduction Group** |
| **Oskari Heikinheimo** | Hospital District of Helsinki and Uusimaa, Helsinki, Finland | Clinical Groups | **Women's Health and Reproduction Group** |
| **Ilkka Kalliala** | Hospital District of Helsinki and Uusimaa, Helsinki, Finland | Clinical Groups | **Women's Health and Reproduction Group** |
| **Lauri Aaltonen** | Hospital District of Helsinki and Uusimaa, Helsinki, Finland | Clinical Groups | **Women's Health and Reproduction Group** |
| **Varpu Jokimaa** | Hospital District of Southwest Finland, Turku, Finland | Clinical Groups | **Women's Health and Reproduction Group** |
| **Johannes Kettunen** | Northern Ostrobothnia Hospital District, Oulu, Finland | Clinical Groups | **Women's Health and Reproduction Group** |
| **Marja Vääräsmäki** | Northern Ostrobothnia Hospital District, Oulu, Finland | Clinical Groups | **Women's Health and Reproduction Group** |
| **Outi Uimari** | Northern Ostrobothnia Hospital District, Oulu, Finland | Clinical Groups | **Women's Health and Reproduction Group** |
| **Laure Morin-Papunen** | Northern Ostrobothnia Hospital District, Oulu, Finland | Clinical Groups | **Women's Health and Reproduction Group** |
| **Maarit Niinimäki** | Northern Ostrobothnia Hospital District, Oulu, Finland | Clinical Groups | **Women's Health and Reproduction Group** |
| **Terhi Piltonen** | Northern Ostrobothnia Hospital District, Oulu, Finland | Clinical Groups | **Women's Health and Reproduction Group** |
| **Katja Kivinen** | Institute for Molecular Medicine Finland (FIMM), HiLIFE, University of Helsinki, Helsinki, Finland | Clinical Groups | **Women's Health and Reproduction Group** |
| **Elisabeth Widen** | Institute for Molecular Medicine Finland (FIMM), HiLIFE, University of Helsinki, Helsinki, Finland | Clinical Groups | **Women's Health and Reproduction Group** |
| **Taru Tukiainen** | Institute for Molecular Medicine Finland (FIMM), HiLIFE, University of Helsinki, Helsinki, Finland | Clinical Groups | **Women's Health and** |

| Full Name | Affiliation | Role 1 | Role 2 |
|---|---|---|---|
| | | | Reproduction Group |
| **Mary Pat Reeve** | Institute for Molecular Medicine Finland (FIMM), HiLIFE, University of Helsinki, Helsinki, Finland | Clinical Groups | **Women's Health and Reproduction Group** |
| **Mark Daly** | Institute for Molecular Medicine Finland (FIMM), HiLIFE, University of Helsinki, Helsinki, Finland; Broad Institute of MIT and Harvard; Massachusetts General Hospital | Clinical Groups | **Women's Health and Reproduction Group** |
| **Niko Välimäki** | University of Helsinki, Helsinki, Finland | Clinical Groups | **Women's Health and Reproduction Group** |
| **Eija Laakkonen** | University of Jyväskylä, Jyväskylä, Finland | Clinical Groups | **Women's Health and Reproduction Group** |
| **Jaakko Tyrmi** | University of Oulu, Oulu, Finland / University of Tampere, Tampere, Finland | Clinical Groups | **Women's Health and Reproduction Group** |
| **Heidi Silven** | University of Oulu, Oulu, Finland | Clinical Groups | **Women's Health and Reproduction Group** |
| **Eeva Sliz** | University of Oulu, Oulu, Finland | Clinical Groups | **Women's Health and Reproduction Group** |
| **Riikka Arffman** | University of Oulu, Oulu, Finland | Clinical Groups | **Women's Health and Reproduction Group** |
| **Susanna Savukoski** | University of Oulu, Oulu, Finland | Clinical Groups | **Women's Health and Reproduction Group** |
| **Triin Laisk** | Estonian biobank, Tartu, Estonia | Clinical Groups | **Women's Health and Reproduction Group** |
| **Natalia Pujol** | Estonian biobank, Tartu, Estonia | Clinical Groups | **Women's Health and Reproduction Group** |
| **Mengzhen Liu** | Abbvie, Chicago, IL, United States | Clinical Groups | **Women's Health and Reproduction Group** |
| **Bridget Riley-Gillis** | Abbvie, Chicago, IL, United States | Clinical Groups | **Women's Health and Reproduction Group** |
| **Rion Pendergrass** | Genentech, San Francisco, CA, United States | Clinical Groups | **Women's Health and Reproduction Group** |
| **Janet Kumar** | GlaxoSmithKline, Collegeville, PA, United States | Clinical Groups | **Women's Health and Reproduction Group** |
| **Kirsi Auro** | GlaxoSmithKline, Espoo, Finland | Clinical Groups | **Women's Health and Reproduction Group** |
| **Iiris Hovatta** | University of Helsinki, Finland | Clinical Groups | **Depression group** |
| **Chia-Yen Chen** | Biogen, Cambridge, MA, United States | Clinical Groups | **Depression group** |
| **Erkki Isometsä** | Hospital District of Helsinki and Uusimaa, Helsinki, Finland | Clinical Groups | **Depression group** |

| Full Name | Affiliation | Role 1 | Role 2 |
|---|---|---|---|
| **Hanna Ollila** | Institute for Molecular Medicine Finland (FIMM), HiLIFE, University of Helsinki, Helsinki, Finland | **Clinical Groups** | **Depression group** |
| **Jaana Suvisaari** | Finnish Institute for Health and Welfare (THL), Helsinki, Finland | **Clinical Groups** | **Depression group** |
| **Thomas Damm Als** | Aarhus University, Denmark | **Clinical Groups** | **Depression group** |
| **Antti Mäkitie** | Department of Otorhinolaryngology - Head and Neck Surgery, University of Helsinki and Helsinki University Hospital, Helsinki, Finland | **Clinical Groups** | **ENT (ear, nose and throath) Group** |
| **Argyro Bizaki-Vallaskangas** | Pirkanmaa Hospital District, Tampere, Finland | **Clinical Groups** | **ENT (ear, nose and throath) Group** |
| **Sanna Toppila-Salmi** | University of Eastern Finland and Kuopio University Hospital, Department of Otorhinolaryngology, Kuopio, Finland and Department of Allergy, Helsinki University Hospital and University of Helsinki, Finland | **Clinical Groups** | **ENT (ear, nose and throath) Group** |
| **Tytti Willberg** | Hospital District of Southwest Finland, Turku, Finland | **Clinical Groups** | **ENT (ear, nose and throath) Group** |
| **Elmo Saarentaus** | Institute for Molecular Medicine Finland (FIMM), HiLIFE, University of Helsinki, Helsinki, Finland | **Clinical Groups** | **ENT (ear, nose and throath) Group** |
| **Antti Aarnisalo** | Hospital District of Helsinki and Uusimaa, Helsinki, Finland | **Clinical Groups** | **ENT (ear, nose and throath) Group** |
| **Eveliina Salminen** | Hospital District of Helsinki and Uusimaa, Helsinki, Finland | **Clinical Groups** | **ENT (ear, nose and throath) Group** |
| **Elisa Rahikkala** | Northern Ostrobothnia Hospital District, Oulu, Finland | **Clinical Groups** | **ENT (ear, nose and throath) Group** |
| **Johannes Kettunen** | Northern Ostrobothnia Hospital District, Oulu, Finland | **Clinical Groups** | **ENT (ear, nose and throath) Group** |
| **Kristiina Aittomäki** | Department of Medical Genetics, Helsinki University Central Hospital, Helsinki, Finland | **Clinical Groups** | **POI (premature ovarian failure) Group** |
| **Fredrik Åberg** | Transplantation and Liver Surgery Clinic, Helsinki University Hospital, Helsinki University, Helsinki, Finland | **Clinical Groups** | **LiverScore Group** |
| **Mitja Kurki** | Institute for Molecular Medicine Finland (FIMM), HiLIFE, University of Helsinki, Helsinki, Finland; Broad Institute, Cambridge, MA, United States | **FinnGen Analysis working group** | **FinnGen Analysis working group** |
| **Samuli Ripatti** | Institute for Molecular Medicine Finland (FIMM), HiLIFE, University of Helsinki, Helsinki, Finland | **FinnGen Analysis working group** | **FinnGen Analysis working group** |
| **Mark Daly** | Institute for Molecular Medicine, Finland (FIMM), HiLIFE, University of Helsinki, Helsinki, Finland; Broad Institute of MIT and Harvard; Massachusetts General Hospital | **FinnGen Analysis working group** | **FinnGen Analysis working group** |
| **Juha Karjalainen** | Institute for Molecular Medicine Finland (FIMM), HiLIFE, University of Helsinki, Helsinki, Finland | **FinnGen Analysis working group** | **FinnGen Analysis working group** |
| **Aki Havulinna** | Institute for Molecular Medicine Finland (FIMM), HiLIFE, University of Helsinki, Helsinki, Finland; Finnish Institute for Health and Welfare (THL), Helsinki, Finland | **FinnGen Analysis working group** | **FinnGen Analysis working group** |
| **Juha Mehtonen** | Institute for Molecular Medicine Finland (FIMM), HiLIFE, University of Helsinki, Helsinki, Finland | **FinnGen Analysis working group** | **FinnGen Analysis working group** |
| **Priit Palta** | Institute for Molecular Medicine Finland (FIMM), HiLIFE, University of Helsinki, Helsinki, Finland | **FinnGen Analysis working group** | **FinnGen Analysis working group** |
| **Shabbeer Hassan** | Institute for Molecular Medicine Finland (FIMM), HiLIFE, University of Helsinki, Helsinki, Finland | **FinnGen Analysis working group** | **FinnGen Analysis working group** |

| Full Name | Affiliation | Role 1 | Role 2 |
|---|---|---|---|
| **Pietro Della Briotta Parolo** | Institute for Molecular Medicine Finland (FIMM), HiLIFE, University of Helsinki, Helsinki, Finland | FinnGen Analysis working group | FinnGen Analysis working group |
| **Wei Zhou** | Broad Institute, Cambridge, MA, United States | FinnGen Analysis working group | FinnGen Analysis working group |
| **Mutaamba Maasha** | Broad Institute, Cambridge, MA, United States | FinnGen Analysis working group | FinnGen Analysis working group |
| **Shabbeer Hassan** | Institute for Molecular Medicine Finland (FIMM), HiLIFE, University of Helsinki, Helsinki, Finland | FinnGen Analysis working group | FinnGen Analysis working group |
| **Susanna Lemmelä** | Institute for Molecular Medicine Finland (FIMM), HiLIFE, University of Helsinki, Helsinki, Finland | FinnGen Analysis working group | FinnGen Analysis working group |
| **Manuel Rivas** | University of Stanford, Stanford, CA, United States | FinnGen Analysis working group | FinnGen Analysis working group |
| **Aarno Palotie** | Institute for Molecular Medicine Finland (FIMM), HiLIFE, University of Helsinki, Helsinki, Finland | FinnGen Analysis working group | FinnGen Analysis working group |
| **Aoxing Liu** | Institute for Molecular Medicine Finland (FIMM), HiLIFE, University of Helsinki, Helsinki, Finland | FinnGen Analysis working group | FinnGen Analysis working group |
| **Arto Lehisto** | Institute for Molecular Medicine Finland (FIMM), HiLIFE, University of Helsinki, Helsinki, Finland | FinnGen Analysis working group | FinnGen Analysis working group |
| **Andrea Ganna** | Institute for Molecular Medicine Finland (FIMM), HiLIFE, University of Helsinki, Helsinki, Finland | FinnGen Analysis working group | FinnGen Analysis working group |
| **Vincent Llorens** | Institute for Molecular Medicine Finland (FIMM), HiLIFE, University of Helsinki, Helsinki, Finland | FinnGen Analysis working group | FinnGen Analysis working group |
| **Hannele Laivuori** | Institute for Molecular Medicine Finland (FIMM), HiLIFE, University of Helsinki, Helsinki, Finland | FinnGen Analysis working group | FinnGen Analysis working group |
| **Taru Tukiainen** | Institute for Molecular Medicine Finland (FIMM), HiLIFE, University of Helsinki, Helsinki, Finland | FinnGen Analysis working group | FinnGen Analysis working group |
| **Mary Pat Reeve** | Institute for Molecular Medicine Finland (FIMM), HiLIFE, University of Helsinki, Helsinki, Finland | FinnGen Analysis working group | FinnGen Analysis working group |
| **Henrike Heyne** | Institute for Molecular Medicine Finland (FIMM), HiLIFE, University of Helsinki, Helsinki, Finland | FinnGen Analysis working group | FinnGen Analysis working group |
| **Nina Mars** | Institute for Molecular Medicine Finland (FIMM), HiLIFE, University of Helsinki, Helsinki, Finland | FinnGen Analysis working group | FinnGen Analysis working group |
| **Joel Rämö** | Institute for Molecular Medicine Finland (FIMM), HiLIFE, University of Helsinki, Helsinki, Finland | FinnGen Analysis working group | FinnGen Analysis working group |
| **Elmo Saarentaus** | Institute for Molecular Medicine Finland (FIMM), HiLIFE, University of Helsinki, Helsinki, Finland | FinnGen Analysis working group | FinnGen Analysis working group |

| Full Name | Affiliation | Role 1 | Role 2 |
|---|---|---|---|
| **Hanna Ollila** | Institute for Molecular Medicine Finland (FIMM), HiLIFE, University of Helsinki, Helsinki, Finland | FinnGen Analysis working group | FinnGen Analysis working group |
| **Rodos Rodosthenous** | Institute for Molecular Medicine Finland (FIMM), HiLIFE, University of Helsinki, Helsinki, Finland | FinnGen Analysis working group | FinnGen Analysis working group |
| **Satu Strausz** | Institute for Molecular Medicine Finland (FIMM), HiLIFE, University of Helsinki, Helsinki, Finland | FinnGen Analysis working group | FinnGen Analysis working group |
| **Tuula Palotie** | University of Helsinki and Hospital District of Helsinki and Uusimaa, Helsinki, Finland | FinnGen Analysis working group | FinnGen Analysis working group |
| **Kimmo Palin** | University of Helsinki, Helsinki, Finland | FinnGen Analysis working group | FinnGen Analysis working group |
| **Javier Garcia-Tabuenca** | University of Tampere, Tampere, Finland | FinnGen Analysis working group | FinnGen Analysis working group |
| **Harri Siirtola** | University of Tampere, Tampere, Finland | FinnGen Analysis working group | FinnGen Analysis working group |
| **Tuomo Kiiskinen** | Institute for Molecular Medicine Finland (FIMM), HiLIFE, University of Helsinki, Helsinki, Finland | FinnGen Analysis working group | FinnGen Analysis working group |
| **Jiwoo Lee** | Institute for Molecular Medicine Finland (FIMM), HiLIFE, University of Helsinki, Helsinki, Finland; Broad Institute, Cambridge, MA, United States | FinnGen Analysis working group | FinnGen Analysis working group |
| **Kristin Tsuo** | Institute for Molecular Medicine Finland (FIMM), HiLIFE, University of Helsinki, Helsinki, Finland; Broad Institute, Cambridge, MA, United States | FinnGen Analysis working group | FinnGen Analysis working group |
| **Amanda Elliott** | Institute for Molecular Medicine Finland (FIMM), HiLIFE, University of Helsinki, Helsinki, Finland; Broad Institute, Cambridge, MA, USA and Massachusetts General Hospital, Boston, MA, USA | FinnGen Analysis working group | FinnGen Analysis working group |
| **Kati Kristiansson** | THL Biobank / Finnish Institute for Health and Welfare (THL), Helsinki, Finland | FinnGen Analysis working group | FinnGen Analysis working group |
| **Mikko Arvas** | Finnish Red Cross Blood Service / Finnish Hematology Registry and Clinical Biobank, Helsinki, Finland | FinnGen Analysis working group | FinnGen Analysis working group |
| **Kati Hyvärinen** | Finnish Red Cross Blood Service, Helsinki, Finland | FinnGen Analysis working group | FinnGen Analysis working group |
| **Jarmo Ritari** | Finnish Red Cross Blood Service, Helsinki, Finland | FinnGen Analysis working group | FinnGen Analysis working group |
| **Olli Carpén** | Helsinki Biobank / Helsinki University and Hospital District of Helsinki and Uusimaa, Helsinki | FinnGen Analysis working group | FinnGen Analysis working group |
| **Johannes Kettunen** | Northern Finland Biobank Borealis / University of Oulu / Northern Ostrobothnia Hospital District, Oulu, Finland | FinnGen Analysis working group | FinnGen Analysis working group |
| **Katri Pylkäs** | University of Oulu, Oulu, Finland | FinnGen Analysis working group | FinnGen Analysis working group |

| Full Name | Affiliation | Role 1 | Role 2 |
|---|---|---|---|
| **Eeva Sliz** | University of Oulu, Oulu, Finland | **FinnGen Analysis working group** | **FinnGen Analysis working group** |
| **Minna Karjalainen** | University of Oulu, Oulu, Finland | **FinnGen Analysis working group** | **FinnGen Analysis working group** |
| **Tuomo Mantere** | Northern Finland Biobank Borealis / University of Oulu / Northern Ostrobothnia Hospital District, Oulu, Finland | **FinnGen Analysis working group** | **FinnGen Analysis working group** |
| **Eeva Kangasniemi** | Finnish Clinical Biobank Tampere / University of Tampere / Pirkanmaa Hospital District, Tampere, Finland | **FinnGen Analysis working group** | **FinnGen Analysis working group** |
| **Sami Heikkinen** | University of Eastern Finland, Kuopio, Finland | **FinnGen Analysis working group** | **FinnGen Analysis working group** |
| **Arto Mannermaa** | Biobank of Eastern Finland / University of Eastern Finland / Northern Savo Hospital District, Kuopio, Finland | **FinnGen Analysis working group** | **FinnGen Analysis working group** |
| **Eija Laakkonen** | University of Jyväskylä, Jyväskylä, Finland | **FinnGen Analysis working group** | **FinnGen Analysis working group** |
| **Nina Pitkänen** | Auria Biobank / University of Turku / Hospital District of Southwest Finland, Turku, Finland | **FinnGen Analysis working group** | **FinnGen Analysis working group** |
| **Samuel Lessard** | Translational Sciences, Sanofi R&D, Framingham, MA, USA | **FinnGen Analysis working group** | **FinnGen Analysis working group** |
| **Clément Chatelain** | Translational Sciences, Sanofi R&D, Framingham, MA, USA | **FinnGen Analysis working group** | **FinnGen Analysis working group** |
| **Lila Kallio** | Auria Biobank / University of Turku / Hospital District of Southwest Finland, Turku, Finland | **Biobank directors** | **Biobank directors** |
| **Tiina Wahlfors** | THL Biobank / Finnish Institute for Health and Welfare (THL), Helsinki, Finland | **Biobank directors** | **Biobank directors** |
| **Jukka Partanen** | Finnish Red Cross Blood Service / Finnish Hematology Registry and Clinical Biobank, Helsinki, Finland | **Biobank directors** | **Biobank directors** |
| **Eero Punkka** | Helsinki Biobank / Helsinki University and Hospital District of Helsinki and Uusimaa, Helsinki | **Biobank directors** | **Biobank directors** |
| **Raisa Serpi** | Northern Finland Biobank Borealis / University of Oulu / Northern Ostrobothnia Hospital District, Oulu, Finland | **Biobank directors** | **Biobank directors** |
| **Sanna Siltanen** | Finnish Clinical Biobank Tampere / University of Tampere / Pirkanmaa Hospital District, Tampere, Finland | **Biobank directors** | **Biobank directors** |
| **Veli-Matti Kosma** | Biobank of Eastern Finland / University of Eastern Finland / Northern Savo Hospital District, Kuopio, Finland | **Biobank directors** | **Biobank directors** |
| **Teijo Kuopio** | Central Finland Biobank / University of Jyväskylä / Central Finland Health Care District, Jyväskylä, Finland | **Biobank directors** | **Biobank directors** |
| **Anu Jalanko** | Institute for Molecular Medicine Finland (FIMM), HiLIFE, University of Helsinki, Helsinki, Finland | **FinnGen Teams** | **Administration** |
| **Huei-Yi Shen** | Institute for Molecular Medicine Finland (FIMM), HiLIFE, University of Helsinki, Helsinki, Finland | **FinnGen Teams** | **Administration** |
| **Risto Kajanne** | Institute for Molecular Medicine Finland (FIMM), HiLIFE, University of Helsinki, Helsinki, Finland | **FinnGen Teams** | **Administration** |
| **Mervi Aavikko** | Institute for Molecular Medicine Finland (FIMM), HiLIFE, University of Helsinki, Helsinki, Finland | **FinnGen Teams** | **Administration** |
| **Helen Cooper** | Institute for Molecular Medicine Finland (FIMM), HiLIFE, University of Helsinki, Helsinki, Finland | **FinnGen Teams** | **Administration** |
| **Denise Öller** | Institute for Molecular Medicine Finland (FIMM), HiLIFE, University of Helsinki, Helsinki, Finland | **FinnGen Teams** | **Administration** |
| **Rasko Leinonen** | Institute for Molecular Medicine Finland (FIMM), HiLIFE, University of Helsinki, Helsinki, Finland; European Molecular Biology Laboratory, European Bioinformatics Institute, Cambridge, UK | **FinnGen Teams** | **Administration** |

| Full Name | Affiliation | Role 1 | Role 2 |
|---|---|---|---|
| **Henna Palin** | Finnish Clinical Biobank Tampere / University of Tampere / Pirkanmaa Hospital District, Tampere, Finland | FinnGen Teams | **Administration** |
| **Malla-Maria Linna** | Helsinki Biobank / Helsinki University and Hospital District of Helsinki and Uusimaa, Helsinki | FinnGen Teams | **Administration** |
| **Mitja Kurki** | Institute for Molecular Medicine Finland (FIMM), HiLIFE, University of Helsinki, Helsinki, Finland; Broad Institute, Cambridge, MA, United States | FinnGen Teams | **Analysis** |
| **Juha Karjalainen** | Institute for Molecular Medicine Finland (FIMM), HiLIFE, University of Helsinki, Helsinki, Finland | FinnGen Teams | **Analysis** |
| **Pietro Della Briotta Parolo** | Institute for Molecular Medicine Finland (FIMM), HiLIFE, University of Helsinki, Helsinki, Finland | FinnGen Teams | **Analysis** |
| **Arto Lehisto** | Institute for Molecular Medicine Finland (FIMM), HiLIFE, University of Helsinki, Helsinki, Finland | FinnGen Teams | **Analysis** |
| **Juha Mehtonen** | Institute for Molecular Medicine Finland (FIMM), HiLIFE, University of Helsinki, Helsinki, Finland | FinnGen Teams | **Analysis** |
| **Wei Zhou** | Broad Institute, Cambridge, MA, United States | FinnGen Teams | **Analysis** |
| **Masahiro Kanai** | Broad Institute, Cambridge, MA, United States | FinnGen Teams | **Analysis** |
| **Mutaamba Maasha** | Broad Institute, Cambridge, MA, United States | FinnGen Teams | **Analysis** |
| **Zhili Zheng** | Broad Institute, Cambridge, MA, United States | FinnGen Teams | **Analysis** |
| **Hannele Laivuori** | Institute for Molecular Medicine Finland (FIMM), HiLIFE, University of Helsinki, Helsinki, Finland | FinnGen Teams | **Clinical Endpoint Development** |
| **Aki Havulinna** | Institute for Molecular Medicine Finland (FIMM), HiLIFE, University of Helsinki, Helsinki, Finland; Finnish Institute for Health and Welfare (THL), Helsinki, Finland | FinnGen Teams | **Clinical Endpoint Development** |
| **Susanna Lemmelä** | Institute for Molecular Medicine Finland (FIMM), HiLIFE, University of Helsinki, Helsinki, Finland | FinnGen Teams | **Clinical Endpoint Development** |
| **Tuomo Kiiskinen** | Institute for Molecular Medicine Finland (FIMM), HiLIFE, University of Helsinki, Helsinki, Finland | FinnGen Teams | **Clinical Endpoint Development** |
| **L. Elisa Lahtela** | Institute for Molecular Medicine Finland (FIMM), HiLIFE, University of Helsinki, Helsinki, Finland | FinnGen Teams | **Clinical Endpoint Development** |
| **Mari Kaunisto** | Institute for Molecular Medicine Finland (FIMM), HiLIFE, University of Helsinki, Helsinki, Finland | FinnGen Teams | **Communication** |
| **Elina Kilpeläinen** | Institute for Molecular Medicine Finland (FIMM), HiLIFE, University of Helsinki, Helsinki, Finland | FinnGen Teams | **E-Science** |
| **Timo P. Sipilä** | Institute for Molecular Medicine Finland (FIMM), HiLIFE, University of Helsinki, Helsinki, Finland | FinnGen Teams | **E-Science** |
| **Oluwaseun Alexander Dada** | Institute for Molecular Medicine Finland (FIMM), HiLIFE, University of Helsinki, Helsinki, Finland | FinnGen Teams | **E-Science** |
| **Awaisa Ghazal** | Institute for Molecular Medicine Finland (FIMM), HiLIFE, University of Helsinki, Helsinki, Finland | FinnGen Teams | **E-Science** |
| **Anastasia Kytölä** | Institute for Molecular Medicine Finland (FIMM), HiLIFE, University of Helsinki, Helsinki, Finland | FinnGen Teams | **E-Science** |
| **Rigbe Weldatsadik** | Institute for Molecular Medicine Finland (FIMM), HiLIFE, University of Helsinki, Helsinki, Finland | FinnGen Teams | **E-Science** |
| **Sanni Ruotsalainen** | Institute for Molecular Medicine Finland (FIMM), HiLIFE, University of Helsinki, Helsinki, Finland | FinnGen Teams | **E-Science** |
| **Kati Donner** | Institute for Molecular Medicine Finland (FIMM), HiLIFE, University of Helsinki, Helsinki, Finland | FinnGen Teams | **Genotyping** |
| **Timo P. Sipilä** | Institute for Molecular Medicine Finland (FIMM), HiLIFE, University of Helsinki, Helsinki, Finland | FinnGen Teams | **Genotyping** |
| **Anu Loukola** | Helsinki Biobank / Helsinki University and Hospital District of Helsinki and Uusimaa, Helsinki | FinnGen Teams | **Sample Collection Coordination** |
| **Päivi Laiho** | THL Biobank / Finnish Institute for Health and Welfare (THL), Helsinki, Finland | FinnGen Teams | **Sample Logistics** |
| **Tuuli Sistonen** | THL Biobank / Finnish Institute for Health and Welfare (THL), Helsinki, Finland | FinnGen Teams | **Sample Logistics** |
| **Essi Kaiharju** | THL Biobank / Finnish Institute for Health and Welfare (THL), Helsinki, Finland | FinnGen Teams | **Sample Logistics** |
| **Markku Laukkanen** | THL Biobank / Finnish Institute for Health and Welfare (THL), Helsinki, Finland | FinnGen Teams | **Sample Logistics** |
| **Elina Järvensivu** | THL Biobank / Finnish Institute for Health and Welfare (THL), Helsinki, Finland | FinnGen Teams | **Sample Logistics** |
| **Sini Lähteenmäki** | THL Biobank / Finnish Institute for Health and Welfare (THL), Helsinki, Finland | FinnGen Teams | **Sample Logistics** |
| **Lotta Männikkö** | THL Biobank / Finnish Institute for Health and Welfare (THL), Helsinki, Finland | FinnGen Teams | **Sample Logistics** |

| Full Name | Affiliation | Role 1 | Role 2 |
|---|---|---|---|
| **Regis Wong** | THL Biobank / Finnish Institute for Health and Welfare (THL), Helsinki, Finland | FinnGen Teams | **Sample Logistics** |
| **Auli Toivola** | THL Biobank / Finnish Institute for Health and Welfare (THL), Helsinki, Finland | FinnGen Teams | **Sample Logistics** |
| **Minna Brunfeldt** | THL Biobank / Finnish Institute for Health and Welfare (THL), Helsinki, Finland | FinnGen Teams | **Registry Data Operations** |
| **Hannele Mattsson** | THL Biobank / Finnish Institute for Health and Welfare (THL), Helsinki, Finland | FinnGen Teams | **Registry Data Operations** |
| **Kati Kristiansson** | THL Biobank / Finnish Institute for Health and Welfare (THL), Helsinki, Finland | FinnGen Teams | **Registry Data Operations** |
| **Susanna Lemmelä** | Institute for Molecular Medicine Finland (FIMM), HiLIFE, University of Helsinki, Helsinki, Finland | FinnGen Teams | **Registry Data Operations** |
| **Sami Koskelainen** | THL Biobank / Finnish Institute for Health and Welfare (THL), Helsinki, Finland | FinnGen Teams | **Registry Data Operations** |
| **Tero Hiekkalinna** | THL Biobank / Finnish Institute for Health and Welfare (THL), Helsinki, Finland | FinnGen Teams | **Registry Data Operations** |
| **Teemu Paajanen** | THL Biobank / Finnish Institute for Health and Welfare (THL), Helsinki, Finland | FinnGen Teams | **Registry Data Operations** |
| **Priit Palta** | Institute for Molecular Medicine Finland (FIMM), HiLIFE, University of Helsinki, Helsinki, Finland | FinnGen Teams | **Sequencing Informatics** |
| **Kalle Pärn** | Institute for Molecular Medicine Finland (FIMM), HiLIFE, University of Helsinki, Helsinki, Finland | FinnGen Teams | **Sequencing Informatics** |
| **Mart Kals** | Institute for Molecular Medicine Finland (FIMM), HiLIFE, University of Helsinki, Helsinki, Finland | FinnGen Teams | **Sequencing Informatics** |
| **Shuang Luo** | Institute for Molecular Medicine Finland (FIMM), HiLIFE, University of Helsinki, Helsinki, Finland | FinnGen Teams | **Sequencing Informatics** |
| **Tarja Laitinen** | Pirkanmaa Hospital District, Tampere, Finland | FinnGen Teams | **Trajectory** |
| **Mary Pat Reeve** | Institute for Molecular Medicine Finland (FIMM), HiLIFE, University of Helsinki, Helsinki, Finland | FinnGen Teams | **Trajectory** |
| **Shanmukha Sampath Padmanabhuni** | Institute for Molecular Medicine Finland (FIMM), HiLIFE, University of Helsinki, Helsinki, Finland | FinnGen Teams | **Trajectory** |
| **Marianna Niemi** | University of Tampere, Tampere, Finland | FinnGen Teams | **Trajectory** |
| **Harri Siirtola** | University of Tampere, Tampere, Finland | FinnGen Teams | **Trajectory** |
| **Javier Gracia-Tabuenca** | University of Tampere, Tampere, Finland | FinnGen Teams | **Trajectory** |
| **Mika Helminen** | University of Tampere, Tampere, Finland | FinnGen Teams | **Trajectory** |
| **Tiina Luukkaala** | University of Tampere, Tampere, Finland | FinnGen Teams | **Trajectory** |
| **Iida Vähätalo** | University of Tampere, Tampere, Finland | FinnGen Teams | **Trajectory** |
| **Jyrki Tammerluoto** | Institute for Molecular Medicine Finland (FIMM), HiLIFE, University of Helsinki, Helsinki, Finland | FinnGen Teams | **Data protection officer** |
| **Marco Hautalahti** | Finnish Biobank Cooperative - FINBB | FinnGen Teams | **FINBB - Finnish biobank cooperative** |
| **Johanna Mäkelä** | Finnish Biobank Cooperative - FINBB | FinnGen Teams | **FINBB - Finnish biobank cooperative** |
| **Sarah Smith** | Finnish Biobank Cooperative - FINBB | FinnGen Teams | **FINBB - Finnish biobank cooperative** |
| **Tom Southerington** | Finnish Biobank Cooperative - FINBB | FinnGen Teams | **FINBB - Finnish biobank cooperative** |
| **Petri Lehto** | Finnish Biobank Cooperative - FINBB | FinnGen Teams | **FINBB - Finnish biobank cooperative** |

**Table S10.** A list Estonian Biobank Research Team authors and their affiliations

| Full Name | Affiliation |
|---|---|
| **Andres Metspalu** | Estonian Genome Centre, Institute of Genomics, University of Tartu, Tartu, Estonia |
| **Mari Nelis** | Estonian Genome Centre, Institute of Genomics, University of Tartu, Tartu, Estonia |
| **Lili Milani** | Estonian Genome Centre, Institute of Genomics, University of Tartu, Tartu, Estonia |
| **Reedik Mägi** | Estonian Genome Centre, Institute of Genomics, University of Tartu, Tartu, Estonia |
| **Georgi Hudjashov** | Estonian Genome Centre, Institute of Genomics, University of Tartu, Tartu, Estonia |
| **Tõnu Esko** | Estonian Genome Centre, Institute of Genomics, University of Tartu, Tartu, Estonia |
